# A clinical laboratory test-wide association scan of polygenic scores identifies biomarkers of psychiatric disorders

**DOI:** 10.64898/2026.10.15.26365624

**Authors:** Makayla L. Reed, Kyra L. Feuer, Penn Medicine BioBank, Rachel L. Kember

## Abstract

**Background:** Psychiatric disorders pose a significant public health burden, and their complex genetic architecture underlies multiple biological pathways, contributing to a lack of diagnostic biomarkers. Laboratory-Wide Association Studies (LabWAS) leverage electronic health record laboratory data as intermediary phenotypes to link genetic risk to physiology.

**Methods:** Polygenic Scores (PGS) for 25 health-related, psychiatric, and substance use phenotypes were generated for individuals of African (AFR; N=10,473) and European (EUR, N=35,131) ancestry from the Penn Medicine Biobank. Laboratory data was prepared using the LabWAS pipeline, encompassing 78 laboratory tests and 6 vital signs. Linear regression models tested the association of each lab value with PGS adjusting for sex, age, and ten within-ancestry genetic principal components.

**Results:** Four PGS were significantly associated with at least one laboratory value in AFR individuals and 22 PGS in EUR individuals. In EUR individuals, psychiatric PGS partitioned into three physiological profiles: anxiety, attention-deficit/hyperactivity disorder, major depressive disorder, and post-traumatic stress disorder shared a metabolic/inflammatory signature (higher BMI, glucose, and monocytes; lower high density lipoprotein cholesterol and oxygen saturation); anorexia nervosa, obsessive compulsive disorder, and schizophrenia were characterized by lower BMI; and bipolar disorder demonstrated a more circumscribed profile with lower electrolytes and renal makers. Substance use disorder PGS similarly clustered: tobacco use disorder and externalizing showed metabolic/inflammatory profiles; alcohol-related phenotypes showed lower red blood cell indices; and cannabis use disorder had a more circumscribed profile.

**Conclusions:** LabWAS reveals physiologic signatures associated with genetic risk for psychiatric disorders, offering new biological insights into their pathology.

## INTRODUCTION

Psychiatric disorders are among the leading contributors to morbidity and mortality worldwide (1), and are frequently comorbid with somatic conditions such as immune, metabolic, and cardiovascular disorders (2,3). However, in contrast to these somatic conditions, psychiatric disorders are usually diagnosed through reported symptoms alone rather than measurement of physiological biomarkers. A lack of objective biomarkers for psychiatric disorders and their symptoms limits accurate diagnosis and monitoring of treatment response.

Psychiatric disorders are highly heritable, with genetic variance explaining between 34% to 77% of the observed phenotypic variance(4). Genome-wide association studies (GWAS) have identified hundreds of loci associated with these conditions, and, in combination with polygenic scores (PGS), have provided a widely used foundation for quantifying genetic risk as a potential biomarker. However, translating these findings into biological insight and clinical applications remains a significant challenge (2,3).

Laboratory measurements are a potential intermediary phenotype that are easily accessible and could facilitate this translation. Although disease status is often thought of in binary terms, many conditions are defined by laboratory thresholds, highlighting that disease often exists on a spectrum. Similarly, while genetic markers are static, laboratory values fluctuate across the lifespan, including across disease development, and could be utilized to predict timing of disease onset in individuals at high genetic risk. Lab measurements can therefore serve as more sensitive indicators of health. Several laboratory measurements have already been shown to differ in populations with psychiatric conditions. Major depressive disorder (MDD) has been linked to elevated levels of inflammatory markers such as C-reactive protein (CRP) and inflammatory cytokines, as well as higher body mass index (BMI)(9–11). Attention deficit hyperactivity disorder (ADHD) has been associated with obesity in both children and adults(12); and bipolar disorder and schizophrenia have both been associated with metabolites and immune markers including IL-6 and cortisol (13–15). However, it is unclear whether these associations were caused by shared genetic liability or the effects of medication, lifestyle, or downstream consequences of illness.

The increasing availability of large-scale electronic health records (EHRs) has provided an unprecedented resource for biomedical research. While phenome-wide association studies (PheWAS) have been instrumental in mapping genetic risk to phenotypes and identifying novel pleiotropic genetic effects(16), there have been limited genetic studies that have leveraged the breadth of clinical lab measurements in EHRs. Laboratory-Wide Association Studies (LabWAS) are an emerging framework that address this gap by testing associations between genetic factors and multiple clinical measurements(17). By treating these routine clinical measurements as phenotypes, LabWAS provides a powerful high-throughput method to discover biological links with genetic risk, allowing us to uncover disease mechanisms and improve the clinical interpretability of PGS.

Psychiatric disorders represent a compelling domain for LabWAS applications because their complex, polygenic architecture may influence a wide range of measurable biological traits, providing an opportunity to identify biomarkers associated with disease-related pathways. Here, we conducted a LabWAS of PGS for psychiatric disorders and related comorbidities within the Penn Medicine BioBank (PMBB). The primary aim was to identify physiological biomarkers associated with genetic risk for psychiatric disorders, as captured through PGS; as well as to identify clusters of psychiatric disorders whose genetic liability manifests in similar physiological associations. To assess whether observed associations were robust to potential confounding by differences in body composition or presence of disorder, sensitivity analyses accounted for BMI and presence of ICD codes for the primary disorder. These results can inform the development of precision psychiatry approaches to enable earlier intervention, individualized monitoring, and target prevention and treatment strategies.

## METHODS

### Penn Medicine Biobank Cohort

The PMBB recruits participants through the University of Pennsylvania Health System. At enrollment, participants provide informed consent, providing either a blood or tissue sample, and grant permission for researchers to access their EHR information. The study was approved by the Institutional Review Board of the University of Pennsylvania. As of March 2026, PMBB includes 356,654 enrolled participants, of whom 57,170 have undergone genome-wide genotyping.

### Genotyping and QC, Phasing and Imputation, Ancestry Estimation

Genotype data available in PMBB for this study included N=57,170 individuals, consisting of N=43,913 samples from Freeze 2.0 genotyped using the Illumina Global Screening Array (GSA) and N=13,257 samples from Freeze 3.0 genotyped by sequencing (GxS) derived from whole-exome sequencing. Due to the variant sets captured by array genotyping and sequencing differing substantially (650,281 variants in Freeze 2.0, 1,414,695 in Freeze 3.0, with only 77,515 shared), the datasets were processed separately prior to imputation and then merged (Supplemental Methods).

Ancestry estimation was performed to assign ancestral superclasses to each sample, resulting in N=41,651 European (EUR) samples, and N=12,042 African/African American (AFR) samples (Supplemental Methods).

### Lab Values

Clinical laboratory test values were extracted from the EHR of PMBB individuals. A LabWAS pipeline derived from the approach developed by Dennis et al.(17) was applied to PMBB in order to generate a LabWAS matrix file of the age adjusted average lab values for each individual. After data normalization and quality filtering a total of 84 clinical biomarkers remained: 3 body composition, 6 vitals, 7 liver function, 6 endocrine, 16 hematologic, 15 immune, 20 metabolic, 10 renal function, and 1 muscular. The final analytic sample included up to 54,952 individuals, with variation in sample size across the biomarkers reflecting differences in clinical testing frequency (Supplementary Table S1).

### PGS

PGS were calculated using PRS-CS(18), using ancestry-matched linkage-disequilibrium (LD) reference panels from the 1000 Genomes Project (19) and ancestry matched summary statistics (Supplementary Table 2). For psychiatric conditions, GWAS that were well-powered were selected, whereas for non-psychiatric conditions, GWASs for traits with known associated clinical biomarkers were chosen. Individual level PGS were calculated using PLINK v1.9 and standardized to have a mean of zero and standard deviation of one.

### Statistical Analysis

Associations between PGS and clinical laboratory traits were calculated using the LabWAS framework. The age adjusted average laboratory value, covariates, and PGS for each individual were merged within ancestry (max N AFR=10,473, max N EUR=35,131). Following the merge, 83 lab values remained. Linear regression models were fit for each clinical lab test where the lab test values were the outcome, and the PGS was the primary predictor, adjusting for relevant covariates (sex, within ancestry genetic PCs1:10). Analyses were restricted to clinical lab tests that had a minimum sample threshold of N=100. Multiple testing correction was performed using both Bonferroni adjustment and false discovery rate (FDR).

## RESULTS

### Sample Characteristics

Across the laboratory tests, the mean age for individuals was 64 years old and sex distribution was relatively even (49.3 % male, 50.7% female). A PGS for a total of 25 phenotypes were generated and tested against the LabWAS dataset: 9 psychiatric, 7 substance use disorders, 7 health related, 1 neurological, and 1 sleep phenotype (Supplementary Table S2). Individuals of AMR, EAS, and SAS ancestry were not included due to sample size limitations.

### Overview of PGS with Significant Associations

Of the 25 phenotypes (all analyzed in EUR, and 9 in AFR), 4 had at least one significantly associated lab by Bonferroni correction in AFR patients, and 22 in EUR patients.

### Primary Associations of Health Phenotype PGSs

To confirm the robustness of laboratory results, PGS of health phenotypes with well-established associations were tested. As expected, PGS for BMI was positively associated with BMI (OR= 1.4, p≤ 2.2 x 10^-308^; Supplementary Fig. S1; Supplementary Table S3); PGS for Type 2 diabetes (T2D) was positively associated with glucose (OR= 1.3, p=2.2 x 10^-308^) and HbA1c (OR= 1.3, p=2.2 x 10^-308^; Supplementary Fig. S2; Supplementary Table S4); PGS for high-density lipoprotein (HDL) cholesterol was positively associated with HDL cholesterol (OR= 1.34, p≤2.20 x 10^-16^; Supplementary Fig. S3; Supplementary Table S5); PGS for low-density lipoprotein (LDL) cholesterol was positively associated with LDL cholesterol (OR= 1.2, p=3.7 x 10^-213^; Supplementary Fig. S4; Supplementary Table S6); PGS for total cholesterol (TC) was positively associated with TC (OR= 1.24, p=5.05 x 10^-196^); Supplementary Fig. S5; Supplementary Table S7); PGS for total triglycerides (TG) was positively associated with TG (OR= 1.33, p≤ 2.20 x 10^-16^; Supplementary Fig. S6; Supplementary Table S8); and PGS for coronary artery disease (CAD) was negatively associated with HDL cholesterol (OR= 0.96, p=1.3 x 10^-11^; Supplementary Fig. S7; Supplementary Table S9), consistent with findings from previous studies(20,21). The Alzheimer’s PGS was also significantly associated with LDL, TC, and HDL values (OR= 0.97-1.06, all p≤ 8.50 x 10^-5^); Supplementary Fig. S8; Supplementary Table S10).

### Primary Associations of Psychiatric PGSs

In individuals of European ancestry, psychiatric PGS partitioned into different physiological clusters. A cluster of disorders including anxiety (ANX, Supplementary Fig. S9; Supplementary Table S11), ADHD (Supplementary Fig. S10; Supplementary Table S12), MDD (Supplementary Fig. S11; Supplementary Table S13), and post-traumatic stress disorder (PTSD, Supplementary Fig. S12; Supplementary Table S14) demonstrated a shared biological profile characterized by metabolic dysregulation and inflammation. All four PGS were significantly associated with increased BMI (OR= 1.04 - 1.08; all p≤ 1.1 x 10^-13^) and elevated glycemic/lipid markers, specifically triglycerides (OR= 1.05 - 1.08; all p≤ 3.8 x 10^-12^) and glucose (OR= 1.04 -1.05; all p≤ 6.2 x 10^-11^). Conversely, all four were negatively associated with HDL cholesterol (OR= 0.94 - 0.96; all p≤ 3.7 x 10^-11^). This group also showed consistent physiological and inflammatory signatures. Higher polygenic risk was associated with increased pulse rate (OR= 1.03 - 1.04; all p≤ 8.7 x 10^-9^) and lower oxygen saturation (OR= ∼0.96; all p≤ 7.8 x 10^-12^); as well as evidence of immune involvement with positive associations observed for monocytes (OR= 1.02 - 1.03; all p≤ 6.6 × 10⁻⁵) and neutrophils (OR= 1.02 - 1.04; all p≤ 1.3 × 10⁻⁴). Secondary clusters emerged within this group; ADHD and ANX were associated with increased respiratory rate (OR= 1.03 - 1.04; all p≤ 1.1 × 10⁻⁵) and lower hemoglobin (OR= 0.96 - 0.98; all p≤ 2.2 × 10⁻⁴), while ADHD, MDD, and PTSD shared an association with lower total bilirubin (OR= 0.97 - 0.98; all p≤ 4.8 × 10⁻⁵).

In contrast, polygenic risk for anorexia nervosa (AN, Supplementary Fig. S13; Supplementary Table S15), obsessive-compulsive disorder (OCD, Supplementary Fig. S14; Supplementary Table S16) and schizophrenia (SCZ, Supplementary Fig. S15; Supplementary Table S17) showed an opposing pattern characterized by lower body composition measures. All scores were associated with lower BMI (OR= 0.97 - 0.98; all p ≤ 4.1 × 10⁻⁶) and reduced body weight (OR= 0.97 - 0.98; all p≤ 1.0 × 10⁻⁶). Among these disorders, AN demonstrated the most extensive metabolic associations, with lower HbA1c (OR= 0.95; p=3.2 × 10⁻⁸), glucose (OR= 0.97; p=8.5 × 10⁻⁸), and triglyceride (OR= 0.96; p=9.5 × 10⁻⁶) markers. AN also showed positive associations with cholesterol markers (HDL (OR= 1.05; p=4.5 × 10⁻⁹) and total cholesterol (OR= 1.03; p=3.3 × 10⁻⁴)), albumin (OR= 1.02; p=4.9 × 10⁻⁴), and oxygen saturation (OR= 1.03; p=1.4 × 10⁻⁴). PGS_SCZ_ was uniquely associated with lower urinary creatinine (OR= 0.96; p=4.2 × 10⁻⁴).

Polygenic risk for bipolar disorder (BD, Supplementary Fig. S16; Supplementary Table S18) showed a more circumscribed profile that differed from both clusters. Higher genetic risk for BD was associated with lower electrolyte and renal markers, including chloride (OR= 0.97; p=9.1 × 10⁻⁶), sodium (OR= 0.98; p=1.8 × 10⁻⁴), and creatinine (OR= 0.98; p=3.7 × 10⁻⁶), while showing positive associations with albumin (OR= 1.03; p=1.9 × 10⁻⁴) and HDL cholesterol (OR= 1.03; p=6.1 × 10⁻⁴).

In individuals of African ancestry, fewer associations were detected compared with those observed in European ancestry individuals. While several psychiatric PGS in the European ancestry analysis demonstrated consistent cardiometabolic and inflammatory signatures, only BD (PGS_BD_ Supplementary Fig. S17; Supplementary Table S19) and PTSD PGS (PGS_PTSD_ Supplementary Fig. S18; Supplementary Table S20) showed significant associations in the African ancestry cohort. PGS_BD_ was negatively associated with neutrophils (OR= 0.95, p=7.4 × 10^-7^), whole blood cell count (WBC) (OR= 0.95, p=9.2 × 10^-6^), monocytes (OR= 0.96, p=2.2 × 10^-4^), and blood urea nitrogen (BUN) (OR= 0.96, p=4.5 × 10^-4^). PGS_PTSD_ was negatively associated with HbA1c (OR= 0.94, p=6.9 × 10^-5^) and potassium (OR= 0.95, p=3.9 × 10^-4^), while being positively associated with WBC (OR= 1.06, p=4.8 × 10^-6^) and neutrophils (OR= 1.06, p=2.7 × 10^-5^). The immune-related associations observed for PTSD in African ancestry individuals were directionally consistent with the inflammatory signals observed for several psychiatric PGS in the European ancestry analysis, although the overall number of associations was substantially reduced.

### Primary Associations of SUD PGSs

In individuals of European ancestry, SUD PGS partitioned into distinct physiological clusters. A cluster including tobacco use disorder (TUD, Supplementary Fig. S19; Supplementary Table S21) and the externalizing factor (EXT, Supplementary Fig. S20; Supplementary Table S22) demonstrated a shared profile characterized by metabolic dysregulation and inflammation. Both PGS were significantly associated with increased BMI (OR =1.05; all p≤ 9.0×10⁻¹⁸), elevated glycemic and lipid markers, specifically glucose (OR= 1.04; all p≤ 2.1×10⁻¹⁰), triglycerides (OR= 1.05; all p≤ 5.6×10⁻¹⁰), and HbA1c (OR= 1.03 - 1.04; all p≤ 1.0×10⁻⁴), and increased pulse rate (OR range = 1.02–1.03; all p≤ 3.8×10⁻^5^). Conversely, both were negatively associated with oxygen saturation (OR= 0.95–0.96; all p≤ 5.7×10⁻¹³), hemoglobin (OR= 0.97; all p≤ 1.1×10⁻⁹), HDL cholesterol (OR= 0.96 - 0.97; all p≤ 1.2×10⁻^6^), albumin (OR= 0.96 - 0.97; all p≤ 2.1×10⁻⁶), sodium (OR= 0.97; all p≤ 8.3×10⁻^7^), and total bilirubin (OR= 0.97 - 0.98; all p≤ 7.7×10⁻⁵). Both PGS also showed evidence of immune involvement, with positive associations observed for monocytes (OR= 1.03 - 1.05; all p≤ 8.7×10⁻^6^) and neutrophils (OR= 1.03 - 1.04; all p≤ 4.6×10⁻⁴). TUD was additionally associated with elevated CRP (OR= 1.07; p=9.2×10⁻⁷) and ESR (OR= 1.05; p=5.5×10⁻⁶), while EXT showed an association with RBC (OR= 0.97; p=4.1×10⁻¹⁰).

In contrast, polygenic risk for alcohol-related phenotypes, including problematic alcohol use (PAU, Supplementary Fig. S21; Supplementary Table S23) and alcohol use disorder (AUD, Supplementary Fig. S22; Supplementary Table S24) showed an opposing pattern characterized by lower red blood cell indices. Both PGS were negatively associated with RBC (OR= 0.96 - 0.97; all p≤ 1.6×10⁻⁸) and hemoglobin (OR= 0.96 - 0.97; all p≤ 1.9×10⁻⁶), while showing positive associations with HDL cholesterol (OR= 1.03; all p≤ 1.2×10⁻^4^). Among these, AUD demonstrated additional associations, with additional negative associations with sodium (OR= 0.97; p=3.9×10⁻⁵). PAU was additionally negatively associated with BUN (OR= 0.98; p=4.5×10⁻⁴).

Polygenic risk for the addiction factor (AF, Supplementary Fig. S23; Supplementary Table S25) showed associations that aligned with both clusters. Higher genetic risk for AF was negatively associated with hemoglobin (OR= 0.97; p=1.6×10⁻⁸), RBC (OR= 0.97; p=5.5×10⁻⁷), oxygen saturation (OR= 0.97; p=2.7×10⁻⁶), and sodium (OR= 0.98; p=9.2×10⁻⁶), while showing positive associations with glucose (OR= 1.02; p=1.7×10⁻⁴), urinary WBC (OR= 1.05; p=1.5×10⁻⁵), and respiratory rate (OR= 1.02; p=5.8×10⁻⁴). Cannabis use disorder (CUD, Supplementary Fig. S24; Supplementary Table S26) showed a more limited profile, with negative associations with oxygen saturation (OR= 0.98; p=1.1×10⁻⁴) and height (OR= 0.98; p=1.6×10⁻⁴), and a positive association with triglycerides (OR= 1.03; p=2.6×10⁻⁴).

In individuals of African ancestry, significantly fewer associations were detected compared with those observed in European ancestry individuals. Only TUD (Supplementary Fig. S25; Supplementary Table S27) and CUD (Supplementary Fig. S26; Supplementary Table S28) PGS showed significant associations in the African ancestry cohort. PGS TUD was negatively associated with WBC (OR= 0.96; p=1.2×10⁻⁵), and polygenic risk for both TUD and CUD were negatively associated with neutrophils (TUD: OR= 0.96, p=2.5×10⁻⁴; CUD: OR= 0.96, p=5.7×10⁻⁴).

### Controlling for BMI and primary diagnosis

To evaluate whether associations were independent of clinical diagnosis and adiposity, sensitivity analyses were conducted adjusting for BMI and the presence of a diagnosis code for the primary phenotype. Overall, the association patterns remained directionally consistent after adjustment, indicating that most observed associations were not fully explained by BMI and diagnostic status.

All PGS with no significant results are presented in the supplementary tables and figures (AFR: AF, Supplementary Table 27, Supplementary Fig. S29; AUD, Supplementary Table 28, Supplementary Fig. S30; MDD, Supplementary table 31, Supplementary Fig. S33; Opioid Use Disorder (OUD), Supplementary Table 32, Supplementary Fig. S34; SCZ, Supplementary Table 34, Supplementary Fig. S36; EUR: Autism Spectrum Disorder (ASD), Supplementary Table 29, Supplementary Fig. S31; Chronotype, Supplementary table 30, Supplementary Fig. S32; OUD, Supplementary table 33, Supplementary Fig. S35).

## DISCUSSION

In this study, a comprehensive LabWAS was conducted within the Penn Medicine BioBank, testing associations between polygenic scores for 25 different PGS and 83 clinical lab tests in individuals of African and European ancestry. PGSs with known physiological associations were used to test the LabWAS pipeline. PGS for BMI, T2D, HDL, LDL, TC, TG, and CAD all showed strong associations with expected biomarkers. Findings for psychiatric and substance use disorders demonstrate that genetic risk is broadly reflected in measurable physiological and clinical traits, such as cardiometabolic, hematologic, and immune system markers. Primary associations remained directionally consistent after adjustment for BMI and primary diagnoses, indicating that these associations were not solely driven by adiposity or disease status, suggesting that genetic liability itself influences physiological associations.

A key finding of this study is the clustering of phenotypes based on their laboratory value association profiles. Notably, psychiatric PGS portioned into distinct physiological clusters, with ANX, ADHD, MDD, and PTSD forming one cluster and AN, OCD, and SCZ forming a second. This pattern is consistent with the underlying genomic factor structure of psychiatric disorders identified by Grotzinger et al. which identified an Internalizing factor defined by PTSD, MDD, and ANX, and a Compulsive factor defined by AN, OCD, and Tourette’s Syndrome (TS) (22).

In the EUR ancestry cohort, the first psychiatric cluster observed included ANX, ADHD, MDD, and PTSD, consistent with the internalizing factor. This cluster’s profile was highly metabolic with elevated BMI, triglycerides, glucose, and pulse rate, and lower HDL cholesterol levels. These findings align with existing research linking these disorders with metabolic dysregulation. Elevated BMI and dyslipidemia have been repeatedly reported in clinical populations with MDD and PTSD, and prior observational and Mendelian randomization studies have suggested bidirectional relationships between depression and metabolic dysfunction (23,24). Additionally, all four disorders in this cluster showed associations with immune markers, including elevated monocytes and neutrophils, providing laboratory-level evidence for the inflammatory hypothesis of psychiatric illness. Elevated inflammatory markers, particularly CRP, neutrophils, and monocyte count, have been reported in studies of MDD and PTSD (25–27). Notably, in the AFR ancestry cohort, PTSD also demonstrated immune-related signals that were directionally consistent with those observed in EUR.

The second cluster, again identified in the EUR ancestry cohort and aligned with the compulsive factor, encompassed AN, OCD, and SCZ, which had a contrasting pattern of lower body composition, with reduced BMI and body weight. A meta-analysis of antipsychotic-naïve and minimally treated patients with psychosis revealed lower BMI compared to healthy controls (28). As expected, AN showed the most extensive metabolic associations with lower glucose, HbA1c, and triglycerides, alongside higher HDL, and total cholesterol. Studies in individuals with AN have reported the glucometabolic homeostasis is significantly lower in AN individuals when compared to normal-weight healthy individuals (29). Paradoxical hypercholesterolemia is a feature of AN that has been seen dating back to the 1960s, with more recent studies backing the idea that there is lipid dysregulation in AN individuals as higher cholesterol levels remained after weight restoration, indicating that there may be a complex relationship between liver function, lipid metabolism, and hormones, rather than the effects of starvation (30–32).

Furthermore, Watson et al. identified eight loci to be significantly associated with AN, including variants associated with lower BMI and higher HDL cholesterol levels, supporting a shared genetic basis between AN and metabolic traits (33). PGS for SCZ was uniquely associated with lower urinary creatinine. Urinary creatinine excretion has been proposed as a marker of metabolically active skeletal muscle mass (34,35), these findings may be consistent with previous reports of reduced skeletal muscle mass, poorer muscle quality, and impaired physical functioning in individuals with schizophrenia (36,37). While there were no significant associations for the AFR ancestry cohort, directionality for several labs remained consistent with those found in Bigdeli et al (38). These convergences were mainly seen in renal and hepatic markers (such as BUN, creatine, and AST/ALT), but notable divergences in lipid profiles. Consistency when looking at the EUR cohort were more robust, with directional alignment in metabolic and inflammatory markers like HbA1c, ferritin, chloride, and HDL.

EUR BD’s profile did not fully align with either cluster as its associations were with lower electrolyte and renal markers, which remain significant after controlling for a BD diagnosis. Predisposition to hyponatremia (low sodium levels) in BD is well documented, although connected back to medication side effects (39). The present associations here were based on polygenic liability rather than medication exposure; however, because lab measures can be influenced by medications commonly used to treat BD, these findings may reflect treatment-related side effects. While the observed associations are consistent with GWAS evidence for BD encompassing genes related to ion channels (40,41), further analyses accounting for medication exposure are needed.

SUD PGS similarly partitioned into distinct clusters. TUD and the externalizing factor demonstrated a profile mirroring that of the internalizing psychiatric cluster: elevated BMI, glycemic markers, triglycerides, and pulse, alongside lower oxygen saturation, hemoglobin, HDL, albumin, sodium, and total bilirubin, with positive immune associations including monocytes, neutrophils, CRP, and ESR. Chronic substance exposure has been linked to immune and inflammatory dysregulation, with smoking, specifically associated with changes in adaptive immunity and low-grade inflammation (42). In contrast, alcohol-related PGS (PAU and AUD) showed a distinct pattern of lower red blood cell indices, reduced RBC and hemoglobin, with elevated HDL, aligning with evidence that alcohol use has hematological consequences as excess alcohol consumption reduces RBC production and leads to impaired hemoglobin synthesis (43). The AF PGS profile overlapped with those of TUD and alcohol, while CUD’s had association with lower oxygen saturation and elevated triglycerides.

This study’s findings need to be interpreted in light of several limitations. First, this study was conducted within a single biobank setting, with the availability of laboratory data relied on for clinical care and not from a research standpoint. Consequently, laboratory measurements may reflect underlying health status and healthcare utilization patterns which may introduce selection bias. To analyze these laboratory phenotypes, the QualityLab pipeline was used to summarize the longitudinal data within the EHR by reducing the measurements into a single median value for each individual. Due to this, associations of laboratory values and the onset or progression of the phenotypes observed cannot be derived. Secondly, while sensitivity analyses adjusting for BMI and ICD-coded diagnoses support the independence of most associations, residual confounding unmeasured variables, including medication use, lifestyle factors, and socioeconomic status are possible.

There are several avenues which future research can take alongside this study. Firstly, longitudinal studies are needed to determine whether the biomarker profiles seen associated with psychiatric and substance use disorder polygenic liabilities precede disease onset, evolve during illness progression, or arise due to treatment effects. Biomarker associations should also be evaluated in the influence of behavioral factors such as diet, physical activity, and sleep. Finally, with the availability of clinical laboratory data in EHR there is an opportunity to examine whether combining PGS with biomarker profiles can improve the prediction of disease onset, trajectory, health comorbidities, and treatment response.

**Table 1.**
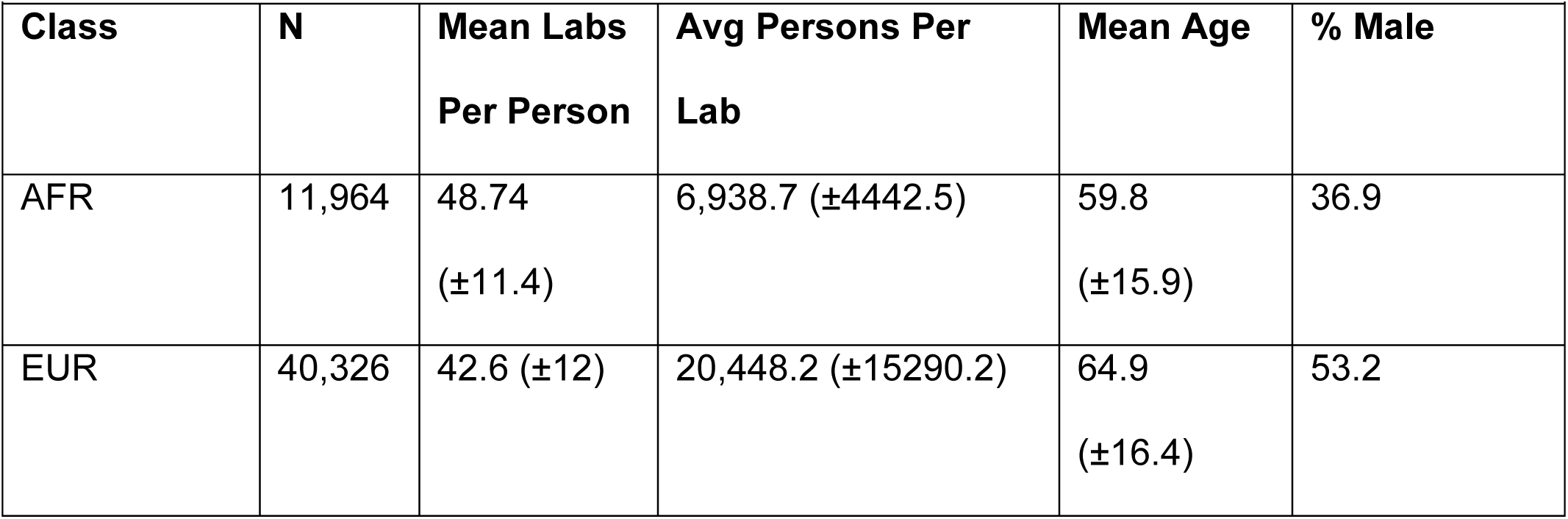
Demographic characteristics and laboratory data availability by ancestry class. Statistics for individuals stratified by ancestry class (Supplemental Table 2). For each class, the table reports the total number of individuals (N), the mean number of labs values per individual, the average number of individuals per lab, mean age, and sex distribution. The average number of individuals per lab test was calculated across all lab tests based on non-missing values.

| Table 1 |  |  |  |  |  |
| --- | --- | --- | --- | --- | --- |
| Class | N | Mean Labs<br>Per Person | Avg Persons Per<br>Lab | Mean Age | % Male |
| AFR | 11,964 | 48.74<br>($\pm 11.4$) | 6,938.7 ($\pm 4442.5$) | 59.8<br>($\pm 15.9$) | 36.9 |
| EUR | 40,326 | 42.6 ($\pm 12$) | 20,448.2 ($\pm 15290.2$) | 64.9<br>($\pm 16.4$) | 53.2 |

**Table 2.** PGS with significant LabWAS analyses in AFR and EUR PMBB patients. PGSs with at least one significantly associated laboratory measure are shown. For each PGS, the table reports the sample size range across the laboratory measures, the number of labs that reach Bonferroni-corrected significance and FDR-Correct significance, and the laboratory measure with the strongest association.

| Table 2 |  |  |  |  |  |
| --- | --- | --- | --- | --- | --- |
| Ancestry | Phenotype/PGS | Sample<br>Size<br>Range | Significant Labs<br>(Bonferroni<br>$p < 0.05$) | Significant<br>Labs<br>(FDR $< 0.5$) | Most Significant Lab |
| AFR | BD | 107-10,473 | 4 | 4 | Neutrophils<br>(OR= 0.95, $p = 7.35 \times 10^{-7}$) |
| | PTSD | | 4 | 5 | WBC<br>(OR= 1.06, $p = 4.75 \times 10^{-6}$) |
| | TUD | | 2 | 2 | WBC (OR= 0.96, $p = 1.17 \times 10^{-5}$) |
| | CUD | | 1 | 1 | Neutrophils (OR= 0.96, $p = 5.73 \times 10^{-4}$) |
| EUR | BMI | 216-35,131 | 44 | 56 | BMI (OR= 1.39, $p \leq 2.20 \times 10^{-16}$) |
| | T2D | | 42 | 55 | Glucose (OR= 1.30, $p \leq 2.20 \times 10^{-16}$) |
| | HDL Cholesterol | | 12 | 21 | HDL (OR= 1.34, $p \leq 2.20 \times 10^{-16}$) |
| | LDL Cholesterol | | 7 | 9 | LDL (OR= 1.25, $p = 3.67 \times 10^{-213}$) |
| | Total Cholesterol | | 8 | 18 | Total Cholesterol<br>(OR= 1.24, $p = 5.05 \times 10^{-196}$) |
| | Total<br>Triglycerides | | 17 | 30 | Triglycerides (OR= 1.33, $p \leq 2.20 \times 10^{-16}$) |
| | CAD | | 8 | 6 | HDL (OR= 0.96, $p = 1.30 \times 10^{-11}$) |
|  | ALZ |  | 3 | 5 | LDL (OR= 1.06, p =<br>4.43 x 10<sup>-16</sup>) |
|  | ANX |  | 19 | 26 | BMI (OR= 1.04, p =<br>1.09 x 10<sup>-13</sup>) |
|  | ADHD |  | 28 | 40 | BMI (OR= 1.08, p =<br>9.13 x 10<sup>-49</sup>) |
|  | MDD |  | 17 | 22 | BMI (OR= 1.05, p =<br>8.6 x 10<sup>-21</sup>) |
|  | PTSD |  | 19 | 27 | BMI (OR= 1.07, p =<br>4.32 x 10<sup>-33</sup>) |
|  | AN |  | 11 | 22 | BMI (OR= 0.93, p =<br>2.46 x 10<sup>-29</sup>) |
|  | OCD |  | 2 | 2 | BMI (OR= 0.98, p =<br>4.05 x 10<sup>-6</sup>) |
|  | SCZ |  | 3 | 3 | Weight (OR= 0.97, p =<br>1.05 x 10<sup>-6</sup>) |
|  | BD |  | 4 | 14 | Creatine (OR= 0.98, p =<br>3.65 x 10<sup>-6</sup>) |
|  | TUD |  | 29 | 42 | BMI (OR= 1.05, p =<br>8.97 x 10<sup>-18</sup>) |
|  | EXT |  | 21 | 27 | BMI (OR= 1.05, p =<br>1.83 x 10<sup>-21</sup>) |
|  | PAU |  | 4 | 8 | RBC (OR= 0.97, p =<br>1.60 x 10<sup>-8</sup>) |
|  | AUD |  | 4 | 14 | RBC (OR= 0.96, p =<br>2.98 x 10<sup>-12</sup>) |
|  | AF |  | 7 | 20 | Hemoglobin (OR=<br>0.97, p = 1.64 x 10<sup>-8</sup>) |
|  | CUD |  | 3 | 3 | O2SAT (OR= 0.98, p =<br>1.13 x 10<sup>-4</sup>) |

**Figure 1.**
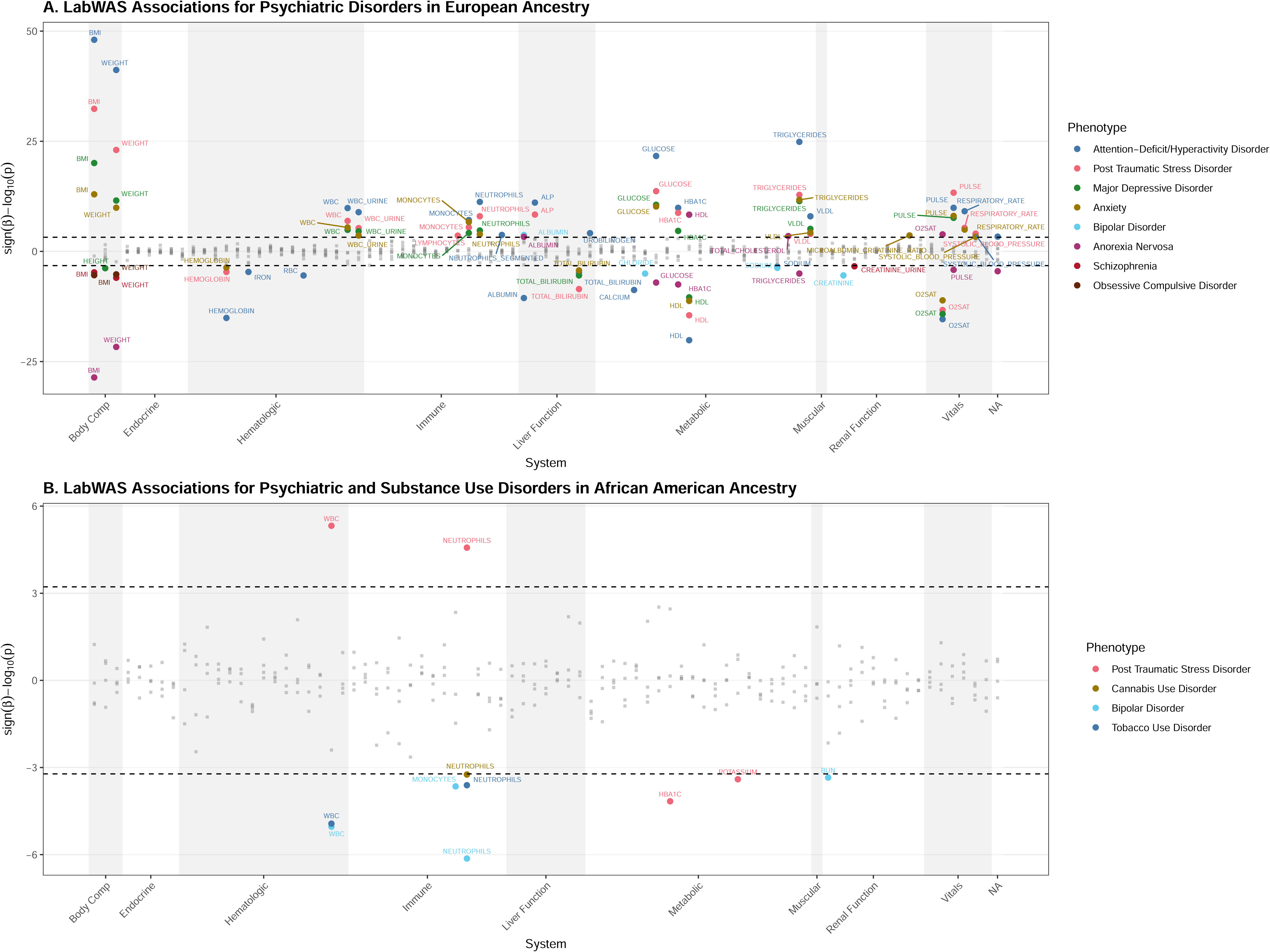
Ancestry-stratified overview of psychiatric PGS-laboratory associations. Shown are the association results for multiple psychiatric disorder polygenic risk scores with laboratory values in EUR (A) and AFR (B) ancestries. Each point represents a lab-PGS association, plotted according to effect size, statistical significance, and lab system. Dashed horizontal lines indicate the Bonferroni-corrected significance threshold.

**Figure 2.**
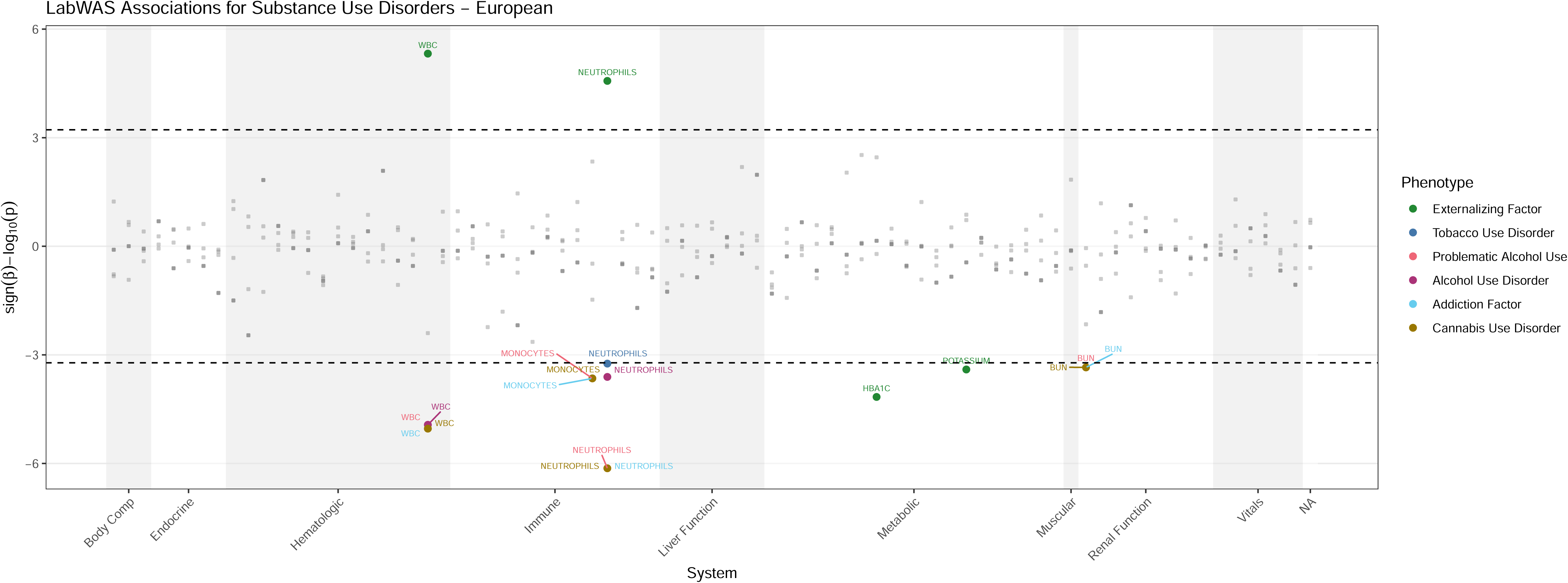
Summary of lab associations for substance use disorders polygenic risk in European ancestry. LabWAS results are shown for the substance use polygenic risk score in individuals of EUR ancestry. Each point represents the associations between the SUD PGS and lab value, ordered by lab system, and plotted based on effect size and statistical significance. Dashed horizontal lines indicate the Bonferroni-corrected significance threshold.

**Figure 3.**
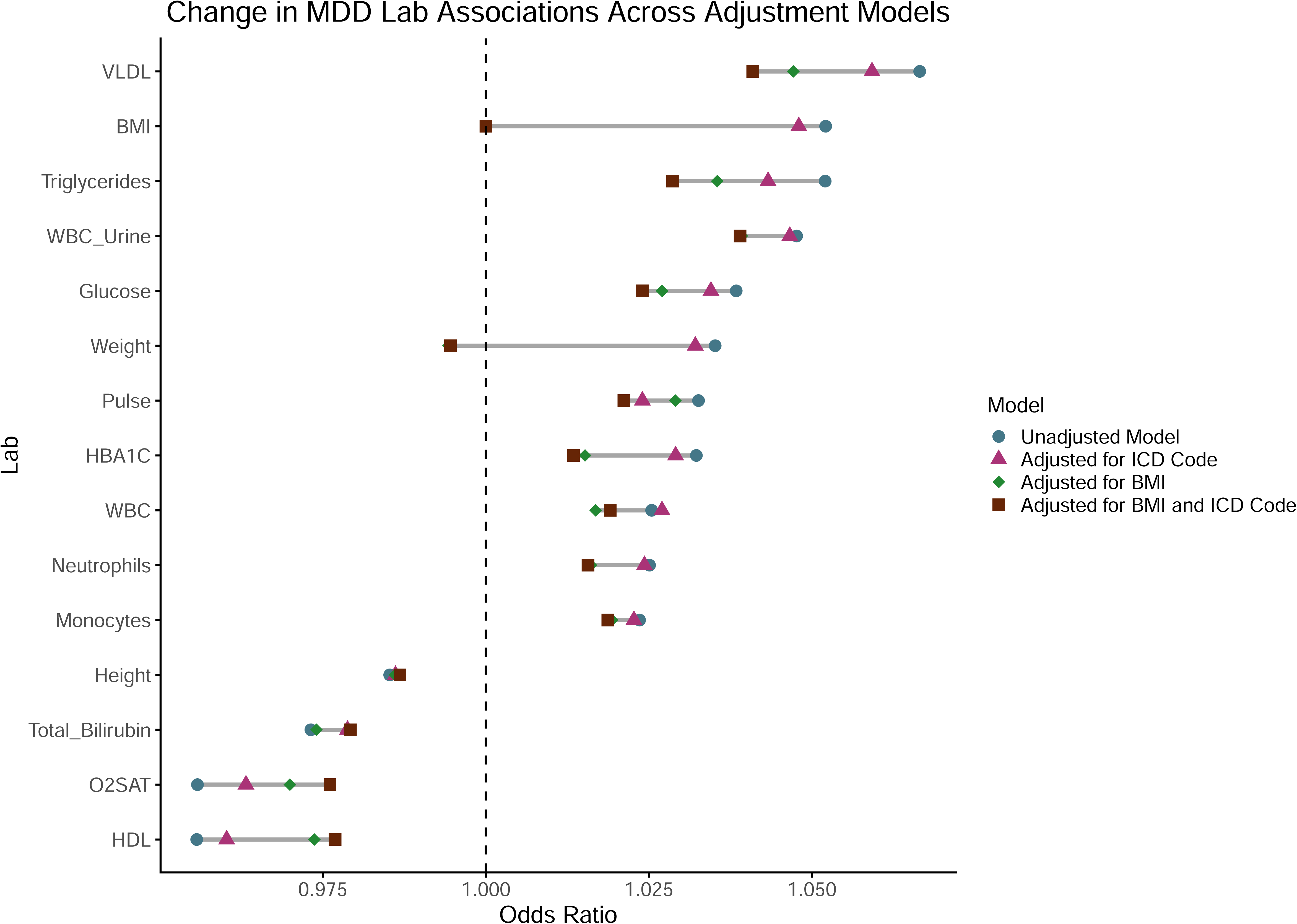
Changes in laboratory association effect sizes for MDD polygenic risk scores across adjustment models. Odds ratios (ORs) for associations between MDD polygenic risk scores and laboratory values were plotted using models with no additional adjustment, adjustment for MDD diagnosis, adjustment for BMI, and adjustment for both BMI and MDD diagnosis (ICD code present).

## Data Availability

The data used in this study are derived from electronic health records and cannot be made publicly available due to patient privacy and data use agreement restrictions. Analysis code are available upon reasonable request to the corresponding author.

## Acknowledgements

This work is supported by National Institutes of Health (Grant No. R01MH137212 to RLK and T32HG009495 to KLF). We acknowledge the Penn Medicine BioBank (PMBB) for providing data and thank the patient-participants of Penn Medicine who consented to participate in this research program. We would also like to thank the Penn Medicine BioBank team and Regeneron Genetics Center for providing genetic variant data for analysis. The PMBB is approved under IRB protocol# 813913 and supported by Perelman School of Medicine at University of Pennsylvania, a gift from the Smilow family, and the National Center for Advancing Translational Sciences of the National Institutes of Health under CTSA award number UL1TR0018.

## REFERENCES

1. Xu J jie, Ding L yu, Sun C cong, Qiao Y, Wang M ti, Zheng J xin, et al. The burden of mental disorders, substance use disorders, and self-harm among youths globally: findings from the 2021 Global Burden of Disease study. Transl Psychiatry. 2025 Sep 29;15(1):346. doi:10.1038/s41398-025-03533-x

2. Wu G, Dong Z, Li Z, Zhao Q, Chen S, Dong Q, et al. Comorbidity patterns and immune-metabolic differences in patients with acute-episode of schizophrenia spectrum disorders. Schizophrenia. 2025 Jul 19;11(1):102. doi:10.1038/s41537-025-00646-6

3. Finlay S, Rudd D, McDermott B, Sarnyai Z. Allostatic load and systemic comorbidities in psychiatric disorders. Psychoneuroendocrinology. 2022 Jun 1;140:105726. doi:10.1016/j.psyneuen.2022.105726

4. Polderman TJC, Benyamin B, de Leeuw CA, Sullivan PF, van Bochoven A, Visscher PM, et al. Meta-analysis of the heritability of human traits based on fifty years of twin studies. Nat Genet. 2015 Jul;47(7):702–9. doi:10.1038/ng.3285

5. Goldstein JA, Weinstock JS, Bastarache LA, Larach DB, Fritsche LG, Schmidt EM, et al. LabWAS: Novel findings and study design recommendations from a meta-analysis of clinical labs in two independent biobanks. PLoS Genet. 2020 Nov 11;16(11):e1009077. doi:10.1371/journal.pgen.1009077 PubMed PMID: 33175840; PubMed Central PMCID: PMC7682892.

6. Genome-wide trans-ancestry meta-analysis provides insight into the genetic architecture of type 2 diabetes susceptibility. Nat Genet. 2014 Mar;46(3):234–44. doi:10.1038/ng.2897 PubMed PMID: 24509480; PubMed Central PMCID: PMC3969612.

7. Sharma V, Sharma I, Sethi I, Mahajan A, Singh G, Angural A, et al. Replication of newly identified type 2 diabetes susceptible loci in Northwest Indian population. Diabetes Res Clin Pract. 2017 Apr 1;126:160–3. doi:10.1016/j.diabres.2017.02.013 PubMed PMID: 28258026.

8. Ustianowski P, Malinowski D, Czerewaty M, Safranow K, Tarnowski M, Dziedziejko V, et al. COBLL1 and IRS1 Gene Polymorphisms and Placental Expression in Women with Gestational Diabetes. Biomedicines. 2022 Aug 9;10(8):1933. doi:10.3390/biomedicines10081933 PubMed PMID: 36009479; PubMed Central PMCID: PMC9405993.

9. Badillo N, Khatib M, Kahar P, Khanna D. Correlation Between Body Mass Index and Depression/Depression-Like Symptoms Among Different Genders and Races. Cureus. 14(2):e21841. doi:10.7759/cureus.21841 PubMed PMID: 35291524; PubMed Central PMCID: PMC8896404.

10. Sforzini L, Cattaneo A, Ferrari C, Turner L, Mariani N, Enache D, et al. Higher immune-related gene expression in major depression is independent of CRP levels: results from the BIODEP study. Transl Psychiatry. 2023 Jun 1;13(1):185. doi:10.1038/s41398-023-02438-x

11. Min X, Wang G, Cui Y, Meng P, Hu X, Liu S, et al. Association between inflammatory cytokines and symptoms of major depressive disorder in adults. Front Immunol. 2023 Feb 13;14:1110775. doi:10.3389/fimmu.2023.1110775 PubMed PMID: 36860860; PubMed Central PMCID: PMC9968963.

12. Cortese S, Moreira-Maia CR, St Fleur D, Morcillo-Peñalver C, Rohde LA, Faraone SV. Association Between ADHD and Obesity: A Systematic Review and Meta-Analysis. Am J Psychiatry. 2016 Jan;173(1):34–43. doi:10.1176/appi.ajp.2015.15020266 PubMed PMID: 26315982.

13. Skorobogatov K, De Picker L, Wu CL, Foiselle M, Richard JR, Boukouaci W, et al. Immune-based Machine learning Prediction of Diagnosis and Illness State in Schizophrenia and Bipolar Disorder. Brain Behav Immun. 2024 Nov;122:422–32. doi:10.1016/j.bbi.2024.08.013 PubMed PMID: 39151650.

14. Girshkin L, Matheson SL, Shepherd AM, Green MJ. Morning cortisol levels in schizophrenia and bipolar disorder: A meta-analysis. Psychoneuroendocrinology. 2014 Nov 1;49:187–206. doi:10.1016/j.psyneuen.2014.07.013

15. Yin B, Cai Y, Teng T, Wang X, Liu X, Li X, et al. Identifying plasma metabolic characteristics of major depressive disorder, bipolar disorder, and schizophrenia in adolescents. Transl Psychiatry. 2024 Mar 26;14(1):163. doi:10.1038/s41398-024-02886-z PubMed PMID: 38531835; PubMed Central PMCID: PMC10966062.

16. Pendergrass SA, Brown-Gentry K, Dudek S, Frase A, Torstenson ES, Goodloe R, et al. Phenome-Wide Association Study (PheWAS) for Detection of Pleiotropy within the Population Architecture using Genomics and Epidemiology (PAGE) Network. PLoS Genet. 2013 Jan 31;9(1):e1003087. doi:10.1371/journal.pgen.1003087 PubMed PMID: 23382687; PubMed Central PMCID: PMC3561060.

17. Dennis JK, Sealock JM, Straub P, Lee YH, Hucks D, Actkins K, et al. Clinical laboratory test-wide association scan of polygenic scores identifies biomarkers of complex disease. Genome Med. 2021 Jan 13;13:6. doi:10.1186/s13073-020-00820-8 PubMed PMID: 33441150; PubMed Central PMCID: PMC7807864.

18. Ge T, Chen CY, Ni Y, Feng YCA, Smoller JW. Polygenic prediction via Bayesian regression and continuous shrinkage priors. Nat Commun. 2019 Apr 16;10:1776. doi:10.1038/s41467-019-09718-5 PubMed PMID: 30992449; PubMed Central PMCID: PMC6467998.

19. The 1000 Genomes Project: Welcome to a New World - PMC [Internet]. [cited 2026 Aug 6]. Available from: https://pmc.ncbi.nlm.nih.gov/articles/PMC4690620/

20. Xia W, Yu H, Wang G. Coronary Artery Disease with Elevated Levels of HDL Cholesterol Is Associated with Distinct Lipid Signatures. Metabolites. 2023 May 26;13(6):695. doi:10.3390/metabo13060695 PubMed PMID: 37367853; PubMed Central PMCID: PMC10301469.

21. Toth PP. High-Density Lipoprotein and Cardiovascular Risk. Circulation. 2004 Apr 20;109(15):1809–12. doi:10.1161/01.CIR.0000126889.97626.B8

22. Grotzinger AD, Werme J, Peyrot WJ, Frei O, de Leeuw C, Bicks LK, et al. Mapping the genetic landscape across 14 psychiatric disorders. Nature. 2026;649(8096):406–15. doi:10.1038/s41586-025-09820-3 PubMed PMID: 41372416; PubMed Central PMCID: PMC12779569.

23. Zhang M, Chen J, Yin Z, Wang L, Peng L. The association between depression and metabolic syndrome and its components: a bidirectional two-sample Mendelian randomization study. Transl Psychiatry. 2021 Dec 13;11:633. doi:10.1038/s41398-021-01759-z PubMed PMID: 34903730; PubMed Central PMCID: PMC8668963.

24. Fang Y, Sen S, Pathak GA, Burmeister M, Richmond-Rakerd LS. Testing bidirectional associations of major depressive disorder with medical conditions: two-sample Mendelian randomization study. Npj Ment Health Res. 2026 Apr 9;5(1):24. doi:10.1038/s44184-026-00204-7

25. van de Wouw M, Sichetti M, Long-Smith CM, Ritz NL, Moloney GM, Cusack AM, et al. Acute stress increases monocyte levels and modulates receptor expression in healthy females. Brain Behav Immun. 2021 May 1;94:463–8. doi:10.1016/j.bbi.2021.03.005

26. Škarpa I, Rubeša G, Moro L, Manestar D. Changes of Cytolytic Cells and Perforin Expression in Patients with Posttraumatic Stress Disorder.

27. Islam MR, Sohan M, Daria S, Masud AA, Ahmed MU, Roy A, et al. Evaluation of inflammatory cytokines in drug-naïve major depressive disorder: A systematic review and meta-analysis. Int J Immunopathol Pharmacol. 2023 Aug 25;37:03946320231198828. doi:10.1177/03946320231198828 PubMed PMID: 37625799; PubMed Central PMCID: PMC10467201.

28. Shah P, Iwata Y, Caravaggio F, Plitman E, Brown EE, Kim J, et al. Alterations in body mass index and waist-to-hip ratio in never and minimally treated patients with psychosis: A systematic review and meta-analysis. Schizophr Res. 2019 Jun 1;208:420–9. doi:10.1016/j.schres.2019.01.005

29. Rigamonti AE, Paroni R, Sadikovic A, Morano C, Bondesan A, Caroli D, et al. Evaluation of the Sphingolipidomic Profile in Women with Anorexia Nervosa: Relationships with Parameters Related to Body Composition, Cardiovascular Function, Glucometabolic Homeostasis, and Lipoprotein Metabolism. J Clin Med. 2025 Sep 15;14(18):6482. doi:10.3390/jcm14186482 PubMed PMID: 41010687; PubMed Central PMCID: PMC12470723.

30. Hussain AA, Hübel C, Hindborg M, Lindkvist E, Kastrup AM, Yilmaz Z, et al. Increased lipid and lipoprotein concentrations in anorexia nervosa: a systematic review and meta-analysis. Int J Eat Disord. 2019 Jun;52(6):611–29. doi:10.1002/eat.23051 PubMed PMID: 30920679; PubMed Central PMCID: PMC6842568.

31. Hussain AA, Carlsson J, Mortensen EL, Hemmingsen SD, Bulik CM, Støving RK, et al. A naturalistic study of plasma lipid alterations in female patients with anorexia nervosa before and after weight restoration treatment. J Eat Disord. 2024 Jul 2;12(1):92. doi:10.1186/s40337-024-01055-3

32. Affaticati LM, Buoli M, Vaccaro N, Manzo F, Scalia A, Coloccini S, et al. The Impact of Clinical Factors, Vitamin B12 and Total Cholesterol on Severity of Anorexia Nervosa: A Multicentric Cross-Sectional Study. Nutrients. 2023 Jan;15(23):4954. doi:10.3390/nu15234954

33. Watson HJ, Yilmaz Z, Thornton LM, Hübel C, Coleman JR, Gaspar HA, et al. Genome-wide Association Study Identifies Eight Risk Loci and Implicates Metabo-Psychiatric Origins for Anorexia Nervosa. Nat Genet. 2019 Aug;51(8):1207–14. doi:10.1038/s41588-019-0439-2 PubMed PMID: 31308545; PubMed Central PMCID: PMC6779477.

34. Huh JH, Choi SI, Lim JS, Chung CH, Shin JY, Lee MY. Lower Serum Creatinine Is Associated with Low Bone Mineral Density in Subjects without Overt Nephropathy. PLoS ONE. 2015 Jul 24;10(7):e0133062. doi:10.1371/journal.pone.0133062 PubMed PMID: 26207750; PubMed Central PMCID: PMC4514793.

35. Proctor DN, O’Brien PC, Atkinson EJ, Nair KS. Comparison of techniques to estimate total body skeletal muscle mass in people of different age groups. Am J Physiol. 1999 Sep;277(3):E489–495. doi:10.1152/ajpendo.1999.277.3.E489 PubMed PMID: 10484361.

36. Marthoenis M, Martina M, Alfiandi R, Dahniar D, Asnurianti R, Sari H, et al. Investigating Body Mass Index and Body Composition in Patients with Schizophrenia: A Case-Control Study. Schizophr Res Treat. 2022 Feb 27;2022:1381542. doi:10.1155/2022/1381542 PubMed PMID: 35265376; PubMed Central PMCID: PMC8898882.

37. Nygård M, Brobakken MF, Roel RB, Taylor JL, Reitan SK, Güzey IC, et al. Patients with schizophrenia have impaired muscle force-generating capacity and functional performance. Scand J Med Sci Sports. 2019 Dec;29(12):1968–79. doi:10.1111/sms.13526 PubMed PMID: 31359490.

38. Bigdeli TB, Chatzinakos C, Bendl J, Barr PB, Venkatesh S, Gorman BR, et al. Biological insights into schizophrenia from ancestrally diverse populations. Nature. 2026 Mar;651(8105):404–13. doi:10.1038/s41586-025-10000-6

39. Fabrazzo M, Fuschillo A, Perris F, Catapano F. The unmasking of hidden severe hyponatremia after long-term combination therapy in exacerbated bipolar patients: a case series. Int Clin Psychopharmacol. 2019 Jul;34(4):206–10. doi:10.1097/YIC.0000000000000265 PubMed PMID: 30998596.

40. Jan WC, Yang SY, Chuang LC, Lu RB, Lu MK, Sun HS, et al. Exploring the associations between genetic variants in genes encoding for subunits of calcium channel and subtypes of bipolar disorder. J Affect Disord. 2014 Mar;157:80–6. doi:10.1016/j.jad.2013.12.044 PubMed PMID: 24581832.

41. Mullins N, Forstner AJ, O’Connell KS, Coombes B, Coleman JRI, Qiao Z, et al. Genome-wide association study of over 40,000 bipolar disorder cases provides new insights into the underlying biology. Nat Genet. 2021 Jun;53(6):817–29. doi:10.1038/s41588-021-00857-4 PubMed PMID: 34002096; PubMed Central PMCID: PMC8192451.

42. Yang L, Wheeler MA. Substance abuse and inflammation: consequences beyond the brain. Trends Immunol. 2026 May 25. doi:10.1016/j.it.2026.04.011

43. Ballard HS. The Hematological Complications of Alcoholism. Alcohol Health Res World. 1997;21(1):42–52. PubMed PMID: 15706762; PubMed Central PMCID: PMC6826798.

