## Supplementary Document for "A clinical laboratory test-wide association scan of polygenic scores identifies biomarkers of psychiatric disorders"

**Supplemental Methods**

**Phasing and Imputation**

Pre-imputation quality control was performed using PLINK software, removing variants with marker call rate < 95% and sample call rate < 90%. Samples for whom genetic sex and reported sex differed and could not be resolved were also excluded. Following quality control, datasets were divided into subsets by chromosome for imputation.

Phasing and imputation for the datasets were performed on the TOPMed Imputation server, with the phasing done using EAGLE v2.4 and imputation done using MINIMAC v4.1.6 software, with the reference panel being the GRCh38 reference genome. Due to the large size of the imputed data, it was chunked into ~1,000 equal sized files across the genome and then merged using bcftools.

Post-imputation filtering retained variants meeting at least one of the following criteria: the average imputation quality score (R^2^) > 0.2, or direct observation in either freezes sequencing data. Chromosome X variants were processed separately by splitting psuedoautosomal regions (PARs) prior to filtering and then merged after.

**Ancestry Estimation**

SNPs were first intersected between the HapMap3 reference panel and the GxS pVCF and filtered to common high-quality variants, then merged GxS pVCF for the HapMap3 dataset. Principal components (PCs) were calculated for HapMap3 samples and all non-HapMap3 samples onto those PCs. A kernel density estimation was trained for PC1 vs PC2 and PC3 vs PC4 for each ancestral group with the PCs of the HapMap3 samples each with their own bandwidth values. For each sample, the PCs were compared to their respective KDE for each HM2 population to make a normalized likelihood vector. Selected ancestral superclasses for each sample were concluded based on the reference populations identified.

Principal component analysis (PCA) to capture population structure were generated by using a linkage disequilibrium-pruned set of common variants. The dataset was merged with reference samples from the 1000 Genomes Project. PCA was conducted using the smartpca module of EIGENSOFT.

**Penn Medicine BioBank Banner Author List and Contribution Statements**

PMBB Leadership Team

Daniel J. Rader, M.D., Marylyn D. Ritchie, Ph.D.

**Contribution**: All authors contributed to securing funding, study design and oversight. All authors reviewed the final version of the manuscript.

Patient Recruitment and Regulatory Oversight

JoEllen Weaver, Nawar Naseer, Ph.D., M.P.H., Giorgio Sirugo, M.D., P.h.D., Afiya Poindexter, Yi-An Ko, Ph.D., Kyle P. Nerz

**Contributions**: JW manages patient recruitment and regulatory oversight of study. NN manages participant engagement, assists with regulatory oversight, and researcher access. GS assists with researcher access. AP, YK, KPN perform recruitment and enrollment of study participants.

Lab Operations

JoEllen Weaver, Meghan Livingstone, Fred Vadivieso, Stephanie DerOhannessian, Teo Tran, Julia Stephanowski, Salma Santos, Ned Haubein, P.h.D., Joseph Dunn

**Contribution**: JW, ML, FV, SD conduct oversight of lab operations. ML, FV, AK, SD, TT, JS, SS perform sample processing. NH, JD are responsible for sample tracking and the laboratory information management system.

Clinical Informatics

Anurag Verma, Ph.D., Colleen Morse Kripke, M.S. DPT, MSA, Marjorie Risman, M.S., Renae Judy, B.S., Colin Wollack, M.S.

**Contribution**: All authors contributed to the development and validation of clinical phenotypes used to identify study subjects and (when applicable) controls.

Genome Informatics

Anurag Verma Ph.D., Shefali S. Verma, Ph.D., Scott Damrauer, M.D., Yuki Bradford, M.S., Scott Dudek, M.S., Theodore Drivas, M.D., Ph.D.,

**Contribution**: AV, SSV, and SD are responsible for the analysis, design, and infrastructure needed to quality control genotype and exome data. YB performs the analysis. TD and AV provides variant and gene annotations and their functional interpretation of variants.

For PMBB, please use:

For Regeneron, please use:
