## Supplementary Figures for "A clinical laboratory test-wide association scan of polygenic scores identifies biomarkers of psychiatric disorders"

### Body Mass Index − European LabWAS

Figure S1

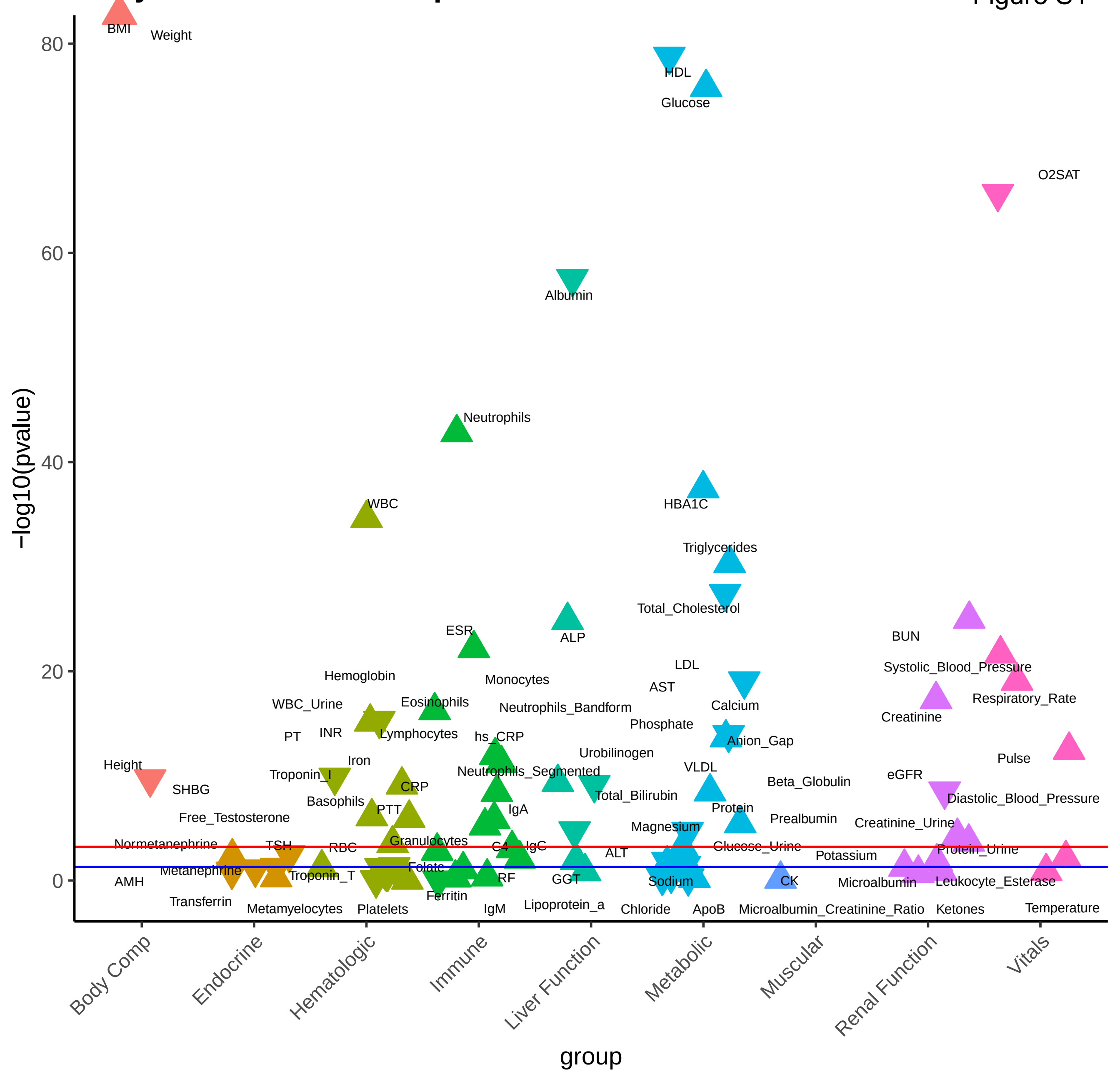

**Type 2 Diabetes − European LabWAS**

Figure S2

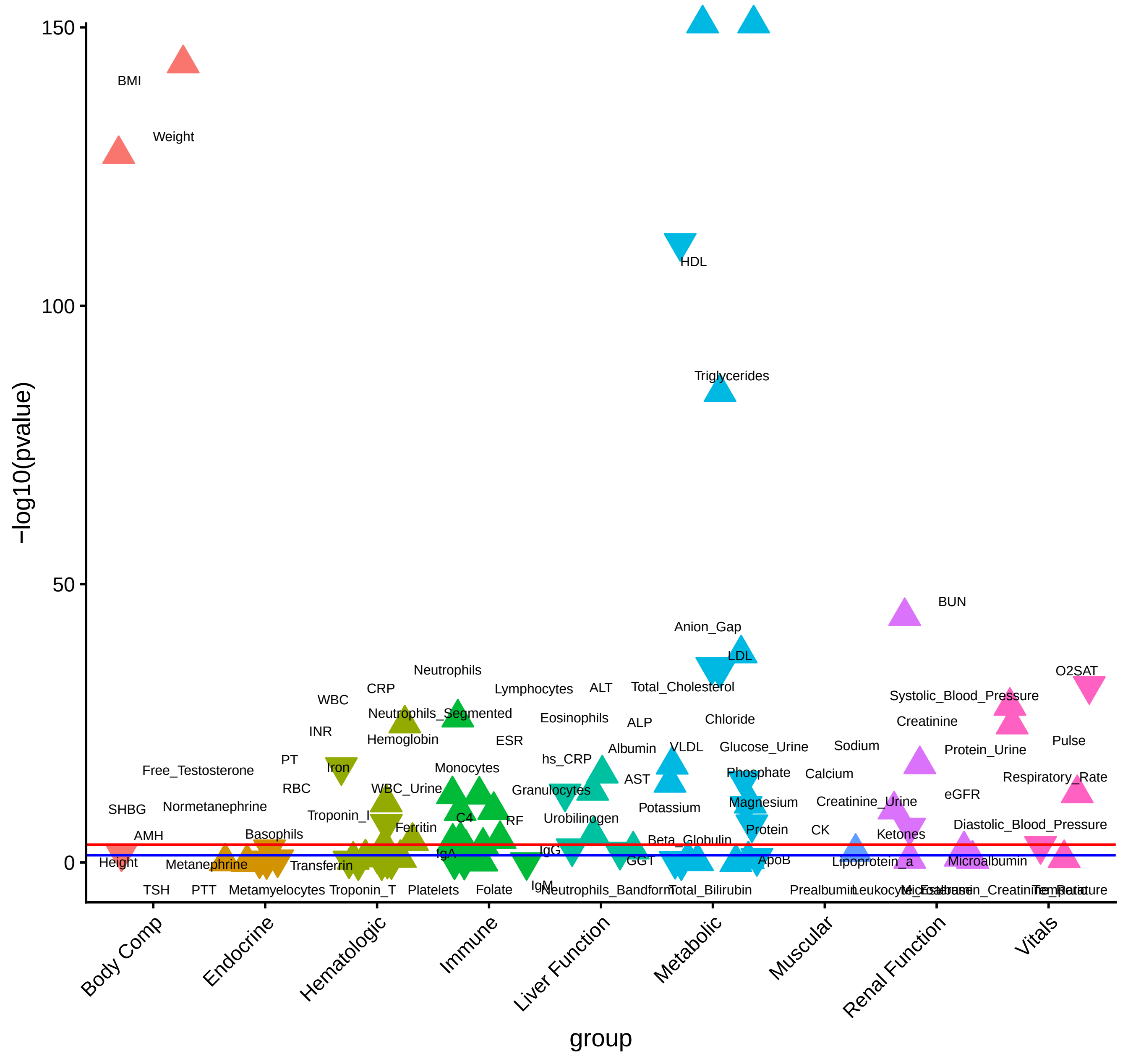

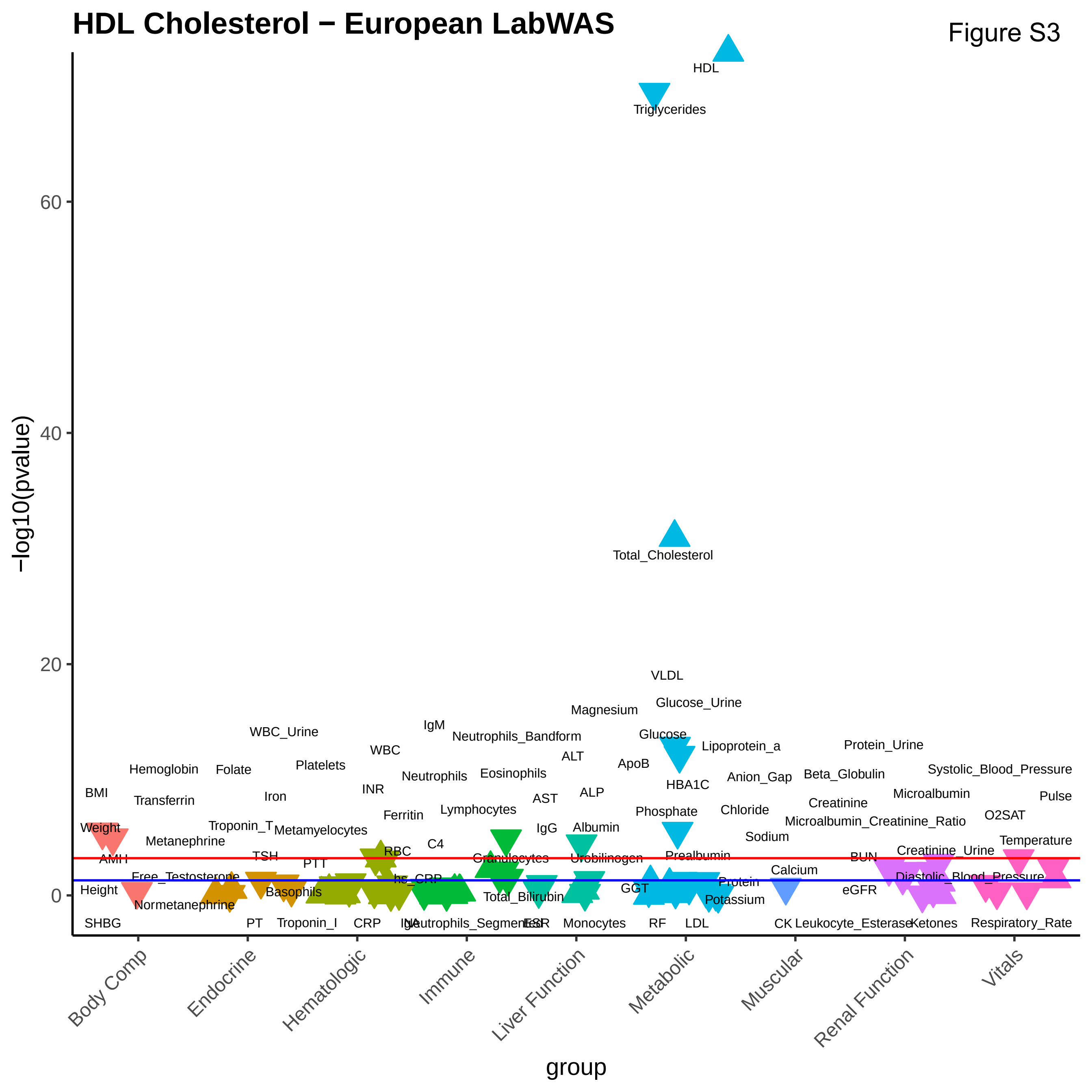

HDL Cholesterol − European LabWAS
Figure S3
−log10(pvalue)
group
Body Comp
Endocrine
Hematologic
Immune
Liver Function
Metabolic
Muscular
Renal Function
Vitals
HDL
Triglycerides
Total_Cholesterol
VLDL
Glucose_Urine
Magnesium
Glucose
Lipoprotein_a
ApoB
HBA1C
Protein_Urine
Systolic_Blood_Pressure
Pulse

Figure S4

#### LDL Cholesterol − European LabWAS

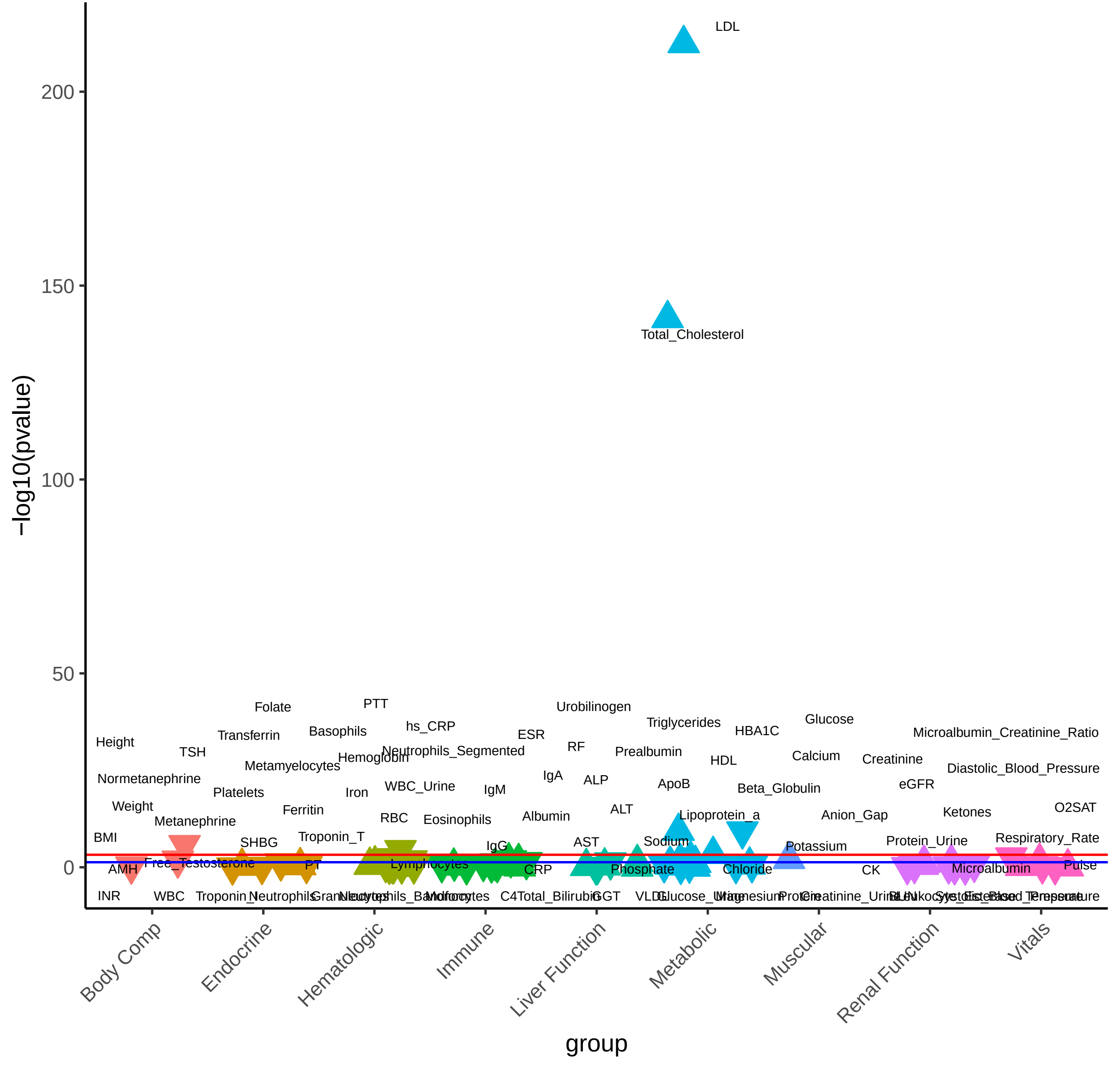

### Total Cholesterol − European LabWAS

Figure S5

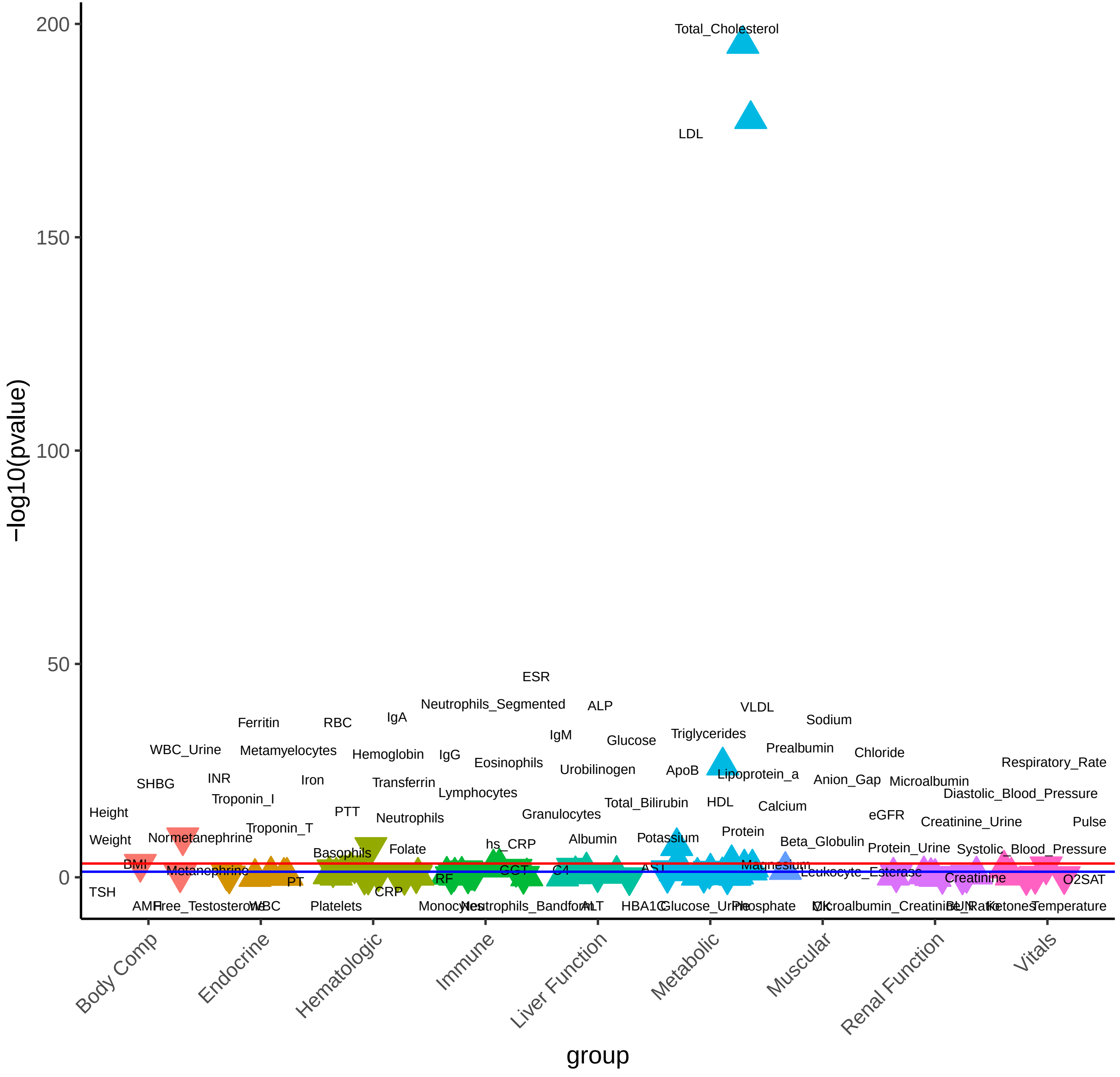

**Total Triglycerides − European LabWAS**

Figure S6

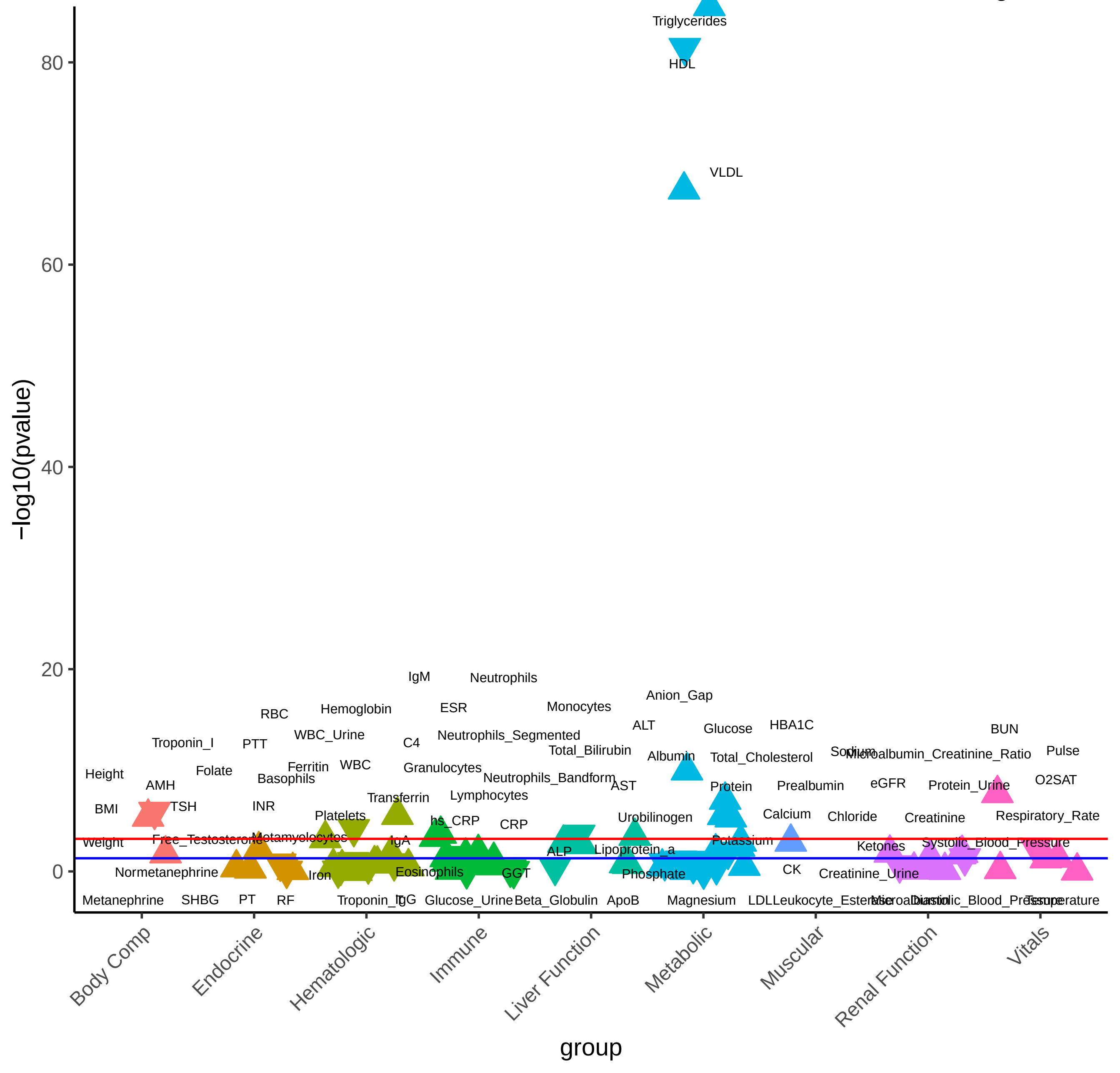

### Coronary Artery Disease − European LabWAS

Figure S7

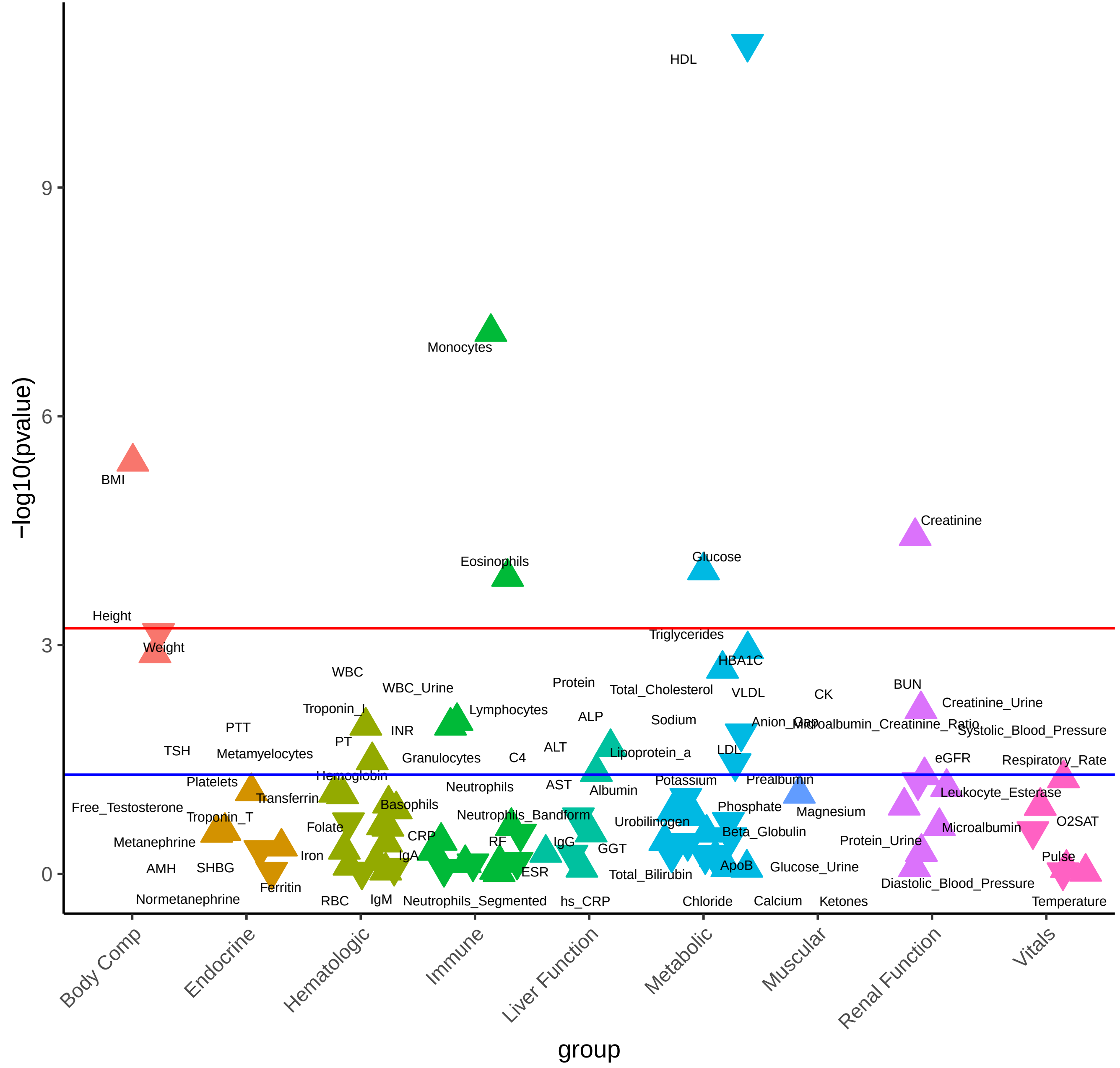

Figure S8

**Alzheimer's − European LabWAS**

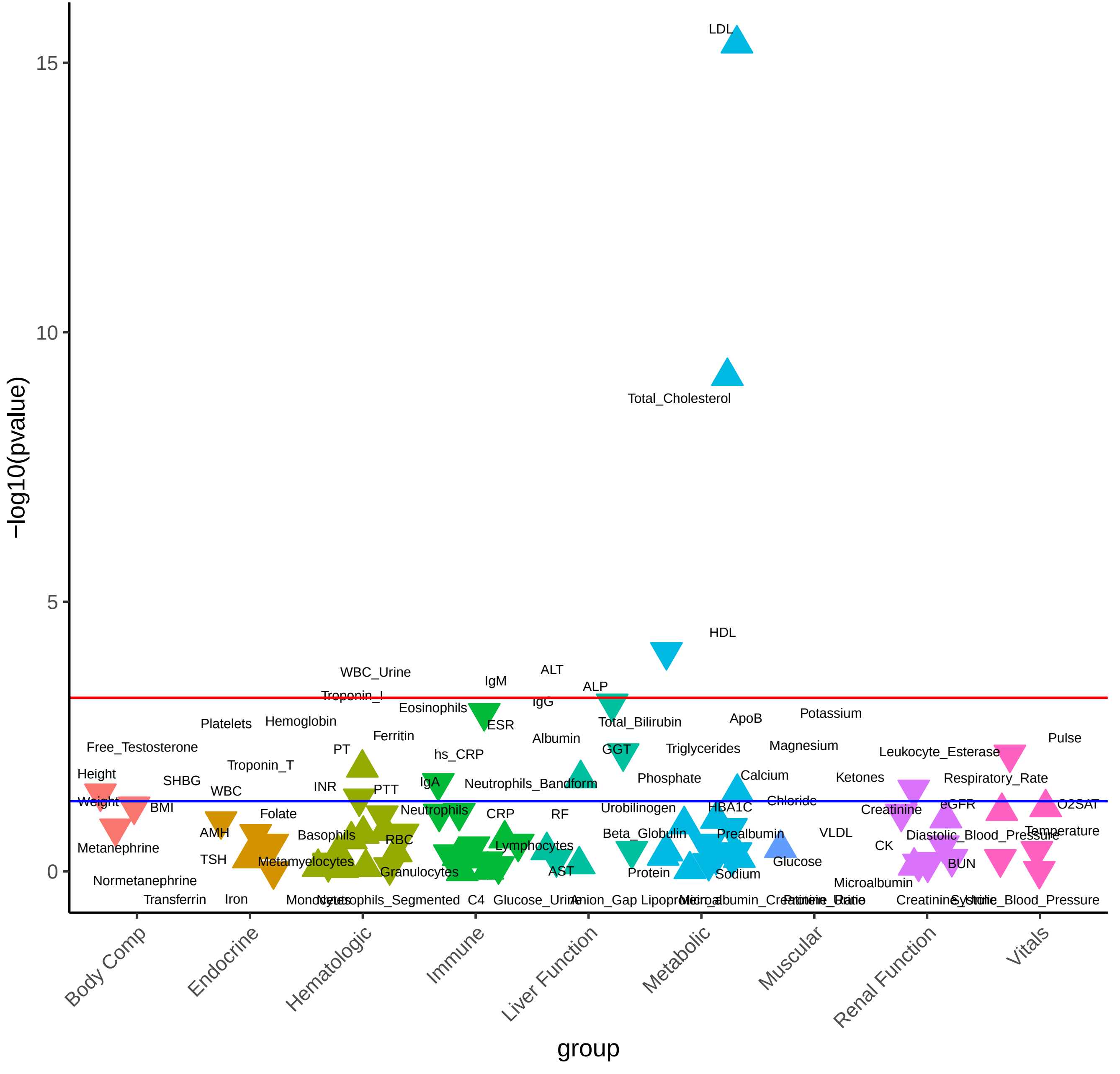

Figure S9

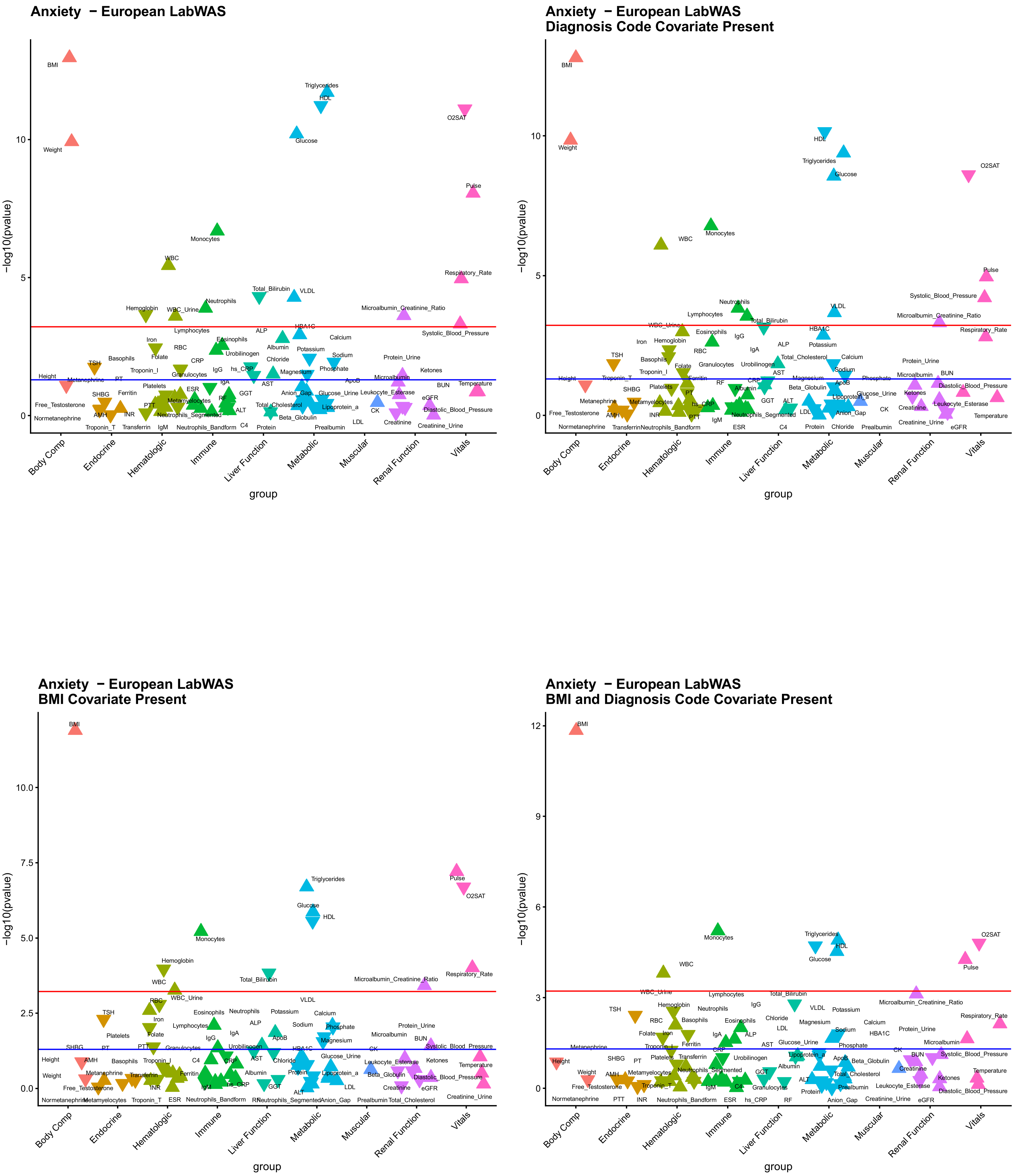
Anxiety − European LabWAS
Anxiety − European LabWAS
Diagnosis Code Covariate Present
Anxiety − European LabWAS
BMI Covariate Present
Anxiety − European LabWAS
BMI and Diagnosis Code Covariate Present
−log10(pvalue)
0
5
10
2.5
7.5
10.0
3
6
9
12
Body Comp
Endocrine
Hematologic
Immune
Liver Function
Metabolic
Muscular
Renal Function
Vitals
group

Figure S10

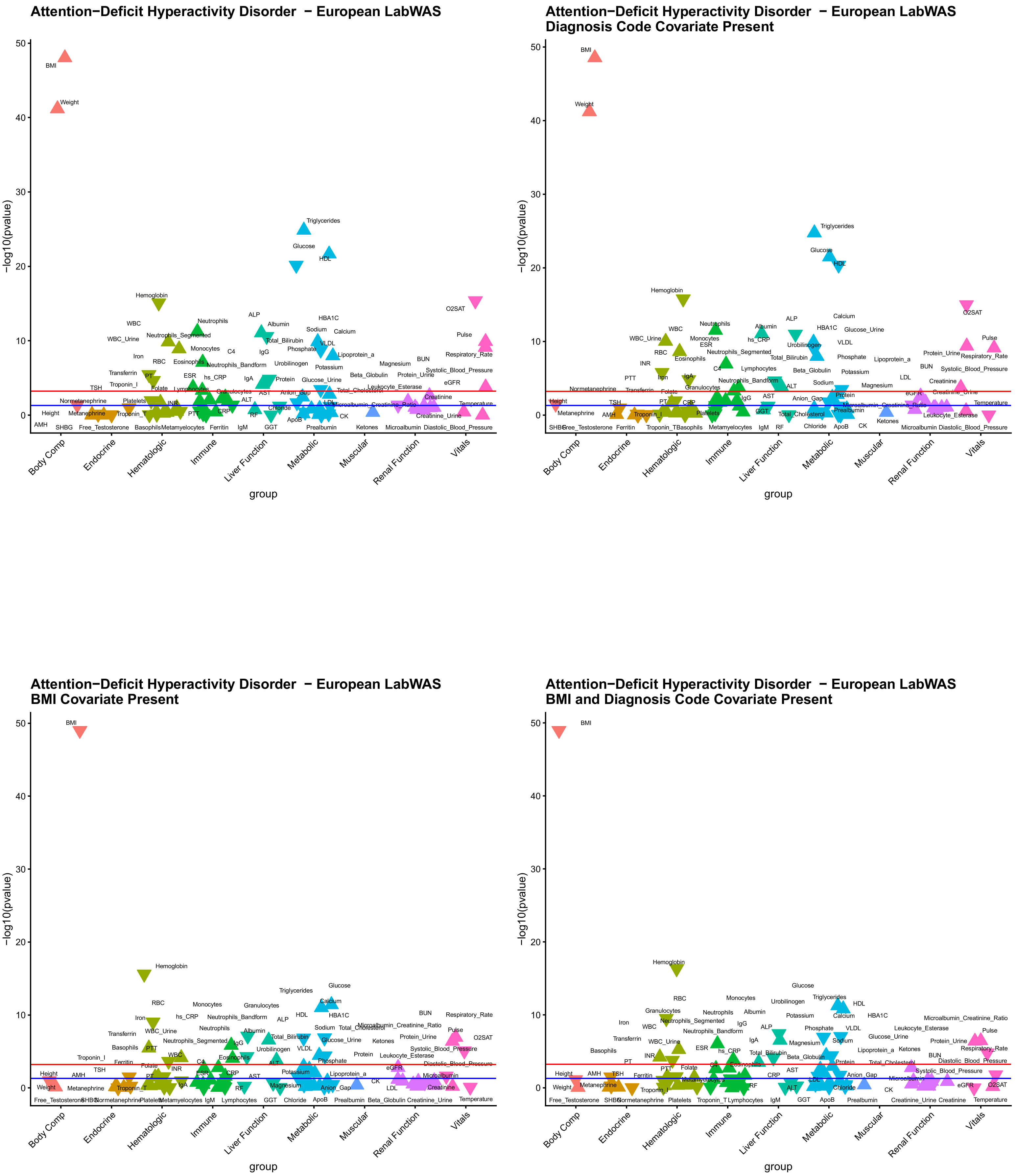

Attention−Deficit Hyperactivity Disorder − European LabWAS
Attention−Deficit Hyperactivity Disorder − European LabWAS
Diagnosis Code Covariate Present
Attention−Deficit Hyperactivity Disorder − European LabWAS
BMI Covariate Present
Attention−Deficit Hyperactivity Disorder − European LabWAS
BMI and Diagnosis Code Covariate Present
−log10(pvalue)
group
Body Comp
Endocrine
Hematologic
Immune
Liver Function
Metabolic
Muscular
Renal Function
Vitals

Figure S11

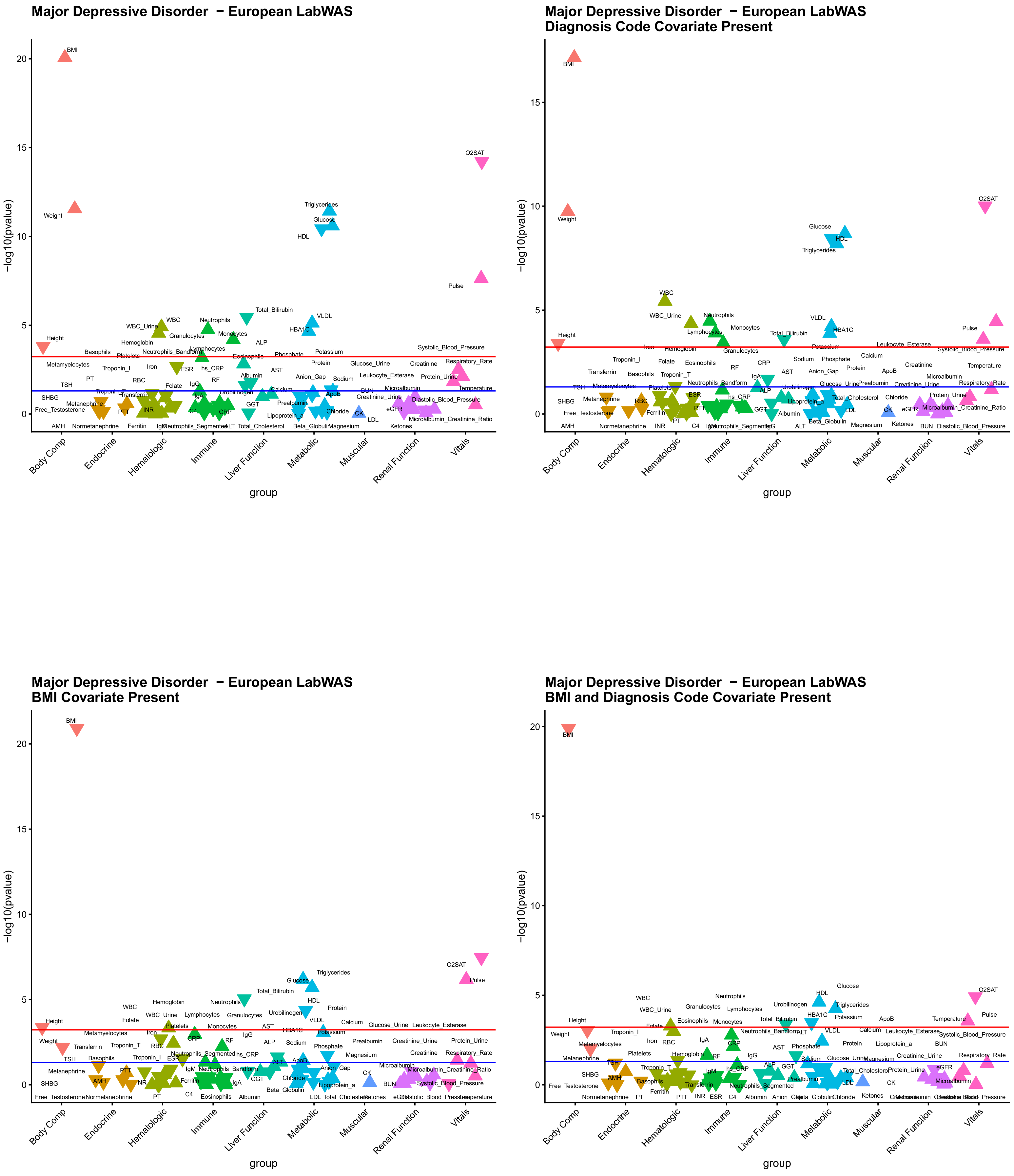
Major Depressive Disorder − European LabWAS
Major Depressive Disorder − European LabWAS
Diagnosis Code Covariate Present
Major Depressive Disorder − European LabWAS
BMI Covariate Present
Major Depressive Disorder − European LabWAS
BMI and Diagnosis Code Covariate Present
−log10(pvalue)
group
Body Comp
Endocrine
Hematologic
Immune
Liver Function
Metabolic
Muscular
Renal Function
Vitals
BMI
Weight
Height
O2SAT
Pulse
Triglycerides
Glucose
HDL
HBA1C
VLDL
WBC
Neutrophils

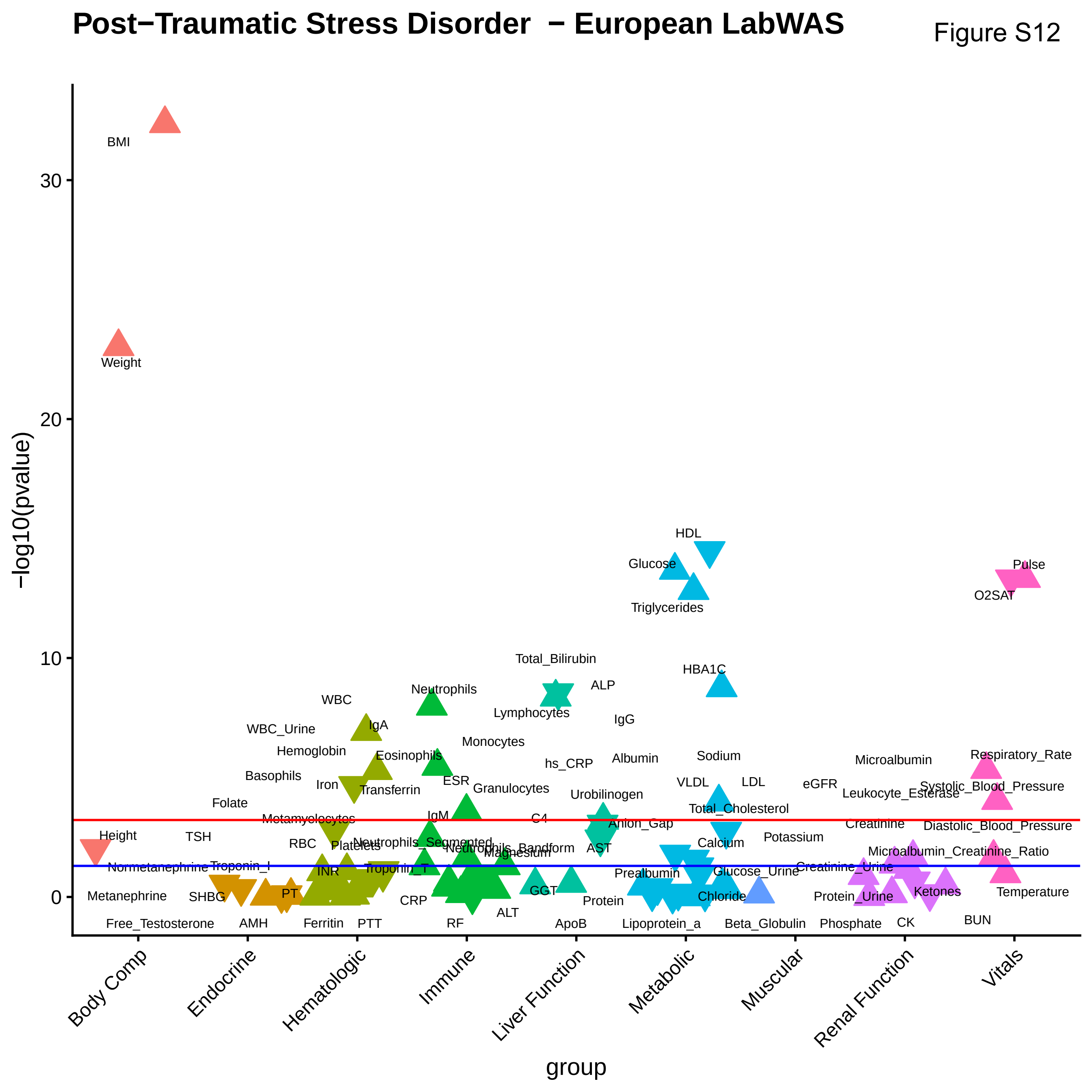
Post−Traumatic Stress Disorder − European LabWAS
Figure S12
−log10(pvalue)
group
Body Comp
Endocrine
Hematologic
Immune
Liver Function
Metabolic
Muscular
Renal Function
Vitals
0
10
20
30
BMI
Weight
Height
HDL
Glucose
Triglycerides
Pulse
O2SAT
Total_Bilirubin
HBA1C
ALP
Neutrophils
WBC
Lymphocytes
IgG
IgA
WBC_Urine
Monocytes
Hemoglobin
Eosinophils
Sodium
Albumin
Microalbumin
Respiratory_Rate
hs_CRP
Basophils
Iron
ESR
Transferrin
Granulocytes
VLDL
LDL
eGFR
Systolic_Blood_Pressure
Leukocyte_Esterase
Urobilinogen
Folate
Total_Cholesterol
Metamyelocytes
IgM
C4
Anion_Gap
Creatinine
Diastolic_Blood_Pressure
TSH
Potassium
RBC
Platelets
Neutrophils_Segmented
Neutrophils_Bandform
Magnesium
AST
Calcium
Microalbumin_Creatinine_Ratio
Normetanephrine
Troponin_I
Troponin_T
INR
Prealbumin
Glucose_Urine
Creatinine_Urine
PT
GGT
Metanephrine
SHBG
CRP
Protein
Chloride
Protein_Urine
Ketones
Temperature
ALT
BUN
Free_Testosterone
AMH
Ferritin
PTT
RF
ApoB
Lipoprotein_a
Beta_Globulin
Phosphate
CK

Figure S13

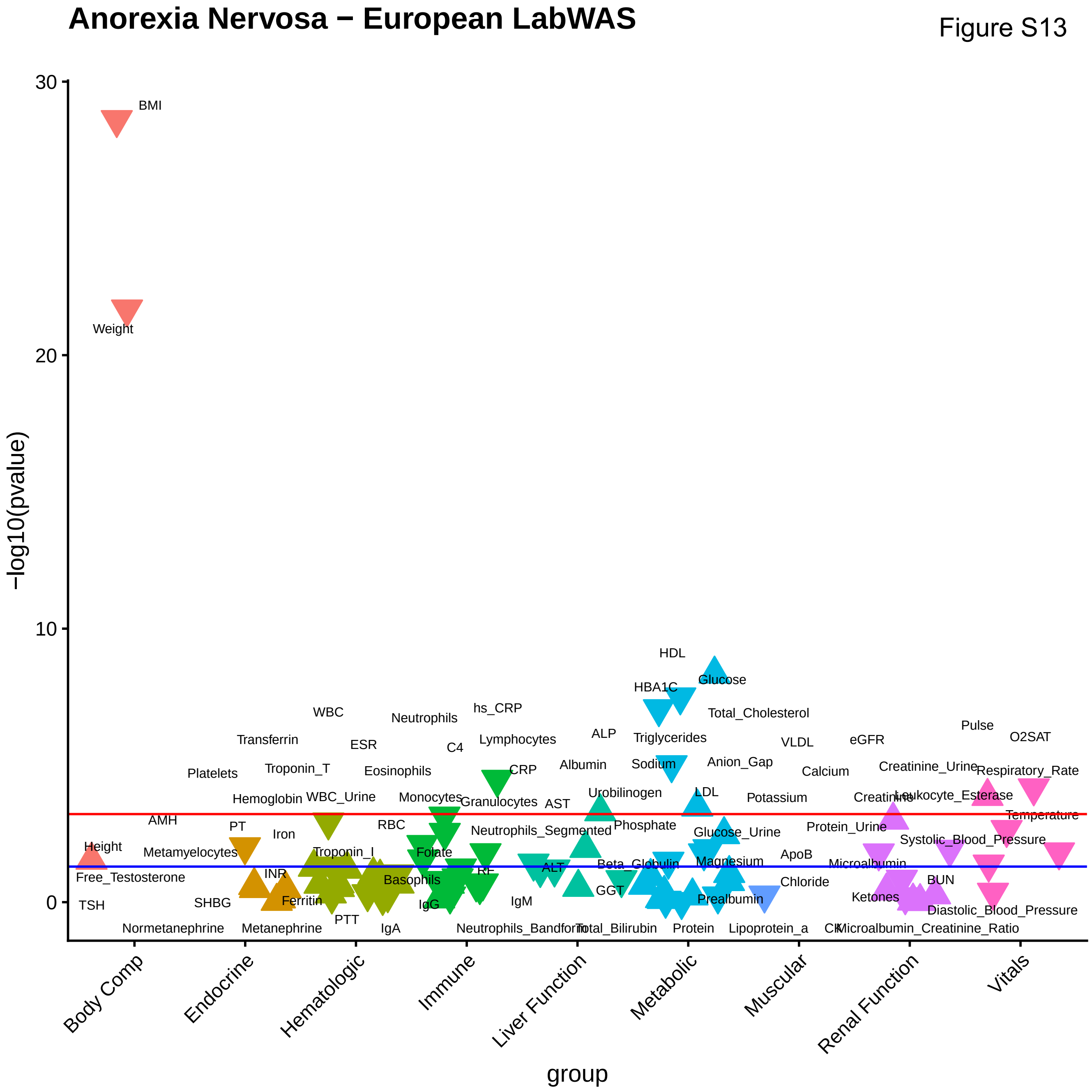
Anorexia Nervosa − European LabWAS
−log10(pvalue)
30
20
10
0
group
Body Comp
Endocrine
Hematologic
Immune
Liver Function
Metabolic
Muscular
Renal Function
Vitals
BMI
Weight
Height
Free_Testosterone
TSH
Normetanephrine
AMH
Metamyelocytes
SHBG
Platelets
Transferrin
Hemoglobin
PT
Iron
INR
Ferritin
Metanephrine
WBC
Troponin_T
WBC_Urine
Troponin_I
PTT
ESR
Eosinophils
RBC
Basophils
IgA
Neutrophils
Monocytes
Folate
IgG
C4
hs_CRP
Lymphocytes
CRP
Granulocytes
Neutrophils_Segmented
RF
IgM
Neutrophils_Bandform
AST
ALT
ALP
Albumin
Urobilinogen
GGT
Total_Bilirubin
HDL
HBA1C
Glucose
Total_Cholesterol
Triglycerides
Sodium
Anion_Gap
LDL
Phosphate
Glucose_Urine
Beta_Globulin
Magnesium
Prealbumin
Protein
Lipoprotein_a
VLDL
Calcium
Potassium
ApoB
Chloride
CK
eGFR
Creatinine_Urine
Creatinine
Protein_Urine
Microalbumin
Ketones
Microalbumin_Creatinine_Ratio
BUN
Leukocyte_Esterase
Systolic_Blood_Pressure
Diastolic_Blood_Pressure
Pulse
O2SAT
Respiratory_Rate
Temperature

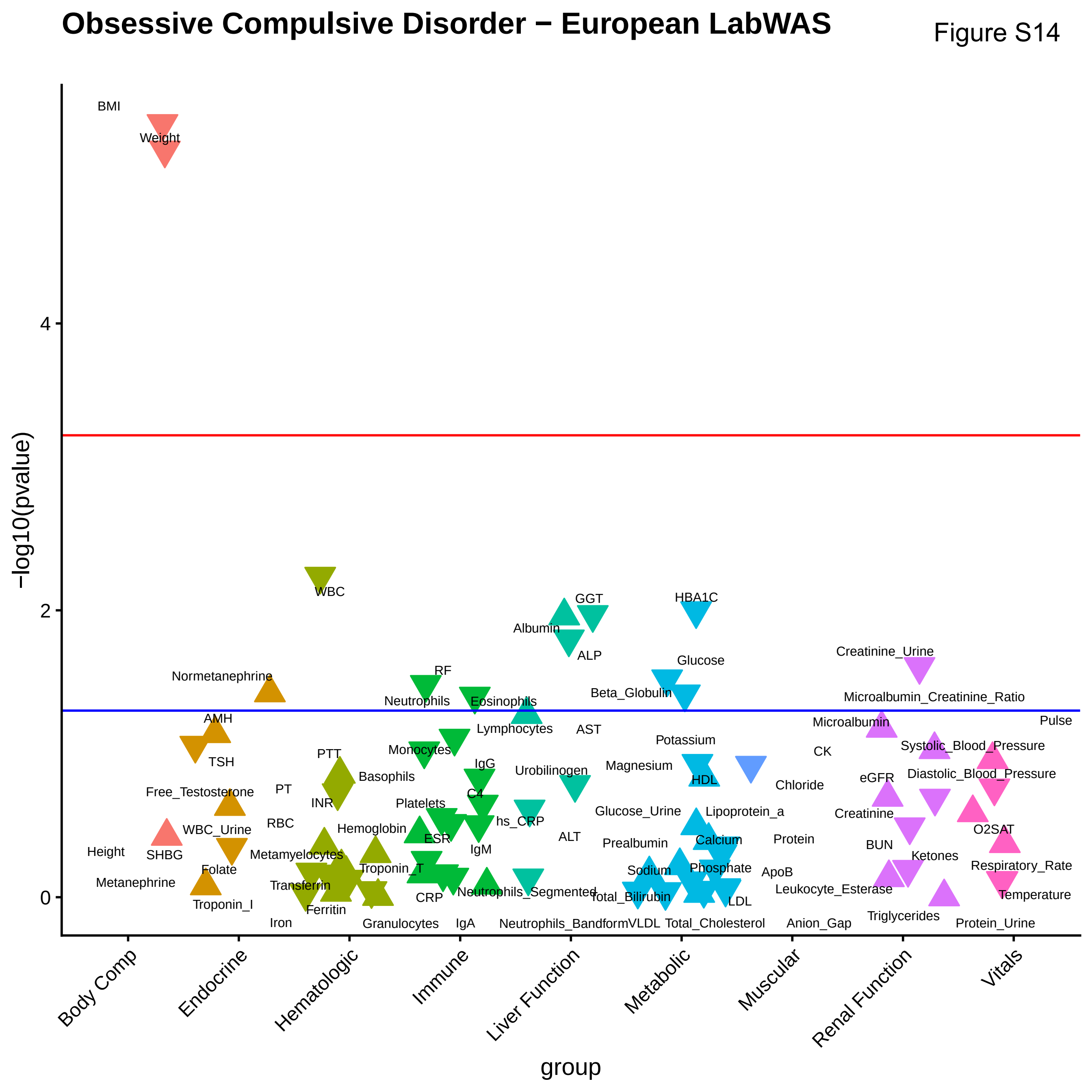

Obsessive Compulsive Disorder − European LabWAS
Figure S14
−log10(pvalue)
group
Body Comp
Endocrine
Hematologic
Immune
Liver Function
Metabolic
Muscular
Renal Function
Vitals
0
2
4
BMI
Weight
Height
Normetanephrine
AMH
TSH
Free_Testosterone
WBC_Urine
SHBG
Folate
Metanephrine
Troponin_I
WBC
PTT
PT
INR
RBC
Basophils
Hemoglobin
Metamyelocytes
Transferrin
Troponin_T
Ferritin
Iron
Granulocytes
RF
Neutrophils
Eosinophils
Lymphocytes
Monocytes
IgG
C4
Platelets
ESR
hs_CRP
IgM
CRP
IgA
Neutrophils_Segmented
GGT
Albumin
ALP
AST
Urobilinogen
ALT
Neutrophils_Bandform
HBA1C
Glucose
Beta_Globulin
Potassium
Magnesium
HDL
Glucose_Urine
Lipoprotein_a
Prealbumin
Calcium
Sodium
Phosphate
Total_Bilirubin
LDL
VLDL
Total_Cholesterol
CK
Chloride
Protein
ApoB
Leukocyte_Esterase
Anion_Gap
Creatinine_Urine
Microalbumin_Creatinine_Ratio
Microalbumin
eGFR
Creatinine
BUN
Ketones
Triglycerides
Pulse
Systolic_Blood_Pressure
Diastolic_Blood_Pressure
O2SAT
Respiratory_Rate
Temperature
Protein_Urine

Figure S15

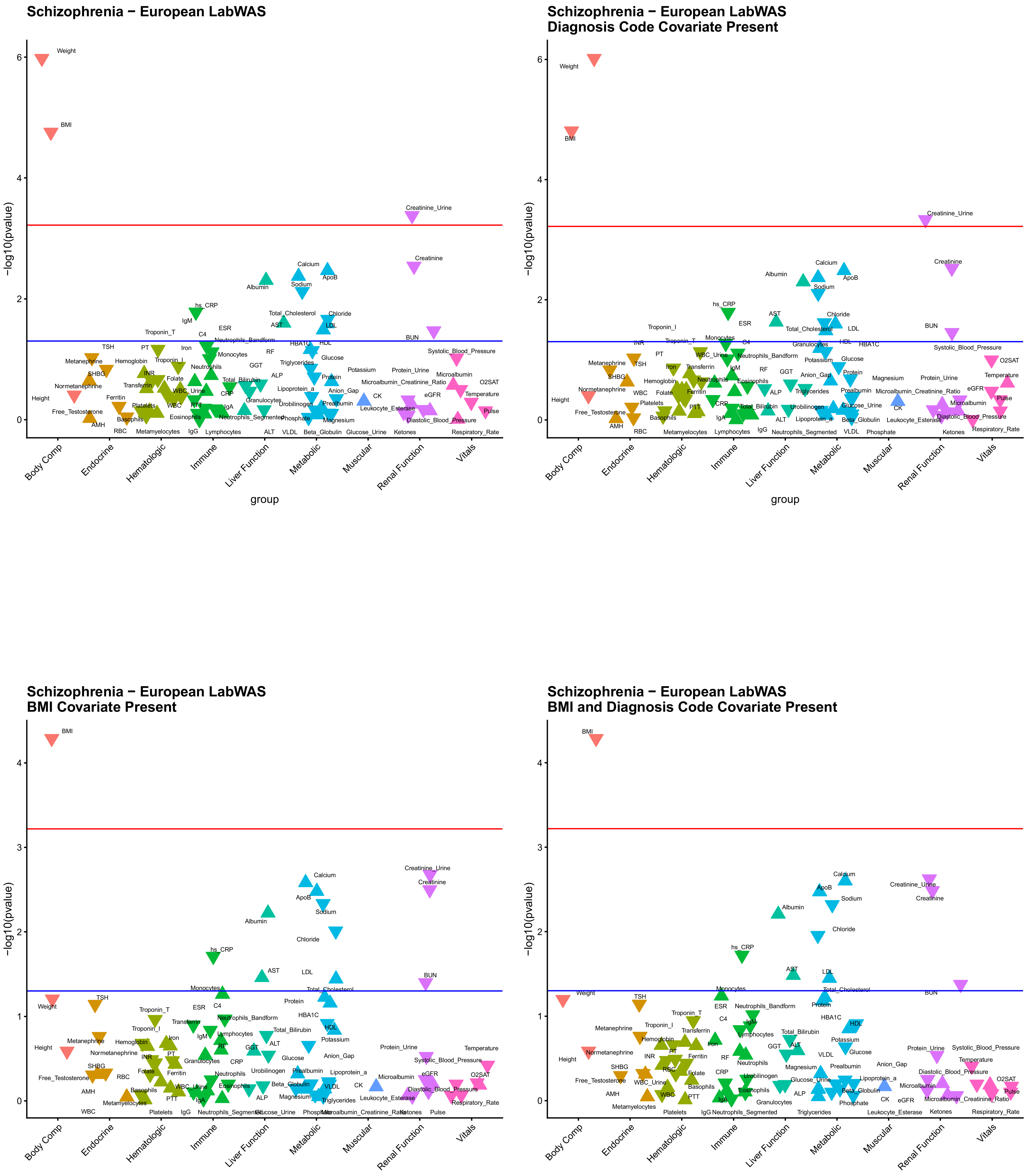

Schizophrenia − European LabWAS
−log10(pvalue)
0
2
4
6
Weight
BMI
Creatinine_Urine
Creatinine
Calcium
ApoB
Albumin
Sodium
hs_CRP
Body Comp
Endocrine
Hematologic
Immune
Liver Function
Metabolic
Muscular
Renal Function
Vitals
group
Schizophrenia − European LabWAS
Diagnosis Code Covariate Present
−log10(pvalue)
0
2
4
6
Weight
BMI
Creatinine_Urine
Creatinine
Calcium
ApoB
Albumin
Sodium
Body Comp
Endocrine
Hematologic
Immune
Liver Function
Metabolic
Muscular
Renal Function
Vitals
group
Schizophrenia − European LabWAS
BMI Covariate Present
−log10(pvalue)
0
1
2
3
4
BMI
Creatinine_Urine
Creatinine
Calcium
ApoB
Sodium
Albumin
Chloride
hs_CRP
AST
LDL
BUN
Body Comp
Endocrine
Hematologic
Immune
Liver Function
Metabolic
Muscular
Renal Function
Vitals
group
Schizophrenia − European LabWAS
BMI and Diagnosis Code Covariate Present
−log10(pvalue)
0
1
2
3
4
BMI
Calcium
ApoB
Sodium
Creatinine_Urine
Creatinine
Albumin
Chloride
hs_CRP
AST
LDL
Body Comp
Endocrine
Hematologic
Immune
Liver Function
Metabolic
Muscular
Renal Function
Vitals
group

Figure S16

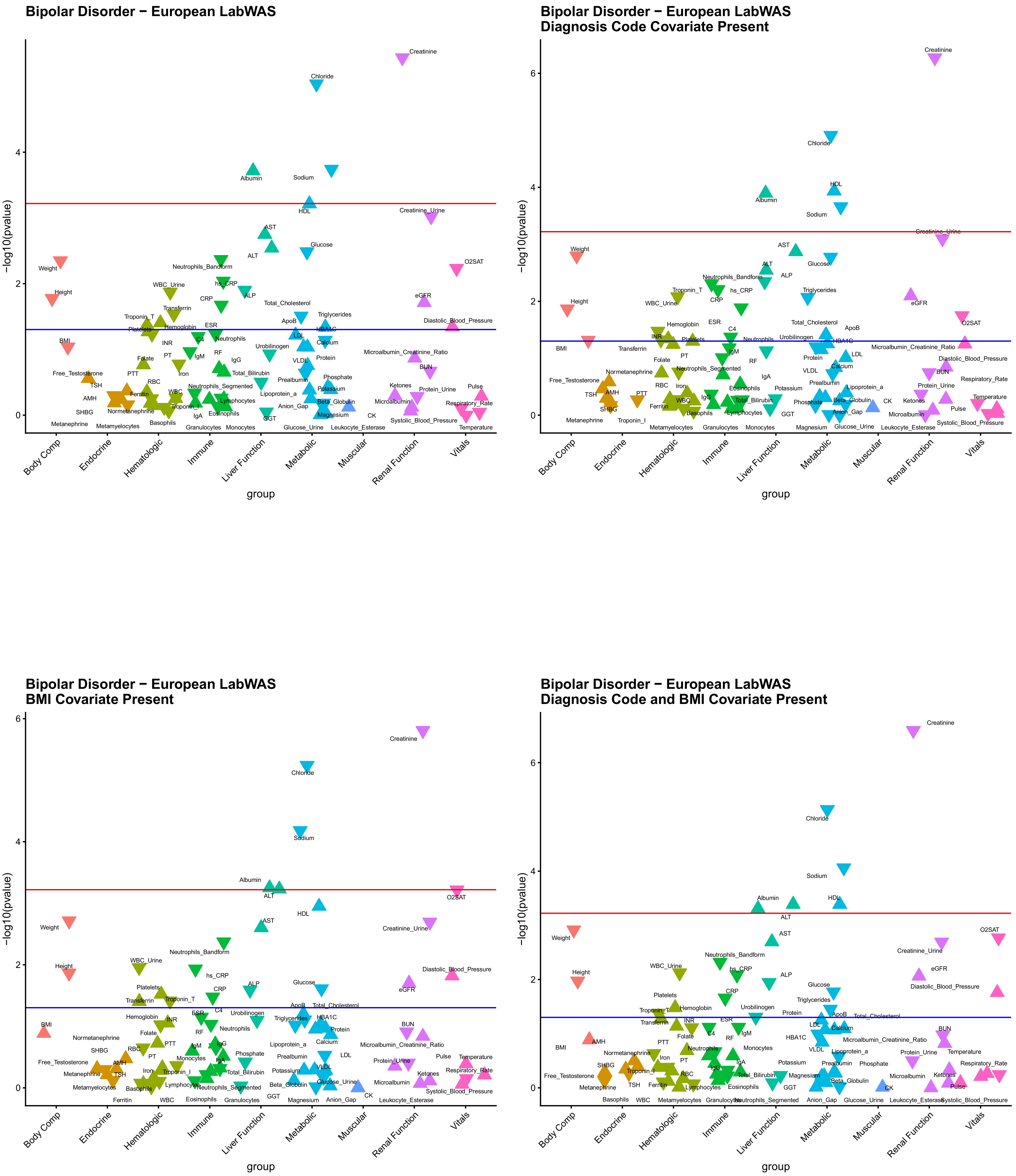

Bipolar Disorder − European LabWAS
Bipolar Disorder − European LabWAS
Diagnosis Code Covariate Present
Bipolar Disorder − European LabWAS
BMI Covariate Present
Bipolar Disorder − European LabWAS
Diagnosis Code and BMI Covariate Present
−log10(pvalue)
group
Body Comp
Endocrine
Hematologic
Immune
Liver Function
Metabolic
Muscular
Renal Function
Vitals

Figure S17

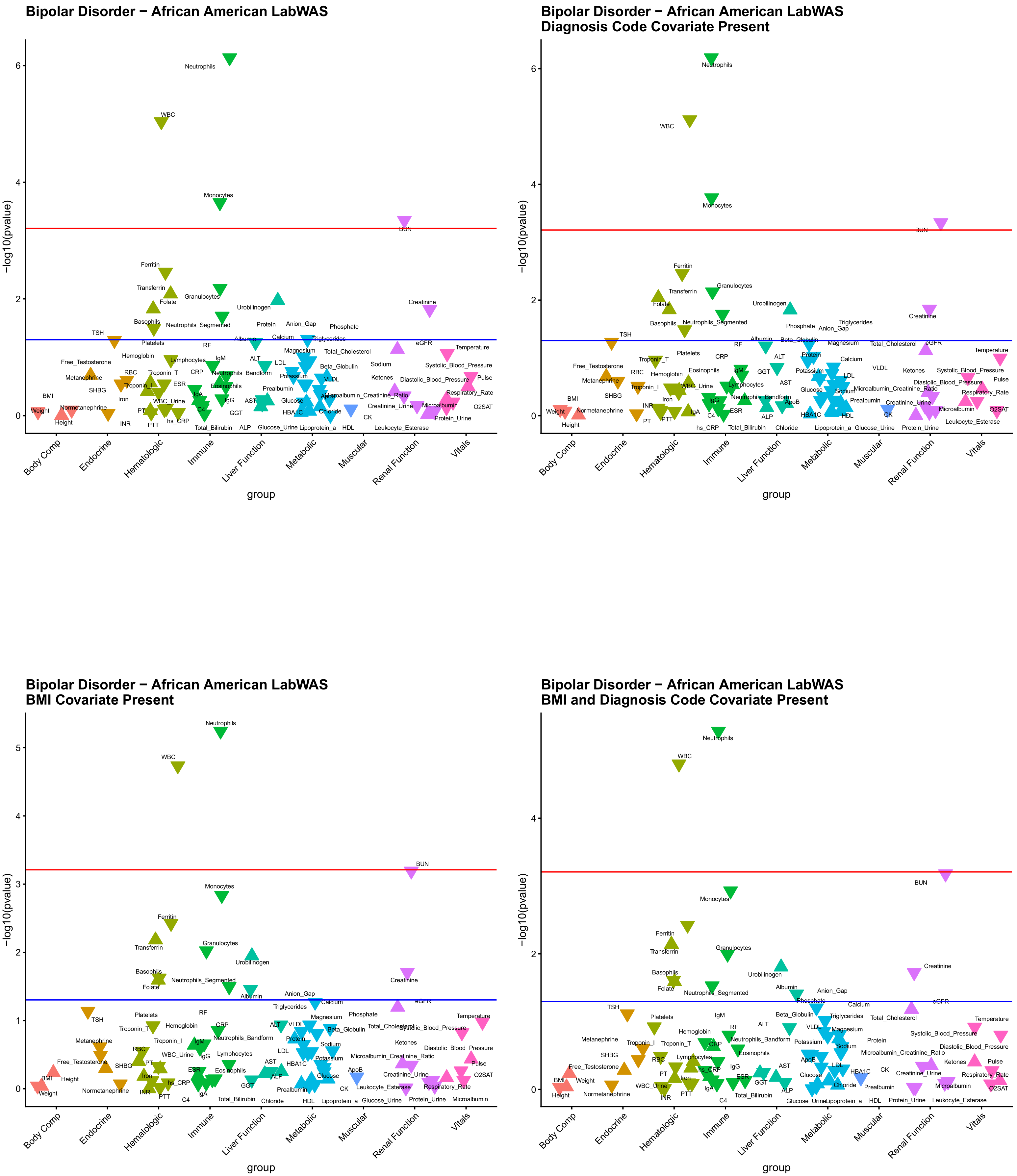

Bipolar Disorder − African American LabWAS
Bipolar Disorder − African American LabWAS
Diagnosis Code Covariate Present
Bipolar Disorder − African American LabWAS
BMI Covariate Present
Bipolar Disorder − African American LabWAS
BMI and Diagnosis Code Covariate Present
−log10(pvalue)
group
Body Comp
Endocrine
Hematologic
Immune
Liver Function
Metabolic
Muscular
Renal Function
Vitals
Neutrophils
WBC
Monocytes
BUN
Ferritin
Transferrin
Folate
Granulocytes
Urobilinogen
Creatinine
Basophils
Neutrophils_Segmented
TSH
eGFR

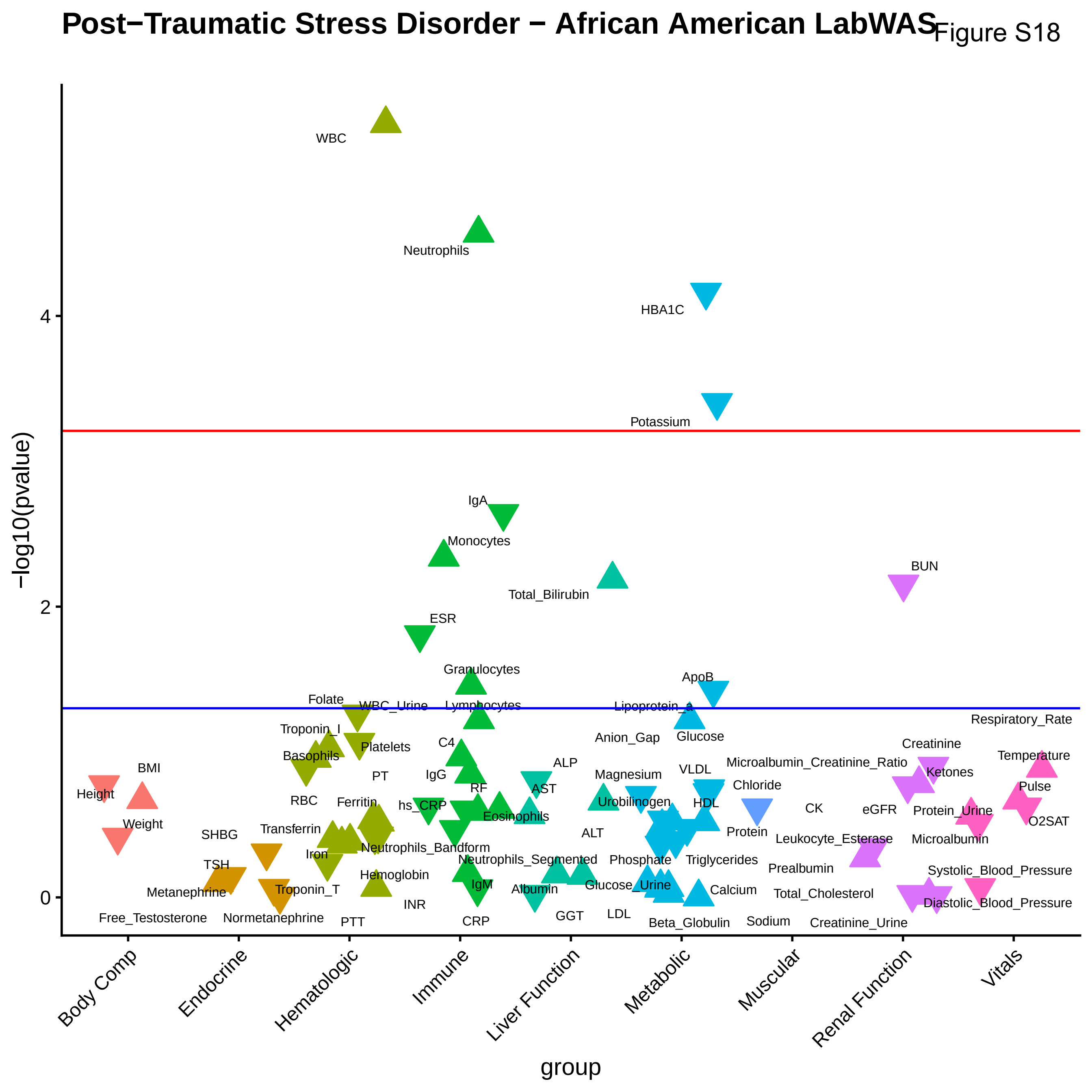
Post−Traumatic Stress Disorder − African American LabWAS
Figure S18
−log10(pvalue)
group
0
2
4
Body Comp
Endocrine
Hematologic
Immune
Liver Function
Metabolic
Muscular
Renal Function
Vitals
WBC
Neutrophils
HBA1C
Potassium
IgA
Monocytes
Total_Bilirubin
BUN
ESR
Granulocytes
ApoB
Folate
WBC_Urine
Lymphocytes
Lipoprotein_a
Respiratory_Rate
Troponin_I
Anion_Gap
Glucose
Creatinine
Basophils
Platelets
C4
Microalbumin_Creatinine_Ratio
Temperature
BMI
PT
IgG
ALP
Magnesium
VLDL
Ketones
Height
RF
AST
Chloride
Pulse
RBC
Ferritin
hs_CRP
Urobilinogen
HDL
CK
eGFR
Protein_Urine
Eosinophils
Weight
O2SAT
Transferrin
SHBG
ALT
Protein
Leukocyte_Esterase
Microalbumin
Iron
Neutrophils_Bandform
TSH
Neutrophils_Segmented
Phosphate
Triglycerides
Prealbumin
Systolic_Blood_Pressure
Hemoglobin
IgM
Metanephrine
Troponin_T
Albumin
Glucose_Urine
Calcium
Total_Cholesterol
INR
Diastolic_Blood_Pressure
Free_Testosterone
Normetanephrine
GGT
LDL
PTT
CRP
Beta_Globulin
Sodium
Creatinine_Urine

Figure S19

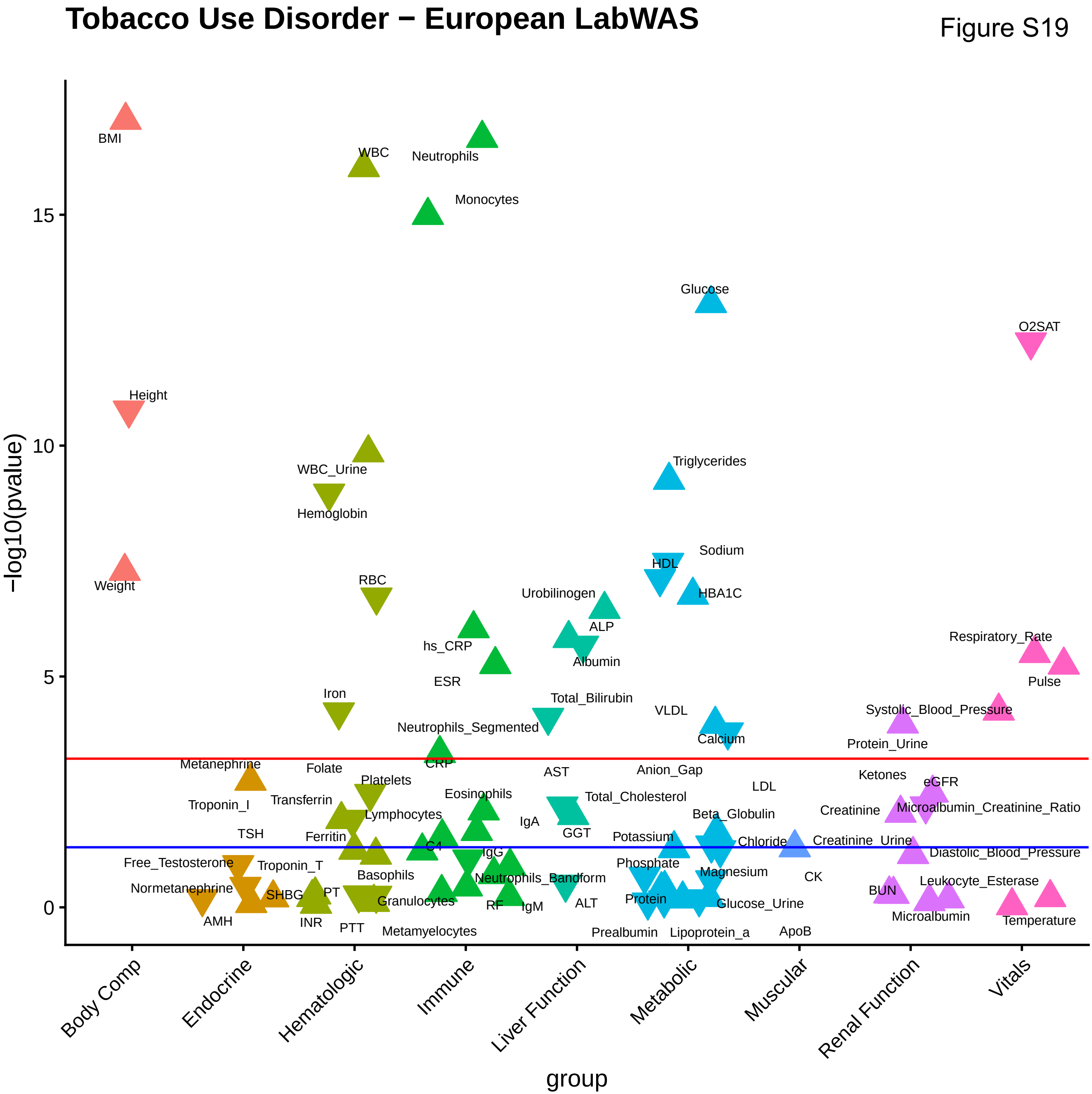
Tobacco Use Disorder − European LabWAS
−log10(pvalue)
group
0
5
10
15
Body Comp
Endocrine
Hematologic
Immune
Liver Function
Metabolic
Muscular
Renal Function
Vitals
BMI
Height
Weight
WBC
Neutrophils
Monocytes
WBC_Urine
Hemoglobin
RBC
Iron
Folate
Platelets
Transferrin
Lymphocytes
Ferritin
Basophils
PT
INR
PTT
Granulocytes
Metamyelocytes
hs_CRP
ESR
Neutrophils_Segmented
CRP
Eosinophils
C4
IgG
IgA
IgM
RF
Neutrophils_Bandform
Metanephrine
Troponin_I
TSH
Free_Testosterone
Troponin_T
Normetanephrine
SHBG
AMH
Urobilinogen
ALP
Albumin
Total_Bilirubin
AST
Total_Cholesterol
GGT
ALT
Glucose
Triglycerides
Sodium
HDL
HBA1C
VLDL
Calcium
Anion_Gap
LDL
Beta_Globulin
Potassium
Chloride
Phosphate
Magnesium
Protein
Glucose_Urine
Prealbumin
Lipoprotein_a
CK
ApoB
Protein_Urine
Ketones
eGFR
Creatinine
Microalbumin_Creatinine_Ratio
Creatinine_Urine
Diastolic_Blood_Pressure
Leukocyte_Esterase
BUN
Microalbumin
O2SAT
Respiratory_Rate
Pulse
Systolic_Blood_Pressure
Temperature

### Externalizing Factor − European LabWAS

Figure S20

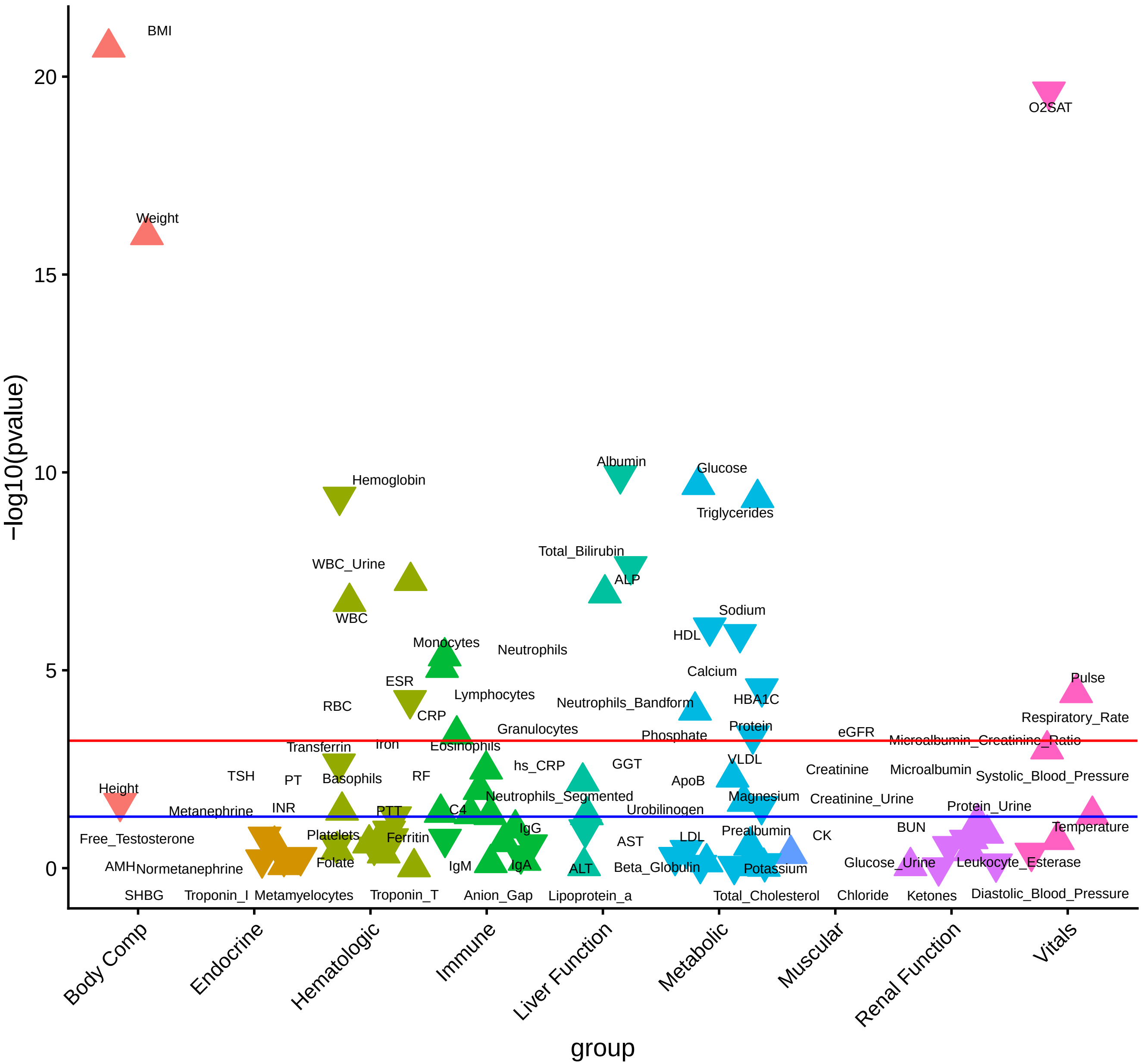

### Problematic Alcohol Use − European LabWAS

Figure S21

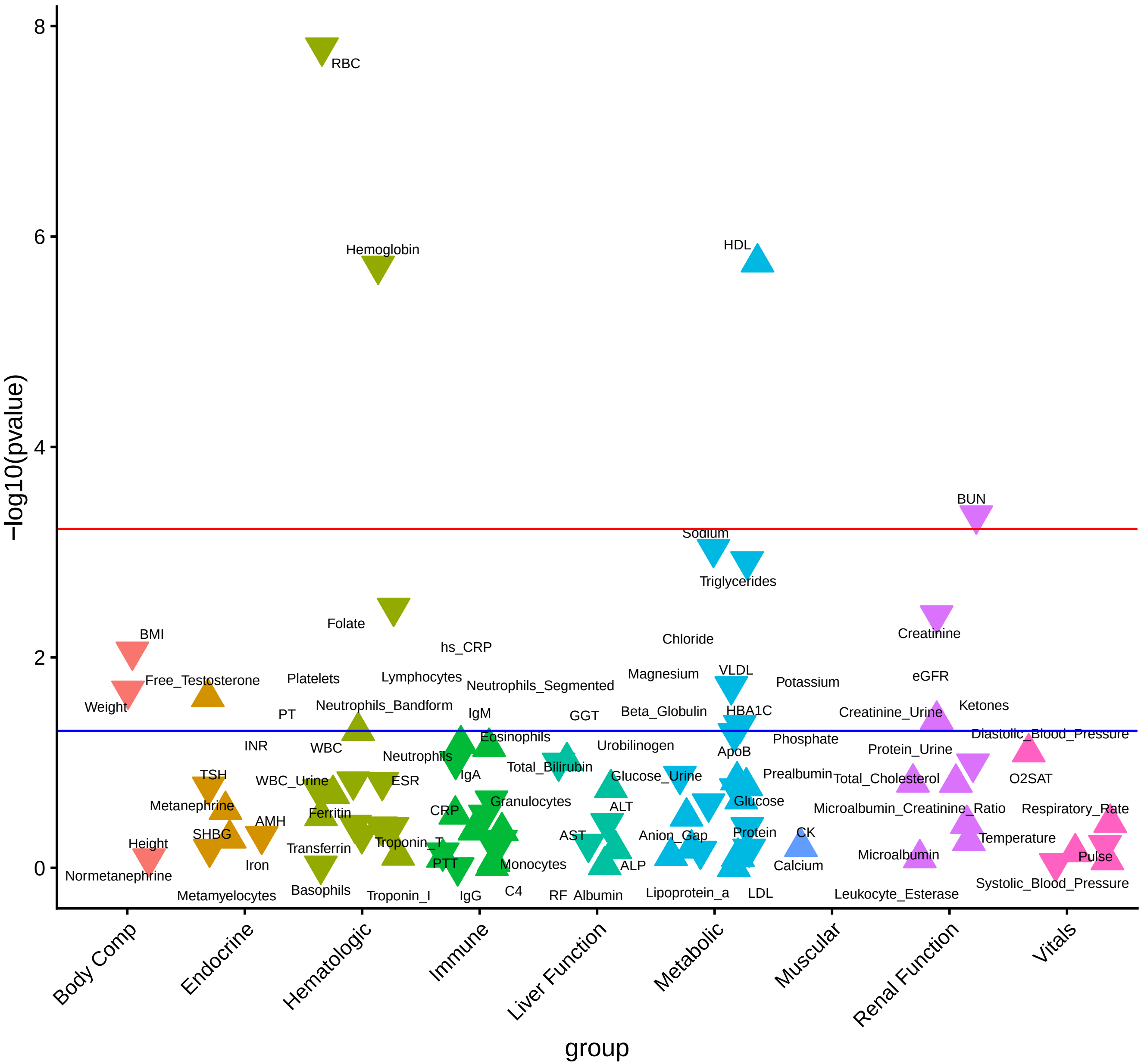

Figure S22

### Alcohol Use Disorder − European LabWAS

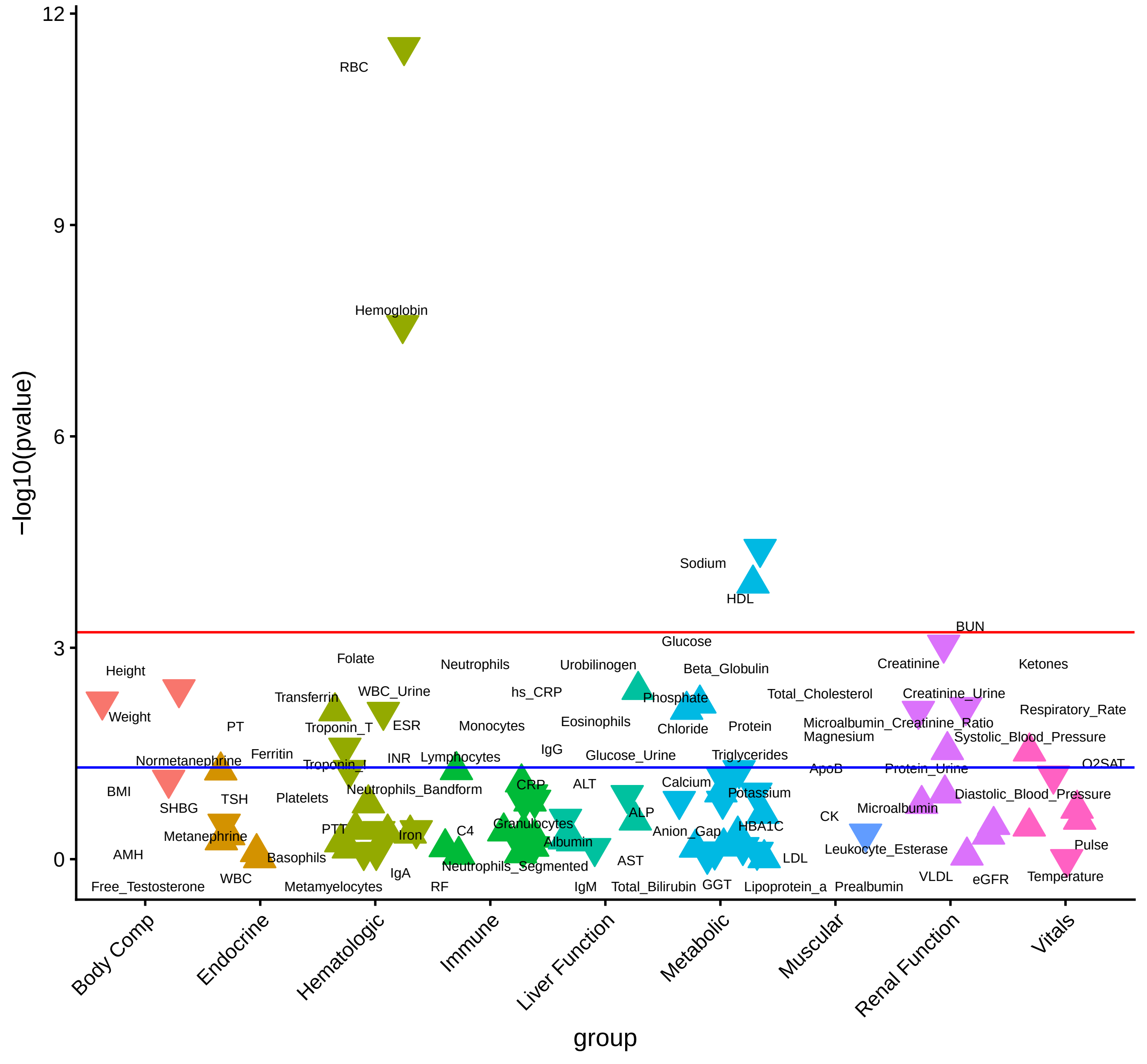

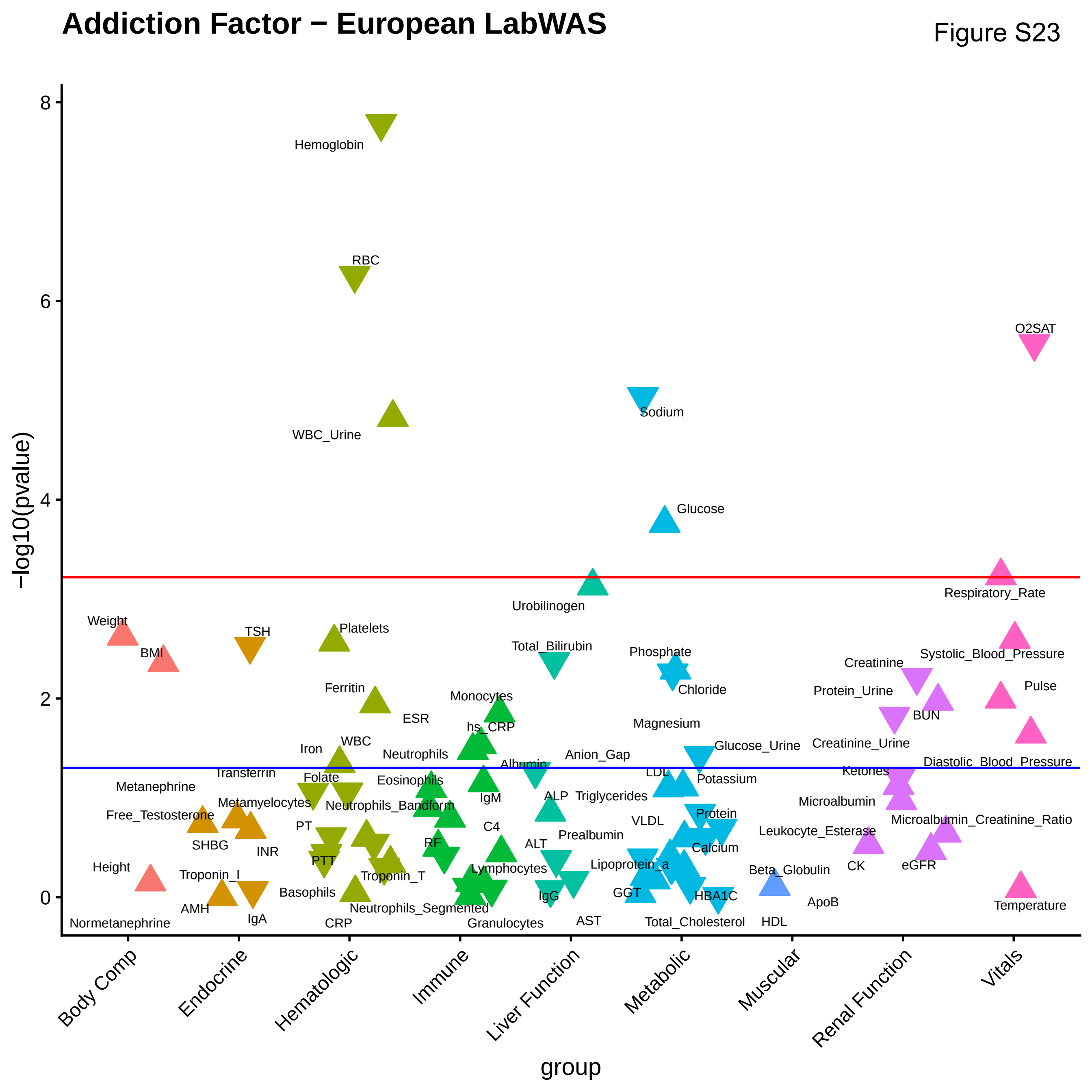
Addiction Factor − European LabWAS
Figure S23
−log10(pvalue)
group
Body Comp
Endocrine
Hematologic
Immune
Liver Function
Metabolic
Muscular
Renal Function
Vitals
Hemoglobin
RBC
WBC_Urine
O2SAT
Sodium
Glucose
Urobilinogen
Respiratory_Rate
Weight
BMI
TSH
Platelets
Total_Bilirubin
Phosphate
Chloride
Systolic_Blood_Pressure
Creatinine
Pulse
Protein_Urine
BUN
Ferritin
ESR
Monocytes
hs_CRP
Magnesium
Glucose_Urine
Creatinine_Urine
Iron
WBC
Neutrophils
Albumin
Anion_Gap
Diastolic_Blood_Pressure
Transferrin
Folate
Eosinophils
LDL
Potassium
Ketones
Metanephrine
Metamyelocytes
Neutrophils_Bandform
IgM
ALP
Triglycerides
Microalbumin
Free_Testosterone
PT
C4
VLDL
Protein
Microalbumin_Creatinine_Ratio
SHBG
INR
RF
ALT
Prealbumin
Leukocyte_Esterase
Calcium
PTT
Lymphocytes
Lipoprotein_a
CK
eGFR
Height
Troponin_I
Troponin_T
Beta_Globulin
Basophils
IgG
GGT
HBA1C
ApoB
AMH
IgA
Neutrophils_Segmented
AST
Temperature
Normetanephrine
CRP
Granulocytes
Total_Cholesterol
HDL

Figure S24

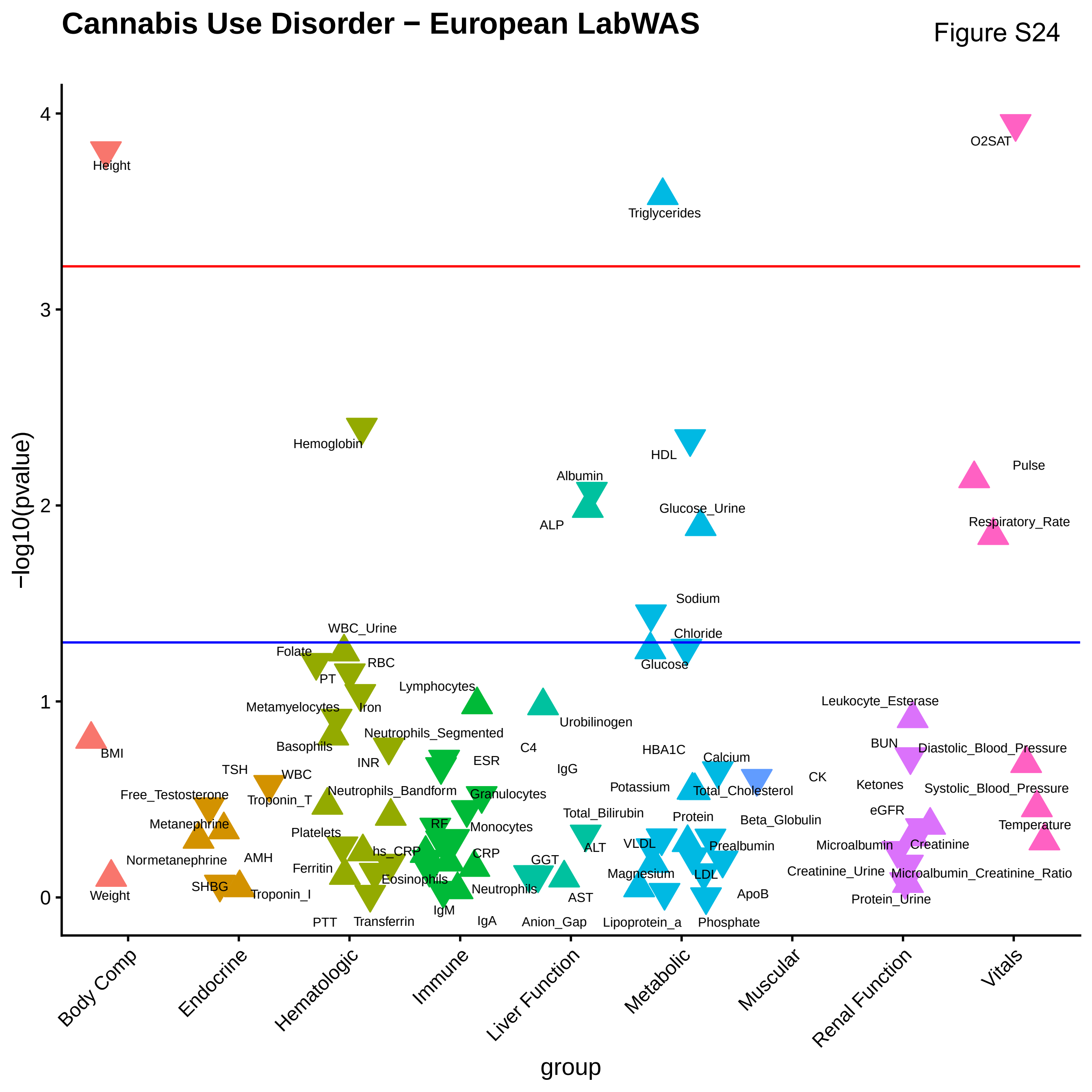
Cannabis Use Disorder − European LabWAS
−log10(pvalue)
group
0
1
2
3
4
Body Comp
Endocrine
Hematologic
Immune
Liver Function
Metabolic
Muscular
Renal Function
Vitals
Height
BMI
Weight
O2SAT
Pulse
Respiratory_Rate
Diastolic_Blood_Pressure
Systolic_Blood_Pressure
Temperature
Triglycerides
HDL
Glucose_Urine
Sodium
Chloride
Glucose
HBA1C
Calcium
Potassium
Total_Cholesterol
Protein
Beta_Globulin
VLDL
Prealbumin
Magnesium
LDL
ApoB
Lipoprotein_a
Phosphate
CK
Hemoglobin
WBC_Urine
Folate
RBC
PT
Iron
Metamyelocytes
Basophils
INR
WBC
Neutrophils_Bandform
Platelets
hs_CRP
Ferritin
Eosinophils
PTT
Transferrin
Lymphocytes
Neutrophils_Segmented
ESR
Granulocytes
RF
Monocytes
CRP
Neutrophils
IgM
IgA
Albumin
ALP
Urobilinogen
C4
IgG
Total_Bilirubin
ALT
GGT
AST
Anion_Gap
TSH
Free_Testosterone
Troponin_T
Metanephrine
Normetanephrine
AMH
SHBG
Troponin_I
Leukocyte_Esterase
BUN
Ketones
eGFR
Microalbumin
Creatinine
Creatinine_Urine
Microalbumin_Creatinine_Ratio
Protein_Urine

Figure S25

### Tobacco Use Disorder − African American LabWAS

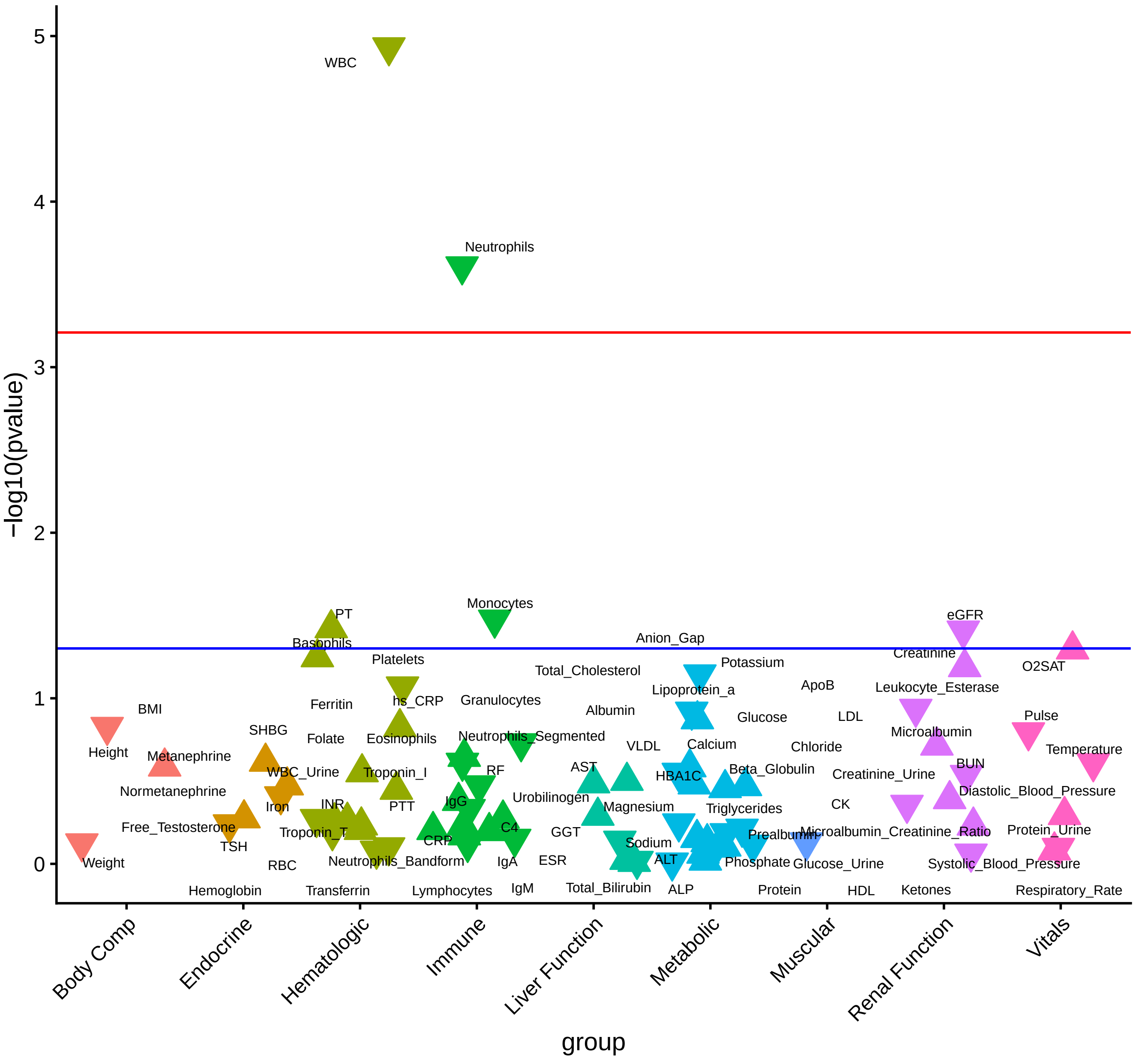

Figure S26

### Cannabis Use Disorder − African American LabWAS

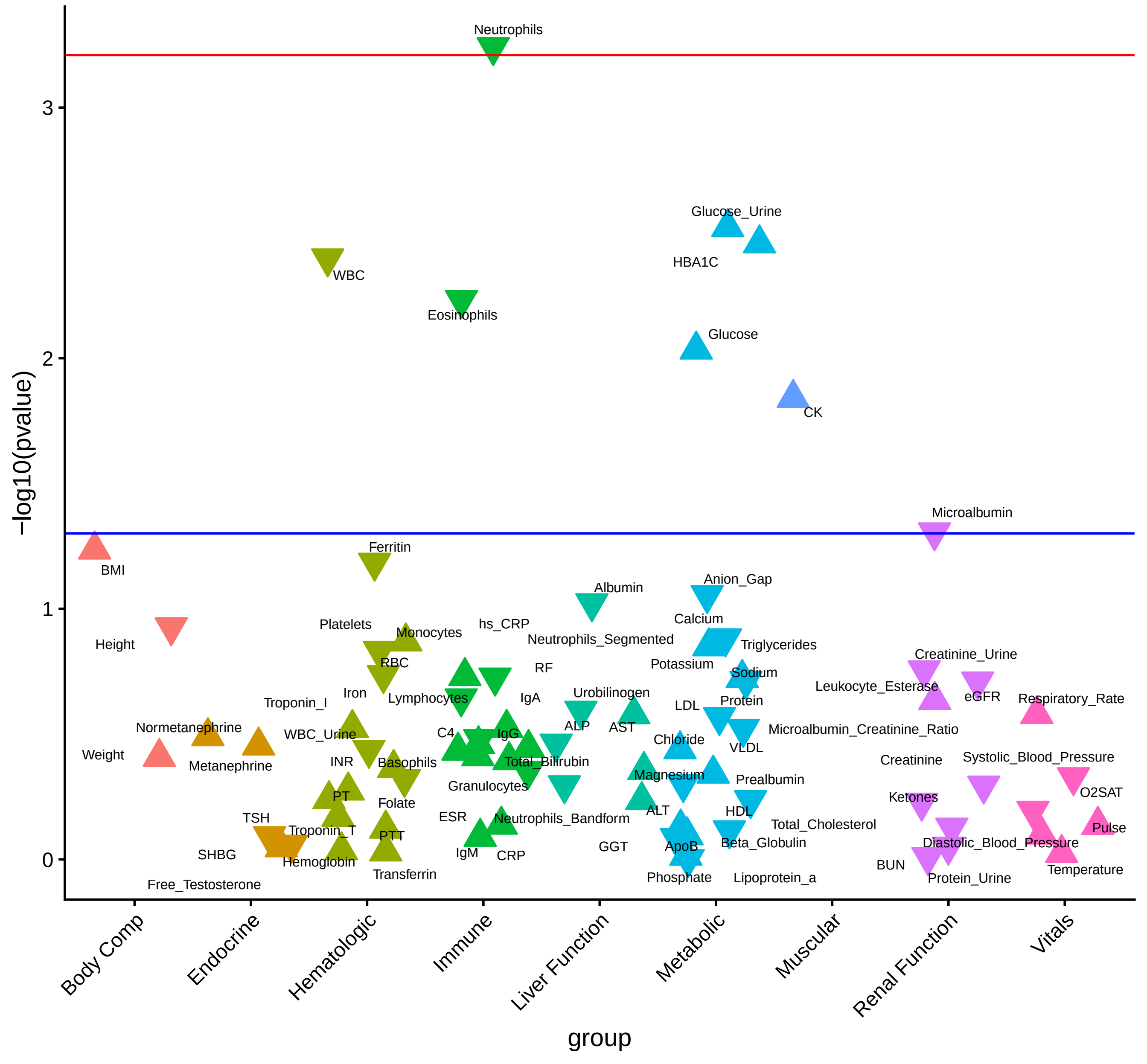

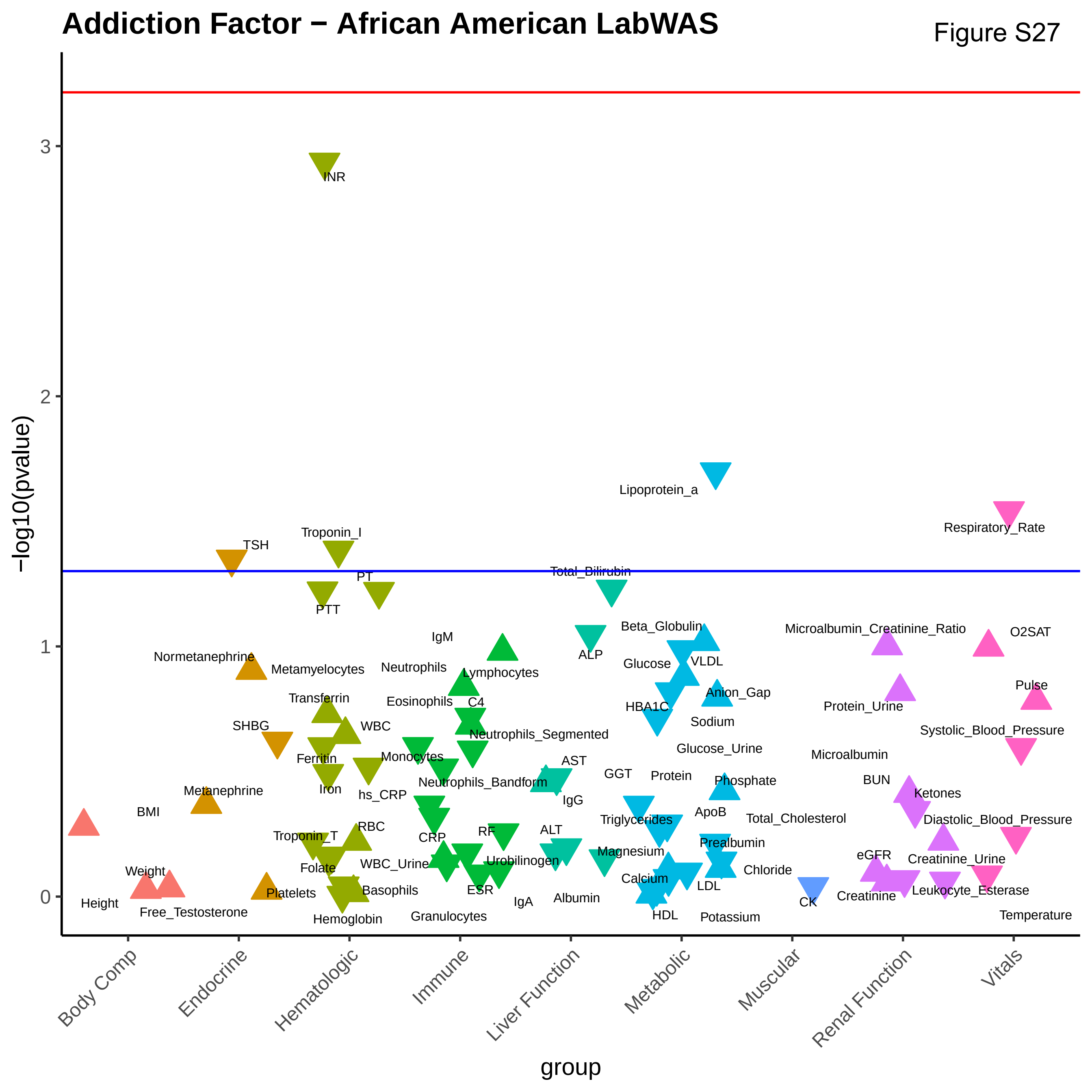

Addiction Factor − African American LabWAS
Figure S27
−log10(pvalue)
group
Body Comp
Endocrine
Hematologic
Immune
Liver Function
Metabolic
Muscular
Renal Function
Vitals
INR
Lipoprotein_a
Respiratory_Rate
TSH
Troponin_I
Total_Bilirubin
PT
PTT
Beta_Globulin
Microalbumin_Creatinine_Ratio
O2SAT
IgM
ALP
Normetanephrine
Metamyelocytes
Neutrophils
Lymphocytes
Glucose
VLDL
Pulse
Transferrin
Eosinophils
C4
HBA1C
Anion_Gap
Protein_Urine
SHBG
WBC
Sodium
Systolic_Blood_Pressure
Neutrophils_Segmented
Glucose_Urine
Ferritin
Monocytes
Microalbumin
AST
GGT
Protein
Iron
Neutrophils_Bandform
Phosphate
BUN
Metanephrine
hs_CRP
IgG
Ketones
BMI
ApoB
Total_Cholesterol
Triglycerides
Diastolic_Blood_Pressure
RBC
Troponin_T
CRP
RF
ALT
Prealbumin
Magnesium
eGFR
Creatinine_Urine
Folate
WBC_Urine
Urobilinogen
Chloride
Weight
Calcium
LDL
Basophils
ESR
Leukocyte_Esterase
Platelets
Creatinine
Height
Albumin
IgA
CK
Free_Testosterone
Granulocytes
HDL
Potassium
Hemoglobin
Temperature

Figure S28

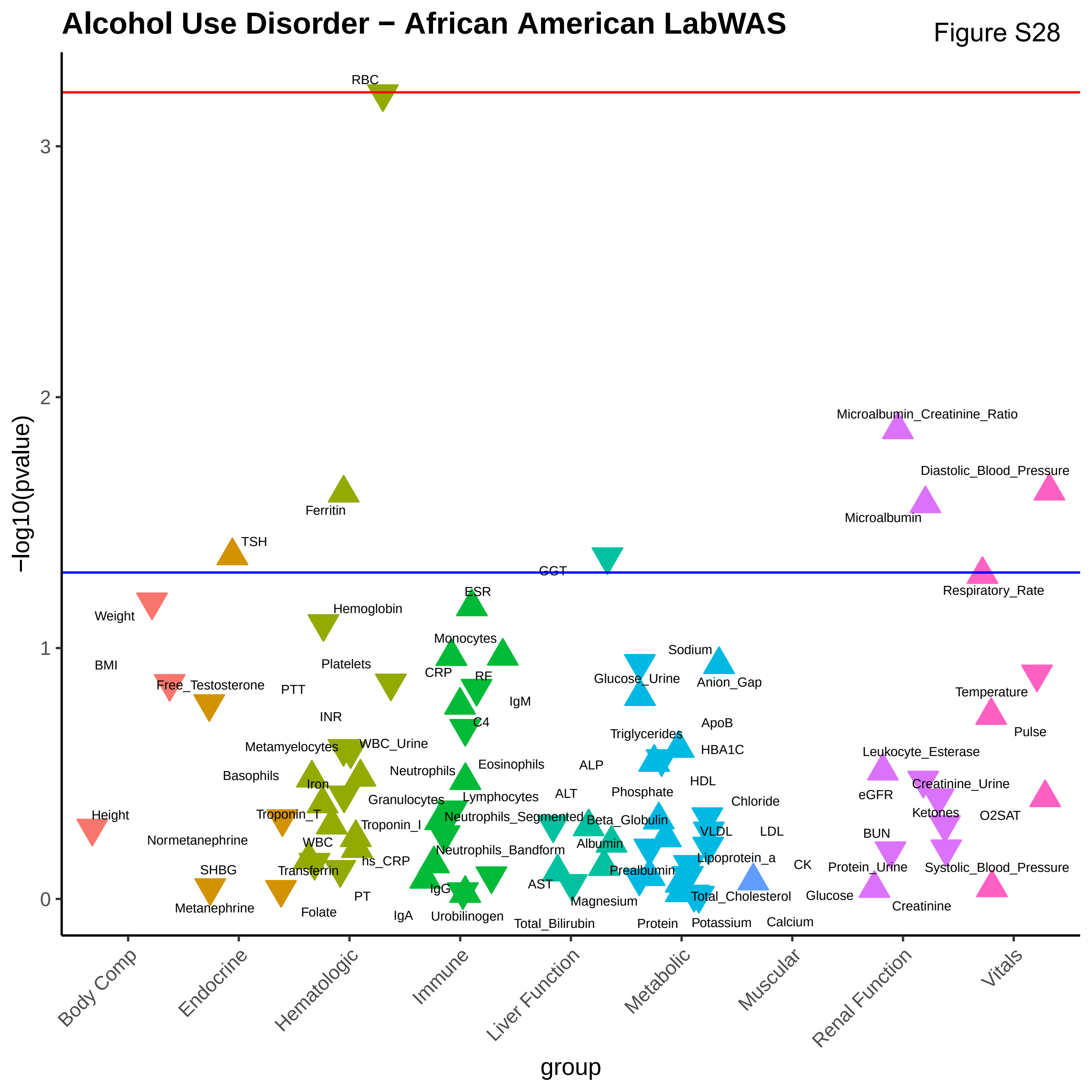
Alcohol Use Disorder − African American LabWAS
−log10(pvalue)
group
Body Comp
Endocrine
Hematologic
Immune
Liver Function
Metabolic
Muscular
Renal Function
Vitals
RBC
Ferritin
TSH
GGT
Microalbumin_Creatinine_Ratio
Diastolic_Blood_Pressure
Microalbumin
Respiratory_Rate
Weight
BMI
Free_Testosterone
PTT
INR
Hemoglobin
Platelets
Metamyelocytes
WBC_Urine
Basophils
Iron
Height
Troponin_T
Normetanephrine
WBC
Transferrin
SHBG
Metanephrine
Folate
PT
IgA
ESR
Monocytes
CRP
RF
IgM
C4
Neutrophils
Eosinophils
Granulocytes
Lymphocytes
Troponin_I
hs_CRP
Neutrophils_Segmented
Neutrophils_Bandform
IgG
Urobilinogen
ALP
ALT
Beta_Globulin
Albumin
AST
Magnesium
Total_Bilirubin
Sodium
Glucose_Urine
Anion_Gap
ApoB
Triglycerides
HBA1C
HDL
Phosphate
Chloride
VLDL
LDL
Lipoprotein_a
Prealbumin
CK
Total_Cholesterol
Protein
Potassium
Calcium
Glucose
Leukocyte_Esterase
eGFR
Creatinine_Urine
Ketones
BUN
Protein_Urine
Creatinine
Temperature
Pulse
O2SAT
Systolic_Blood_Pressure

**Autism Spectrum Disorder − European LabWAS**

Figure S29

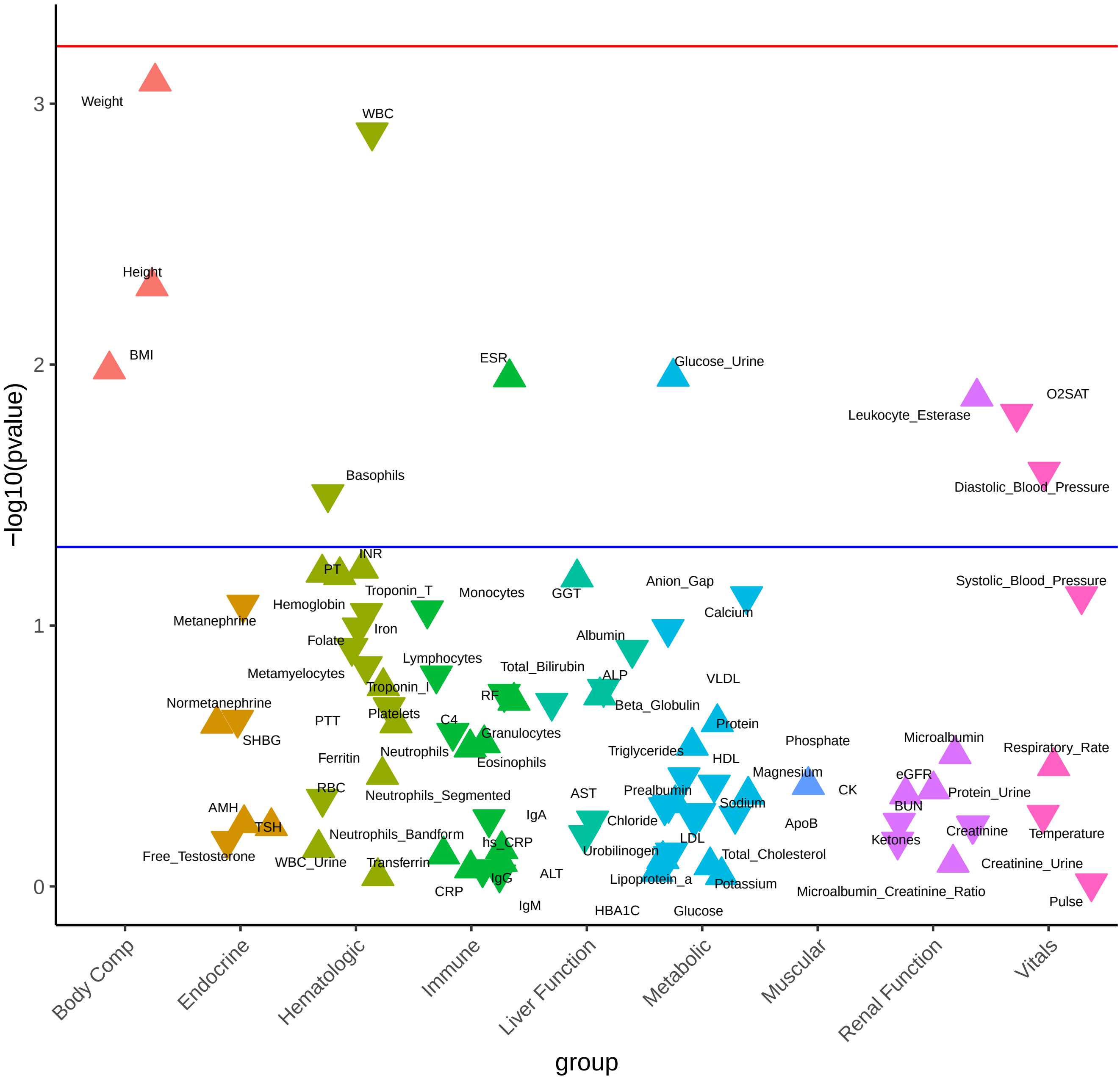

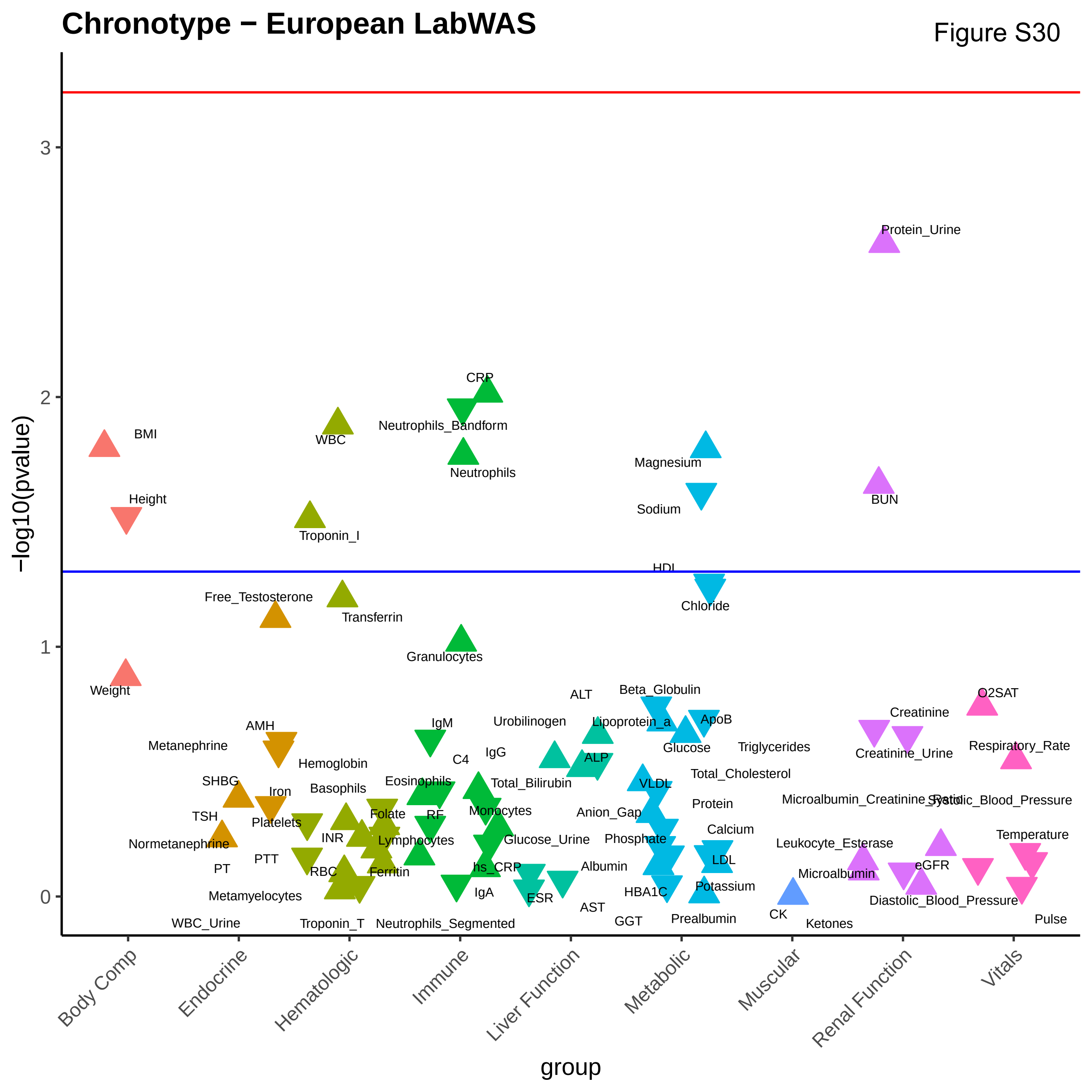
Chronotype − European LabWAS
−log10(pvalue)
group
Body Comp
Endocrine
Hematologic
Immune
Liver Function
Metabolic
Muscular
Renal Function
Vitals
0
1
2
3
BMI
Height
Weight
WBC
Troponin_I
Transferrin
Free_Testosterone
CRP
Neutrophils_Bandform
Neutrophils
Granulocytes
Magnesium
Sodium
HDL
Chloride
Protein_Urine
BUN
AMH
Metanephrine
Hemoglobin
SHBG
Iron
TSH
Platelets
Normetanephrine
PT
PTT
INR
RBC
Metamyelocytes
WBC_Urine
Troponin_T
Basophils
Folate
Lymphocytes
Ferritin
Eosinophils
IgM
C4
IgG
RF
Monocytes
hs_CRP
IgA
Neutrophils_Segmented
Total_Bilirubin
Glucose_Urine
Urobilinogen
ALT
Lipoprotein_a
ALP
Albumin
ESR
AST
GGT
Beta_Globulin
ApoB
Glucose
Triglycerides
Total_Cholesterol
VLDL
Anion_Gap
Protein
Phosphate
Calcium
LDL
HBA1C
Potassium
Prealbumin
CK
Ketones
Creatinine
Creatinine_Urine
Microalbumin_Creatinine_Ratio
Leukocyte_Esterase
Microalbumin
eGFR
Diastolic_Blood_Pressure
O2SAT
Respiratory_Rate
Systolic_Blood_Pressure
Temperature
Pulse

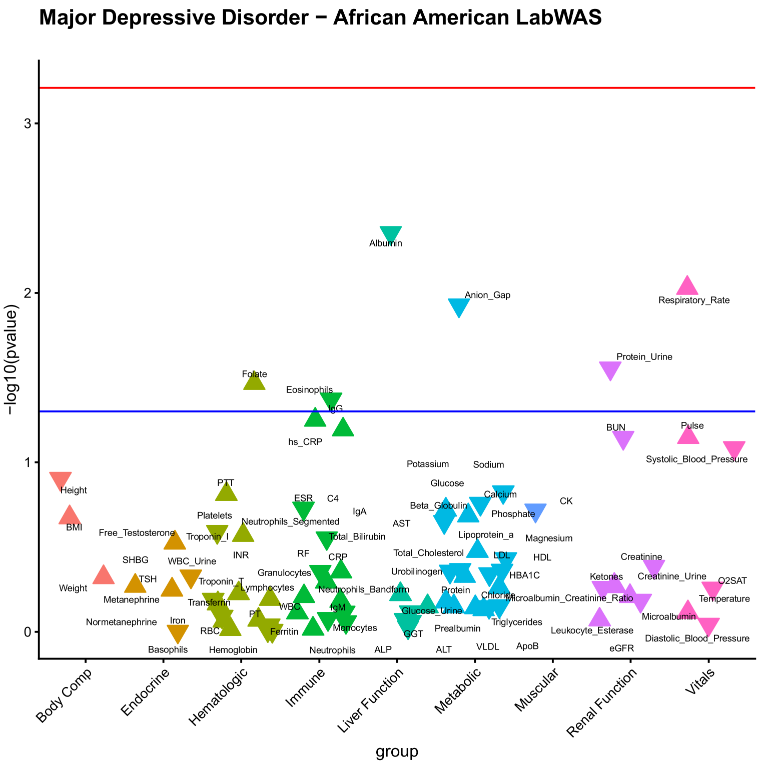
Major Depressive Disorder − African American LabWAS
−log10(pvalue)
group
Body Comp
Endocrine
Hematologic
Immune
Liver Function
Metabolic
Muscular
Renal Function
Vitals

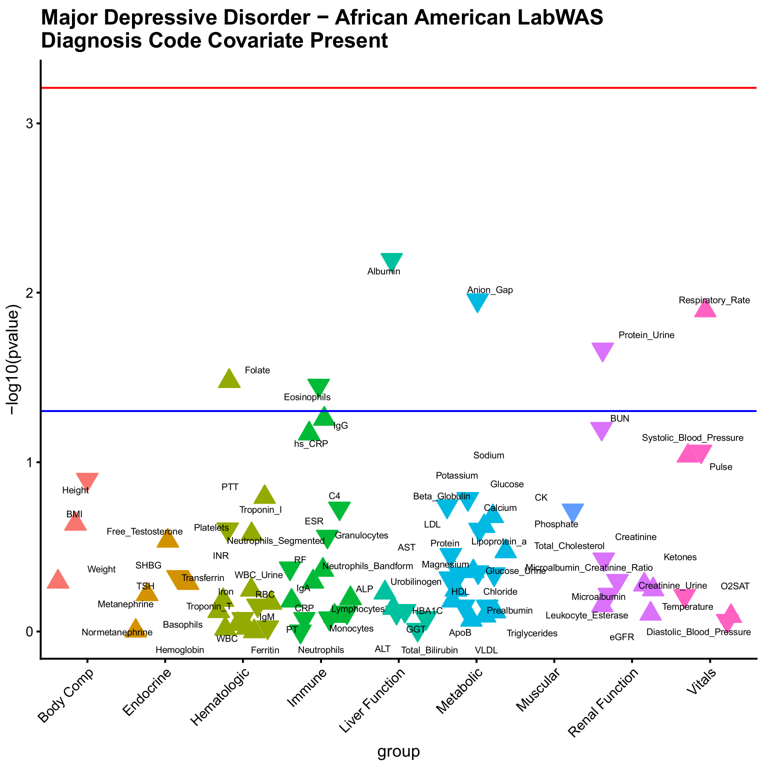
Major Depressive Disorder − African American LabWAS
Diagnosis Code Covariate Present
−log10(pvalue)
group
Body Comp
Endocrine
Hematologic
Immune
Liver Function
Metabolic
Muscular
Renal Function
Vitals

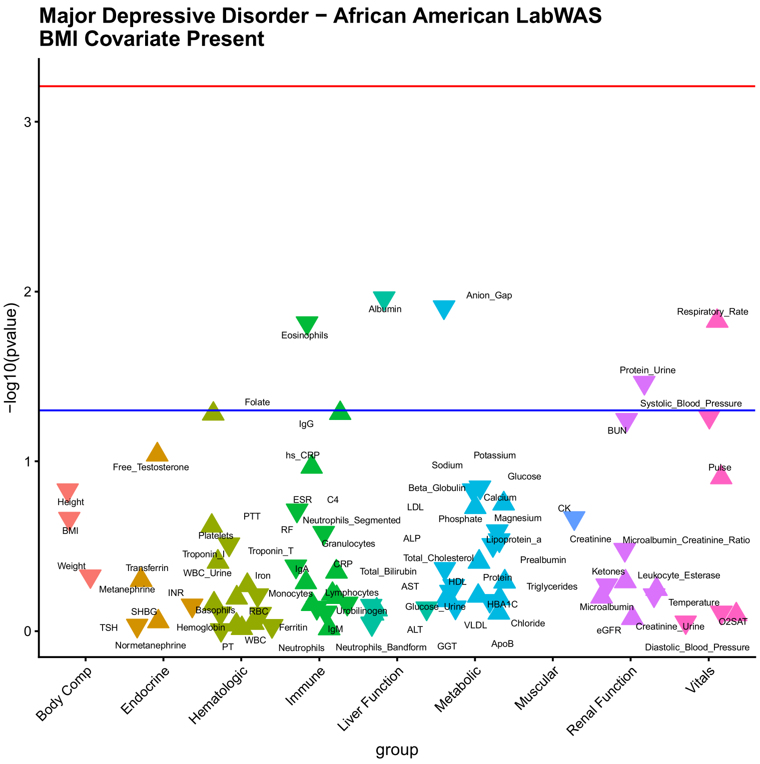
Major Depressive Disorder − African American LabWAS
BMI Covariate Present
−log10(pvalue)
group
Body Comp
Endocrine
Hematologic
Immune
Liver Function
Metabolic
Muscular
Renal Function
Vitals

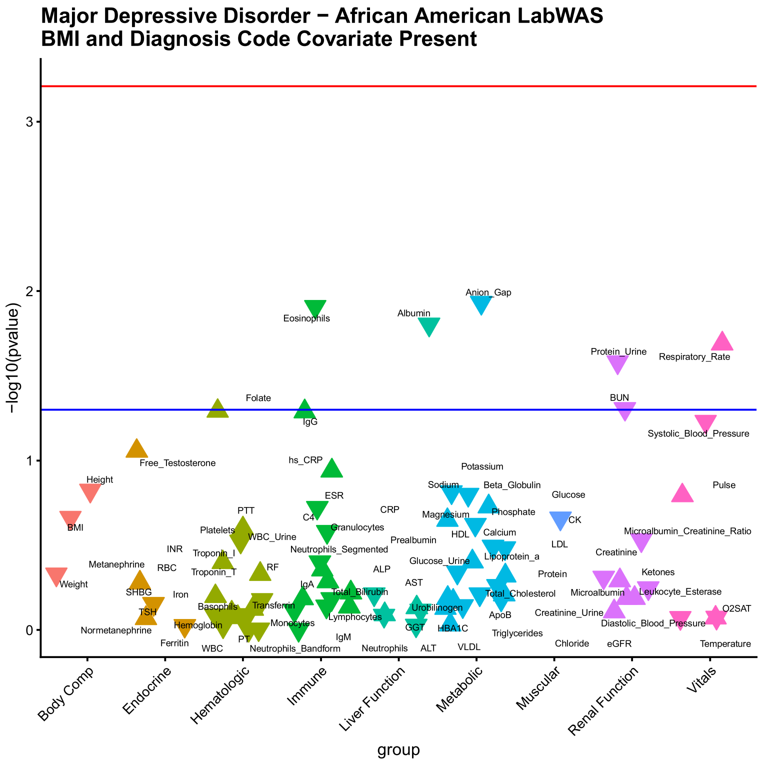
Major Depressive Disorder − African American LabWAS
BMI and Diagnosis Code Covariate Present
−log10(pvalue)
group
Body Comp
Endocrine
Hematologic
Immune
Liver Function
Metabolic
Muscular
Renal Function
Vitals

### Opioid Use Disorder − African American LabWAS

Figure S32

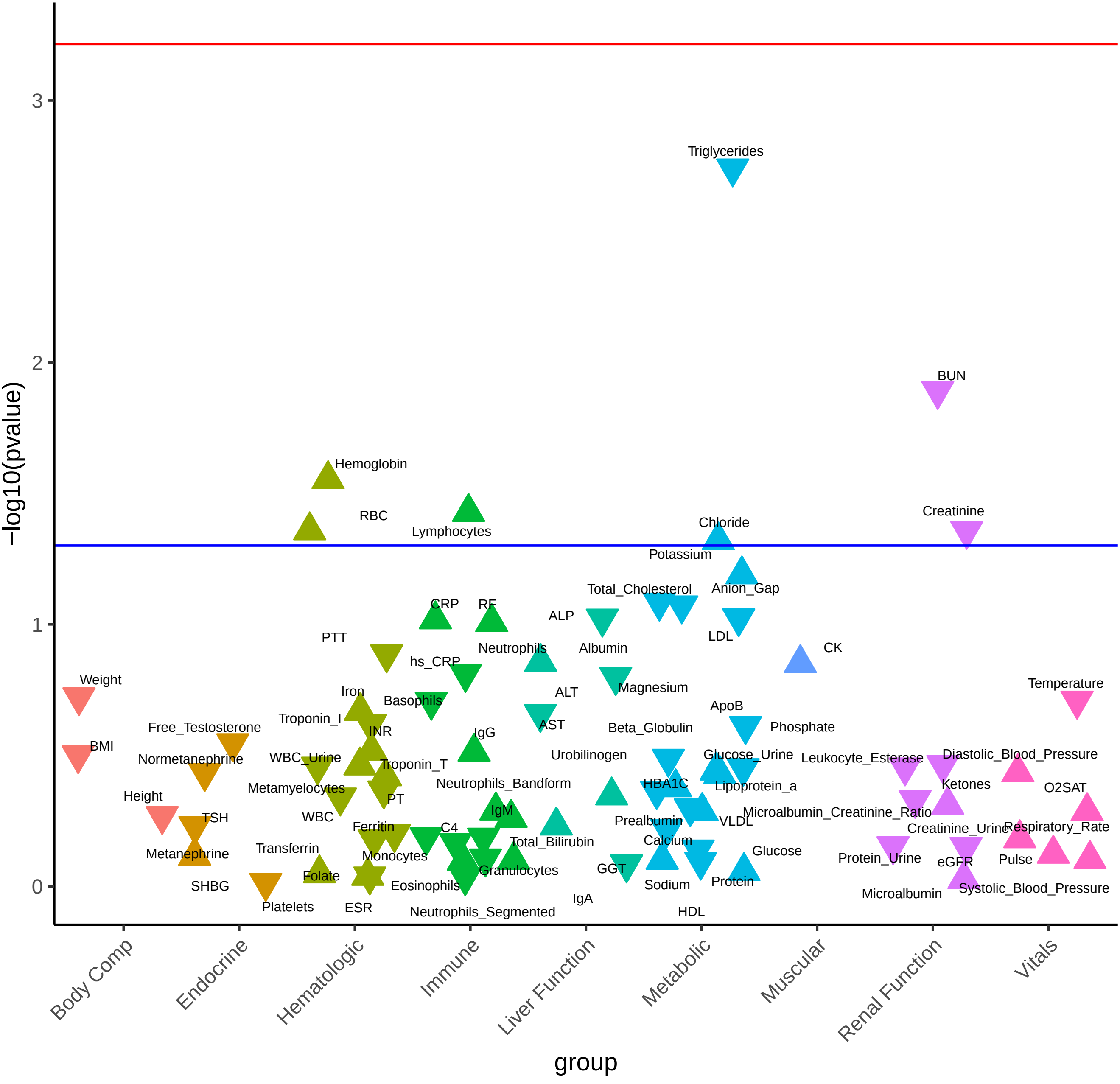

Figure S33

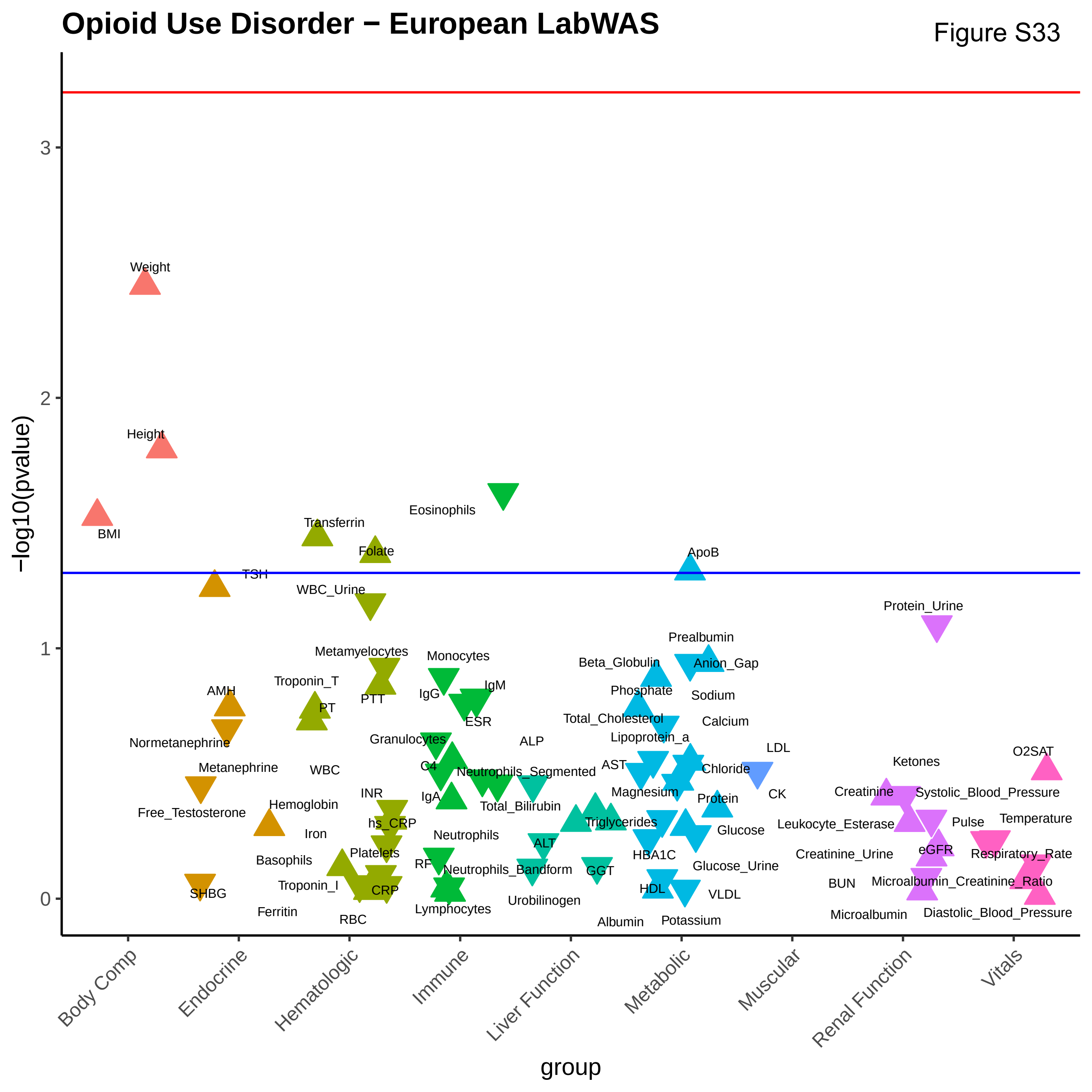
Opioid Use Disorder − European LabWAS
−log10(pvalue)
group
0
1
2
3
Body Comp
Endocrine
Hematologic
Immune
Liver Function
Metabolic
Muscular
Renal Function
Vitals
Weight
Height
BMI
Transferrin
Eosinophils
Folate
ApoB
TSH
WBC_Urine
Protein_Urine
Prealbumin
Metamyelocytes
Monocytes
Beta_Globulin
Anion_Gap
Troponin_T
AMH
IgG
IgM
PTT
Phosphate
Sodium
PT
ESR
Total_Cholesterol
Calcium
Normetanephrine
Granulocytes
ALP
Lipoprotein_a
LDL
O2SAT
Ketones
Metanephrine
WBC
C4
Neutrophils_Segmented
AST
Chloride
CK
INR
IgA
Magnesium
Protein
Creatinine
Systolic_Blood_Pressure
Hemoglobin
Total_Bilirubin
Free_Testosterone
hs_CRP
Triglycerides
Leukocyte_Esterase
Pulse
Temperature
Iron
Neutrophils
Glucose
ALT
HBA1C
eGFR
Respiratory_Rate
Platelets
Creatinine_Urine
Basophils
RF
Glucose_Urine
Neutrophils_Bandform
GGT
Troponin_I
BUN
Microalbumin_Creatinine_Ratio
HDL
CRP
SHBG
VLDL
Urobilinogen
Lymphocytes
Ferritin
Diastolic_Blood_Pressure
Microalbumin
RBC
Albumin
Potassium

Figure S34

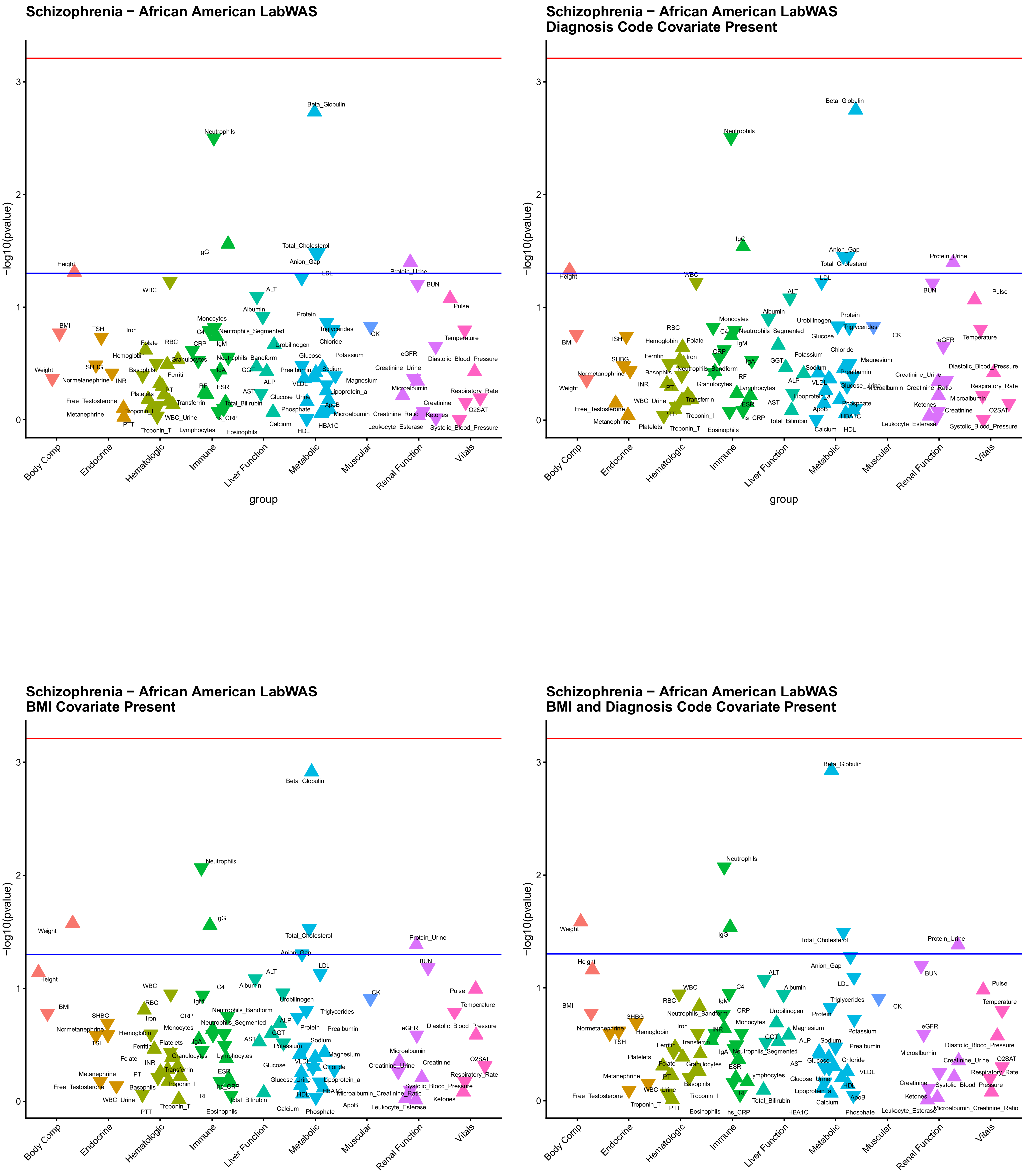
Schizophrenia − African American LabWAS
Schizophrenia − African American LabWAS
Diagnosis Code Covariate Present
Schizophrenia − African American LabWAS
BMI Covariate Present
Schizophrenia − African American LabWAS
BMI and Diagnosis Code Covariate Present
−log10(pvalue)
group
Body Comp
Endocrine
Hematologic
Immune
Liver Function
Metabolic
Muscular
Renal Function
Vitals
